# Discovering the genetic architecture of sleep regulation: genome-wide association study of device-measured sleep traits

**DOI:** 10.1101/2025.07.17.25331692

**Authors:** Laura Portas, Hang Yuan, Lina Cai, Karl Smith-Byrne, Stefan van Duijvenboden, Simon D. Kyle, David Ray, Joanna MM Howson, Aiden Doherty

**Author notes:** **Corresponding author:** Laura Portas, PhD, Big Data Institute Old Road Campus, Oxford, UK OX3 7LF.

## Abstract

Sleep is essential for health and regulated by genetic and environmental factors. We performed genome-wide association studies of device-measured sleep duration, efficiency, and REM/NREM phases in 80,013 UK Biobank participants using accelerometer measurements separately validated against polysomnography. We identified 20 autosomal loci, 12 of which were novel, and report the first genome-wide significant signals for REM and NREM sleep duration. *MEIS1* showed strong opposing effects on REM and NREM durations and is intolerant to loss-of-function mutations, suggesting a regulatory role. Functional enrichment implicated chromatin remodelling, lipid metabolism, and metal ion homeostasis; tissue enrichment highlighted regions including the hypothalamus and frontal cortex. Sex-stratified analyses identified distinct loci: *FOXP2* and *NRXN3* in females and *LRP1B*, *NPBWR2*, and *PABPC4* in males. Mendelian randomization supported associations between shorter sleep duration and higher cardiometabolic risk. Our findings highlight novel sex- and phase-specific regulators of human sleep architecture, offering biological insights and potential therapeutic targets.

## Introduction

Sleep is essential for overall health and well-being, with disruptions linked to a higher risk of cardiometabolic ^1^, and mental health conditions ^2^. Sleep is regulated by a complex architecture that includes two main phases: rapid eye movement (REM) sleep, which accounts for 20-25% of total sleep, and non-rapid eye movement (NREM) sleep, which makes up 75-80% of total sleep. NREM consists of three stages ^3^ with distinct brain activity patterns and is associated with restorative processes, while REM is characterised by desynchronised brain activity, muscle atonia, and rapid eye movements, playing a critical role in memory consolidation and emotion regulation. Disruptions to sleep structure, duration, continuity, and timing can lead to inadequate sleep, and are associated with a range of adverse health outcomes ^1,2^.

Despite the well-described neural circuitry and neurochemistry underpinning sleep state transitions and the coordination between the sleep and circadian systems, the full spectrum of mechanisms governing sleep regulation, particularly the genetic factors influencing sleep traits and their impact on health, remain poorly understood ^4^. Sleep behaviours are influenced by a combination of environmental, lifestyle, habitual and genetic factors, with genetics playing a significant role in the variability of sleep traits. For example, studies have identified associations between genetic variation and sleep duration ^5–9^, sleep-wake preferences ^10–12^, ease of waking ^13^, and sleep disturbances ^14–16^, emphasising the heritable nature of sleep homeostasis.

Previous large-scale genome-wide association studies (GWAS) on sleep traits have been limited by the absence of measurements of REM and NREM sleep. Most relied on self-reported data, which are prone to bias and lack the precision needed to capture sleep architecture complexity ^17^. Recent advances in self-supervised machine learning techniques now make it possible to infer REM and NREM sleep traits from accelerometer datasets, a capacity previously considered unattainable ^18^. Leveraging such data provides a valuable opportunity to deepen our understanding of sleep biology, support personalized medicine, and guide targeted interventions for sleep-related disorders.

In this study we analysed four device-derived sleep traits from a large subset of UK Biobank participants. By incorporating accelerometer-based estimates of both REM and NREM sleep, traits not previously explored in GWAS, we aim to uncover their genetic architecture and address a major gap in sleep genomics.

## Results

### Study sample characteristics

We performed a genome-wide association study (GWAS) of over 9.8 million common variants in 80,013 UK Biobank participants to identify loci associated with four accelerometer-derived sleep traits (Supplementary Figure 1).

Participants had a mean age of 63 years (standard deviation (SD) = 7.8) and 56% were women (Supplementary Table 1). The cohort was relatively healthy and active, with a mean body mass index (BMI) of 26.6 kg/m² (SD = 4.5), an average daily step count of 9,481, and moderate to high socioeconomic status: 44% reported a high level of education, 57% were never smokers, and 49% consumed alcohol more than three times per week.

On average participants slept 6.8 hours per night (SD = 0.9), including 1.5 hours of REM sleep (SD = 0.6) and 5.3 hours of NREM sleep (SD = 0.9). Sleep efficiency averaged 82.0%, indicating relatively good sleep continuity. Women tended to have better sleep efficiency but also reported greater use of sleep-influencing medications, particularly antidepressants. Overall, 3.7% of participants reported taking medications that affect sleep, (e.g., hypnotics, psychotropics), and 0.1% had documented movement disorders, including restless legs syndrome.

### Heritability, polygenicity, and correlation between sleep traits

SNP-based heritability (h^2^_SNP_) estimates for the sleep traits ranged from 9% (sleep efficiency) to 13% (REM and night-time sleep). Genomic inflation was minimal (λ= 1.13-1.19), with LD score regression intercepts ≤ 1.02, suggesting limited confounding. The ratio of intercepts suggested that only 4-8% of the observed inflation could be attributed to sources other than polygenicity (Supplementary Figure 2). Night-time sleep showed strong genetic (r_g_ = 0.79, P < 2.2 × 10^-16^) and phenotypic (r = 0.79, P < 2.2 × 10^-16^) correlation with NREM sleep. Moderate genetic correlations were also observed between night-time sleep and sleep efficiency (r_g_ = 0.45, P < 2.2 × 10^-16^) and REM sleep (r_g_ = 0.43, P < 2.2 × 10^-16^), mirrored by phenotypic correlations of r = 0.42 (P < 2.2 × 10^-16^) and r = 0.36 (P < 2.2 × 10^-16^), respectively. Sleep efficiency was genetically correlated with REM (r_g_ = 0.22, P = 9.5 × 10^-6^) and NREM sleep (r_g_ = 0.35, P = 1.9 × 10^-12^), with corresponding phenotypic correlations of r = 0.18 (P < 2.2 × 10^-16^) and r = 0.31 (P < 2.2 × 10^-16^), respectively. Finally, REM and NREM sleep showed modest negative genetic (r_g_ = -0.21, P = 1.7 × 10^-5^) and phenotypic (r = -0.29; P < 2.2 × 10^-16^) correlations (Supplementary Figure 3).

### SNP-level analysis

We identified 20 independent autosomal loci associated with sleep traits at genome-wide significance (P < 5.0 × 10^−8^), including 12 novel signals (Table 1; Figure 1; Supplementary Figures 4 and 5; Supplementary Table 2). Six loci were associated with night-time sleep duration, of which three were novel (2p16.1, 6p22.3, 7q31.1). For sleep efficiency, one locus was identified at 7q11.22, previously linked to total sleep duration but not sleep efficiency. This study is the first GWAS for REM and NREM sleep, and we identified five loci for REM sleep (1p21.3, 3p11.1, 11q13.2, 11q13.4, and 22q13.1) and three for NREM sleep (13q14.2, 14q22.3, and 15q23), all novel and not previously implicated in sleep-related traits. A signal at 8q24.3 was associated with both REM and NREM sleep. No significant associations were detected on chromosome X. Multi-trait analysis of GWAS (MTAG) did not yield additional loci beyond those identified in single-trait analyses, likely due to modest genetic correlations between traits and the high power of the individual GWAS (Supplementary Table 3; Supplementary Figure 3).

**Figure 1.**
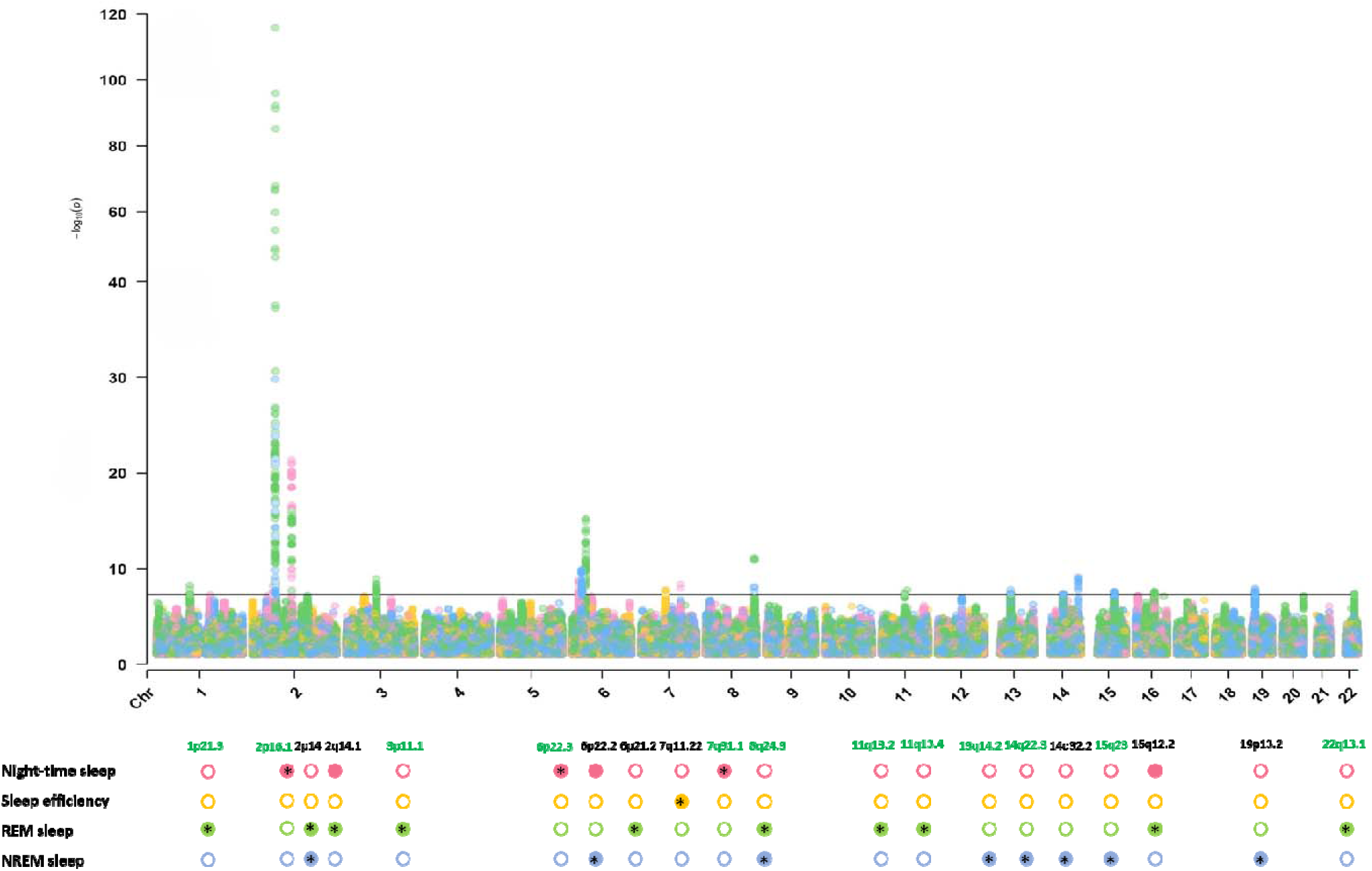
Combined Manhattan plot of GWAS results for the four device-measured sleep traits in 80,013 UK Biobank participants. Coloured circles represent genome-wide significant signals for each trait, while empty circles denote non-significant signals. Aserisks (*) indicate signals not previously associated with the same or equivalent sleep traits in GWAS. The loci highlighted in green contain signals that have never been associated with any sleep-related traits.

**Table 1.** Genome-wide significant (P < 5 × 10^-8^) loci associated with sleep traits, in subjects of European ancestry in the UK Biobank.

| Locus | Lead SNP | Position | Most severe | Nearest | Allele | EAF | Info | Beta | SE | P | Most likely causal SNP |
| --- | --- | --- | --- | --- | --- | --- | --- | --- | --- | --- | --- |
| Night-time sleep duration, minutes/day |  |  |  |  |  |  |  |  |  |  |  |
| 2p16.1 | rs11125769 | 59,310,698 | intergenic variant | *FANCL* | A/C | 0.645 | 1.00 | -1.705 | 0.294 | 6.51E-09 | rs11125769 (0.24) |
| 2q14.1 | rs199993536 | 114,082,628 | intergenic variant | *PAX8* | T/TA | 0.782 | 0.98 | -3.297 | 0.341 | 4.14E-22 | rs199993536 (0.43) |
| 6p22.3 | rs112507172 | 19,096,689 | intergenic variant | *RNF144B* | A/G | 0.752 | 1.00 | 1.954 | 0.324 | 1.73E-09 | rs112507172 (0.22) |
| 6p22.2 | rs6940638 | 27,046,250 | upstream gene variant | *H2BC11* | A/G | 0.771 | 1.00 | 1.813 | 0.331 | 4.54E-08 | rs79887176 (0.08) |
| 7q31.1 | rs13237149 | 113,871,993 | intron variant | *FOXP2* | T/A | 0.686 | 0.98 | -1.794 | 0.305 | 4.27E-09 | rs13237149 (0.62) |
| 16q12.2 | rs62033413 | 53,830,055 | intron variant | *FTO* | C/G | 0.585 | 1.00 | 1.568 | 0.284 | 3.39E-08 | rs186842218 (0.06) |
| Sleep efficiency, % |  |  |  |  |  |  |  |  |  |  |  |
| 7q11.22 | rs2006810 | 69,902,152 | intron variant | *AUTS2* | T/C | 0.604 | 0.99 | 0.245 | 0.044 | 1.76E-08 | rs3113257 (0.14) |
| REM sleep, minutes/day |  |  |  |  |  |  |  |  |  |  |  |
| 1p21.3 | rs11165579 | 96,759,720 | intron variant | *LINC01787* | A/G | 0.706 | 1.00 | 1.124 | 0.193 | 5.42E-09 | rs11165579 (0.09) |
| 2p14 | rs113851554 | 66,750,564 | intron variant | *MEIS1* | G/T | 0.943 | 0.93 | 9.051 | 0.394 | 1.22E-116 | rs113851554 (1.00) |
|  | rs4544423* | 66,750,017 | intron variant | *MEIS1* | T/G | 0.404 | 1.00 | 1.126 | 0.177 | 1.94E-10 |  |
|  | rs182588061* | 66,757,709 | intron variant | *MEIS1* | G/T | 0.990 | 0.83 | 3.812 | 0.670 | 1.25E-08 |  |
| 2q14.1 | rs199993536 | 114,082,628 | intergenic variant | *PAX8* | T/TA | 0.782 | 0.98 | -1.784 | 0.215 | 9.86E-17 | rs199993536 (0.27) |
| 3p11.1 | rs6779623 | 87,849,642 | intergenic variant | *HTR1F* | G/C | 0.705 | 0.99 | 1.178 | 0.193 | 1.11E-09 | rs6779623 (0.20) |
| 6p21.2 | rs4714163 | 38,438,771 | intron variant | *BTBD9* | T/C | 0.708 | 1.00 | -1.566 | 0.193 | 5.48E-16 | rs4714163 (0.38) |
| 8q24.3 | rs2542425 | 140,632,384 | intron variant | *KCNK9* | T/C | 0.554 | 1.00 | 1.214 | 0.177 | 6.66E-12 | rs888346 (0.64) |
|  | rs888346* | 140,713,897 | intron variant | *KCNK9* | C/T | 0.474 | 0.98 | 0.980 | 0.177 | 3.32E-08 |  |
| 11q13.2 | rs10680443 | 66,685,778 | intron variant | *PC* | G/GG | 0.646 | 0.99 | -1.021 | 0.185 | 3.24E-08 | rs10680443 (0.16) |
| 11q13.4 | rs7116582 | 73,732,100 | intron variant | *C2CD3* | G/A | 0.974 | 1.00 | -3.075 | 0.545 | 1.72E-08 | rs7116582 (0.95) |
| 16q12.2 | rs62033400 | 53,811,788 | intron variant | *FTO* | A/G | 0.603 | 1.00 | 1.003 | 0.180 | 2.33E-08 | rs62033417 (0.08) |
| 22q13.1 | rs6001802 | 40,543,215 | intron variant | *TNRC6B* | G/A | 0.745 | 0.98 | -1.116 | 0.203 | 3.71E-08 | rs761126143 (0.08) |
| Non-REM sleep, minutes/day |  |  |  |  |  |  |  |  |  |  |  |
| 2p14 | rs113851554 | 66,750,564 | intron variant | *MEIS1* | G/T | 0.943 | 0.93 | -6.917 | 0.602 | 1.58E-30 | rs113851554 (1.00) |
| 6p22.2 | rs12215241 | 27,023,081 | intergenic variant | *H2BC11* | G/A | 0.772 | 1.00 | 2.052 | 0.319 | 1.28E-10 | rs371182726 (0.19) |
| 8q24.3 | rs2542423 | 140,635,688 | intron variant | *KCNK9* | G/C | 0.556 | 0.99 | -1.571 | 0.272 | 7.49E-09 | rs2542423 (0.47) |
| 13q14.2 | rs7491366 | 47,928,776 | intergenic variant | *HTR2A* | T/C | 0.661 | 0.99 | 1.610 | 0.285 | 1.65E-08 | rs7491366 (0.21) |
| 14q22.3 | rs1209087 | 55,493,220 | intron variant | *WDHD1* | C/T | 0.429 | 0.99 | -1.502 | 0.273 | 3.67E-08 | rs1882747021 (0.05) |
| 14q32.2 | rs7146019 | 99,734,984 | intron variant | *BCL11B* | A/G | 0.429 | 0.97 | -1.708 | 0.277 | 7.12E-10 | rs7146019 (0.23) |
| 15q23 | rs12902804 | 67,988,658 | intron variant | *MAP2K5* | G/A | 0.503 | 1.00 | -1.504 | 0.270 | 2.42E-08 | rs12902804 (0.08) |
| 19p13.2 | rs12973413 | 9,942,222 | upstream gene variant | *UBL5* | A/G | 0.447 | 0.98 | -1.554 | 0.272 | 1.13E-08 | rs1292220483 (0.40) |

E= effect allele, A= alternative allele, Chr= chromosome, EAF= effect allele frequency, INFO= imputation quality, PIP= posterior inclusion probability. Analyses are adjusted for age, sex, centre, season, genetic ancestry and genotyping array. <sup>§</sup> Distance to the transcription start site (TSS) was considered for non intronic variants. \* Secondary signals identified after conditional SNP association analysis (1Mb window around the lead SNP).

Conditional analyses identified additional independent signals within several loci. At 2p14, two intronic variants in *MEIS1*, rs4544423 (β = 1.13 min/night; P = 1.9 × 10^-10^) and rs182588061 (β = 3.81 min/night; P = 1.2 × 10^-8^), remained significantly associated with REM sleep after adjusting for the lead SNP rs113851554 (β = 9.05 min/night; P = 1.2 × 10^-116^). Similarly, at 8q24.3, a secondary intronic variant in *KCNK9* (rs888346; β = 0.98 min/night; P = 3.3 × 10^-8^) was identified after conditioning on rs2542425 (β = 1.21 min/night; P = 6.7 × 10^-12^). Fine-mapping analyses highlighted coding variants with high posterior probability of causality (PIP ≥ 95%). At 2p14, the intronic variant rs113851554 in *MEIS1* was associated with both REM (β = 9.05 min/night; P = 1.2 × 10^-116^) and NREM sleep (β = -6.92 min/night; P = 1.6 × 10^-30^) and had a posterior probability of causality (PIP) of 100%. This gene is highly intolerant to loss-of-function mutations (pLI = 1), supporting an essential biological role.

At 11q13.4, the intronic variant rs7116582 in *C2CD3* (PIP = 95%), was associated with REM sleep (β = -3.07 min/night; P = 1.7 × 10^-8^), potentially influencing gene expression or splicing.

Sensitivity analyses excluding individuals with documented movement disorders or those using medications known to affect sleep, as well as models adjusted for BMI, did not materially change the top associations. However, some signals lost genome-wide significance due to reduced statistical power (Supplementary Table 4).

### Sex-specific genetic effects

Sex-stratified GWAS identified loci with sex-specific associations, suggesting sex-based regulation of sleep traits (Supplementary Table 4; Supplementary Figure 6). Four loci showed significant sex heterogeneity: 7q31.1 for night-time sleep (I^2^ = 91.20, P = 0.001), and 1p21.3 (I^2^ = 79.80, P = 0.026), 2p14 (I^2^ = 96.60, P = 6.47 × 10^-8^), and 6p21.2 (I^2^ = 83.20, P = 0.015) for REM sleep. Additional sexually dimorphic loci, not genome-wide significance in the overall analysis, were also identified (Supplementary Table 5). In females, an intergenic variant (14:78496241_CA_C) was significantly associated with night-time sleep duration (β = 3.63 min/night; P = 4.16 × 10^-8^) and lies ∼140 kb from the *NRXN3*. Another female-specific intergenic variant (4:104463040_AT_A), was associated with sleep efficiency (β = 38.6%; P = 4.0^-8^), and is located ∼178 kb from *TACR3*. In males, three intronic variants showed significant associations with REM sleep: rs568010150 in *LRP1B* (β = -1.96 min/night; P = 1.1 × 10^-8^), rs398111 in *NPBWR2* (β = 1.71 min/night; P = 3.3 × 10^-8^), and rs1180331 in *PABPC4* (β = -1.58 min/night; P = 1.1 × 10^-8^). For NREM sleep, a female-specific association was found for rs1799252 in *FOXP2* (β = -2.16 min/night; P = 6.4 × 10^-9^), while an intergenic variant (rs6465150) was associated in males (β = -3.69 min/night; P = 2.4 × 10^-8^), and is located ∼78 kb from the *ZNF804B* gene. Given the mean age of female participants (62.1 years), most were likely postmenopausal, and 38.2% reported ever using hormone replacement therapy (HRT), which may partly contribute to the observed sex-specific associations. These findings highlight sex-specific genetic contributions to sleep traits and suggest distinct biological pathways, warranting further investigation of hormonal and regulatory mechanisms.

### Gene-level analysis

Gene-based analyses revealed significant enrichment in pathways related to chromatin organization, metal ion homeostasis (particularly copper, zinc, cadmium), and lipid metabolism (Supplementary Table 6).

Chromatin-related pathways such as nucleosome assembly and DNA-protein interactions were enriched across all sleep traits, while fatty acid metabolism showed a specific association with REM sleep.

Tissue enrichment analysis highlighted multiple brain regions for night-time sleep (e.g., cerebellum, cortex, basal ganglia, hypothalamus, amygdala) and the pituitary gland, suggesting a role for neuroendocrine regulation (Supplementary Figure 7). REM sleep was enriched in the frontal cortex, general cortex, and cingulate cortex, while NREM sleep showed enrichment in the cerebellum and nucleus accumbens.

Gene-level aggregation also identified ten additional trait-associated loci beyond SNP-level analysis. These included one locus associated with night-time sleep duration (near *SGCZ*), three with sleep efficiency (near *FAM150B*, *PBRM1*, and *ITGA2B*), four with REM sleep (near *CAMTA1*, *PKP4*, *SKOR1*, and *TBX6*), and two with NREM sleep (near *SGCZ*, and *CAND1*) (Supplementary Table 7).

### Chromatin interaction, eQTL mapping, and eQTL colocalization analysis

Integrated analyses suggest that sleep regulation involves an interplay between neurological, metabolic, gastrointestinal, and immune systems (Supplementary Tables 8 and 9, Supplementary Figures 8-9).

Chromatin interaction mapping linked several loci to candidate genes, including *PSD4*, *PAX8*, *FTO*, and *IRX3* for night-time sleep, and *MEIS1*, *HTR1F*, and *HTR2A* for REM and NREM sleep.

eQTL mapping revealed tissue-specific gene expression patterns, and colocalization analysis supported regulatory roles for several key genes. For night-time sleep, colocalization at 2q14.1 with *FOXD4L1* and *CBWD2* in thyroid tissue suggests a neuroendocrine component, in line with prior associations at PAX8, a thyroid lineage transcription factor identified in early sleep GWAS. For REM sleep, *HTR1F* colocalized in spleen, thyroid, adipose, and heart tissue, reflecting potential immune-metabolic contributions. For NREM sleep, relevant colocalizations included *BTN2A2* (within the MHC region), *WDHD1*, and *PIN1-DT*, with signals in pancreas, esophageal mucosa, and frontal cortex. Although the MHC locus requires cautious interpretation due to its complex LD structure, *BTN2A2*, while related to the immunoglobulin genes, has distinct roles in lipid metabolism, further emphasising links between immune and metabolic functions in deep, restorative sleep.

No significant sex-specific colocalizations were detected, likely due to limited power. Nonetheless, these findings highlight a complex regulatory network involving both central and peripheral pathways. For example, *HTR1F* was regulated by variants such as rs6779623 and rs56169023, which influence gene expression in peripheral tissues, underlining the relevance of non-neuronal contributions to sleep physiology. Similarly, genes such as *WDHD1* and *SKOR1*, enriched in gastrointestinal and metabolic tissues, reinforce the idea that peripheral systems contribute substantially to NREM sleep. Together, these results point to multi-system mechanisms where neurological, metabolic, and immune processes converge to regulate sleep duration and architecture.

### Sleep traits and health outcomes

LD score regression (LDSC) revealed significant genetic correlation between sleep traits and various health outcomes (Figure 2). Night-time sleep was negatively correlated with BMI, type 2 diabetes mellitus (T2DM), coronary artery disease (CAD), and heart failure (HF), while sleep efficiency was negatively correlated with BMI and T2DM. REM sleep showed negative genetic correlations with several cardiometabolic traits (HF, CAD, T2DM, BMI) and biological age, but a positive correlation with Alzheimer’s disease. In contrast, NREM sleep was positively associated with biological age. Pairwise colocalization analyses identified shared loci between sleep traits and T2DM, notably at 16q12.2 (FTO, rs55872725, PP.H4 = 0.83 for night-time sleep and PP.H4 = 0.82 for REM sleep), and 22q13.1 (TNRC6B, rs6001802, PP.H4 = 0.80 for REM sleep), including genes previously linked to chronotype, suggesting a possible role for circadian misalignment. Given previous associations of these loci with obesity, we performed additional analyses using BMI-adjusted summary statistics to assess potential confounding. Adjustment for BMI did not materially affect association at 22q13.1 (rs6001802, PP.H4 = 0.79 for REM sleep), but at 16q12.2, the colocalization signal weakened (decrease in PP.H4, increase in PP.H3), suggesting partial mediation by BMI. Similar patterns were observed for other traits: colocalization signals at 16q12.2 for BMI (rs1558902, PP.H4 = 0.85 for night-time sleep and PP.H4 = 0.84 for REM sleep) and heart failure (16:53803223_G_A, PP.H4 = 0.85 for night-time sleep and PP.H4 = 0.87 for REM sleep) was reduced after adjustment (PP.H4 < 0.5). Likewise, the association with HbA1c (16:53798523_G_A, PP.H4 = 0.75 for night-time sleep) was no longer supported post-adjustment.

**Figure 2.**
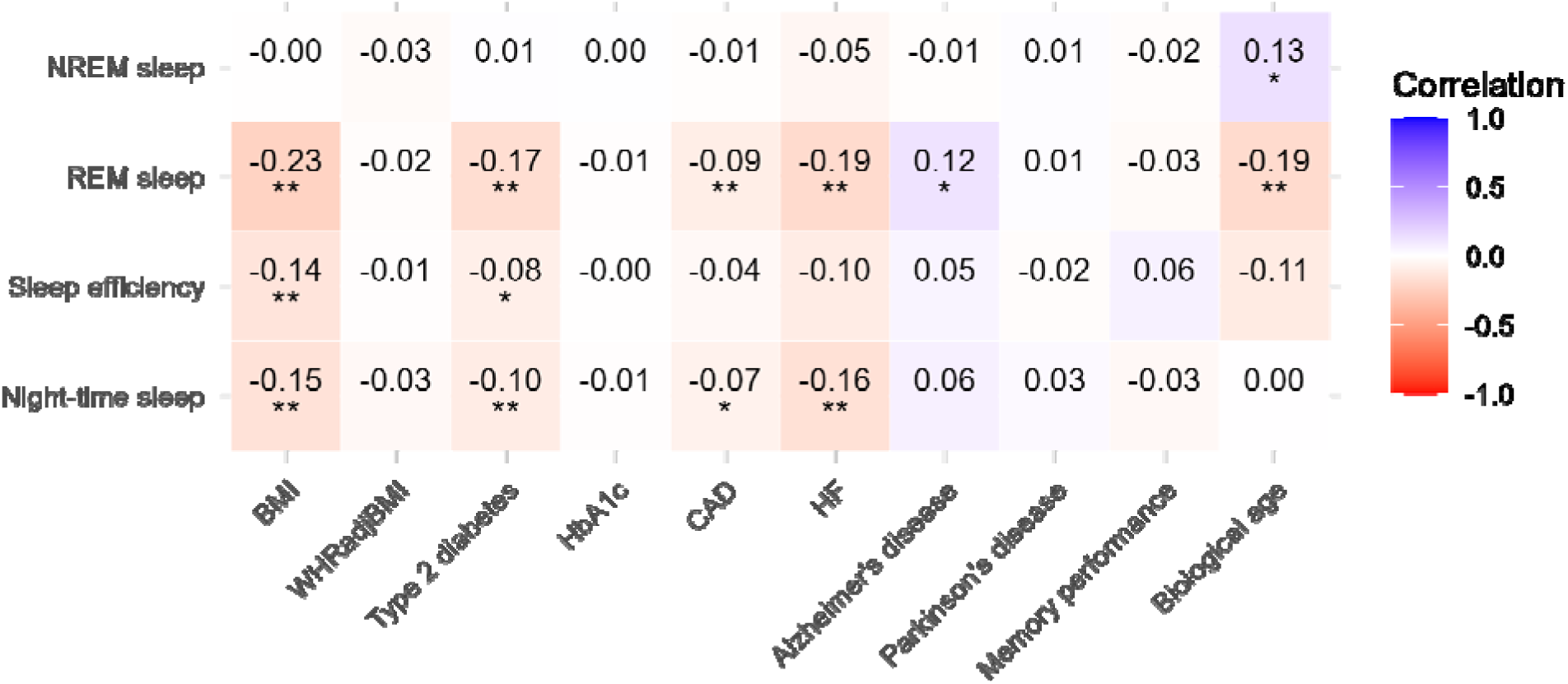
Genetic correlations of sleep traits with other phenotypes. The genetic correlation coefficient was calculated with LDSC and is denoted by color scale from −1 (red; negatively (anti-)correlated) to +1 (blue; positively correlated). Single asterisk indicates two-sided P ≤ 0.05 and double asterisk indicates two-sided P ≤ 0.01. Abbreviations: BMI: body mass index; WHRadjBMI: waist-hip ratio adjusted for BMI, HbA1c: hemoglobin A1c; CAD: coronary artery disease; HF: heart failure.

Mendelian randomization analyses (Supplementary Table 10; Figure 3) supported beneficial effects of longer night-time sleep on BMI (β = -0.366 per hour, se = 0.084, P = 7.4 × 10^-6^), HbA1c (β = -0.090, se = 0.024, P = 5.7 × 10^-5^), T2DM risk (Odds Ratio (OR) = 0.610, 95% Confidence Interval (CI) = 0.415-0.894, P = 0.011), and biological age (β = -0.414, se = 0.120, P = 7.6 × 10^-4^). Longer REM sleep showed suggestive associations with improved memory (β = 0.036, se = 0.018, P = 0.014), lower HF (OR = 0.710, 95% CI = 0.529-0.953, P = 0.020) and CAD risk (OR = 0.845, 95% CI = 0.717-0.997, P = 0.043), and reduced HbA1c (β = -0.084, se = 0.036, P = 0.015), but also a higher waist-to-hip ratio adjusted for BMI (WHRadjBMI) (β = 0.072, se = 0.024, P = 0.003). Higher sleep efficiency was linked to lower BMI (β = -2.322, se = 0.692, P = 8.00 × 10^-4^ per 1% increase), but higher WHRadjBMI (β = 2.322, se = 0.733, P = 0.001). Reverse MR indicated that higher BMI causally reduced REM (β = -3.716 min/night per kg/m^2^, se = 0.463, P = 1.0 × 10^-15^), night-time sleep (β = -3.004 min/night per kg/m^2^, se = 0.746, P= 5.6 × 10^-5^), and sleep efficiency (β = -0.490% per kg/m^2^, se = 0.110, P = 5.0 × 10^-6^). Higher Alzheimer’s risk was associated with shorter night-time sleep (β = -1.271 min/night, se = 0.423, P = 0.003), and NREM sleep (β = -1.358 min/night, se = 0.418, P = 0.001).

**Figure 3.**
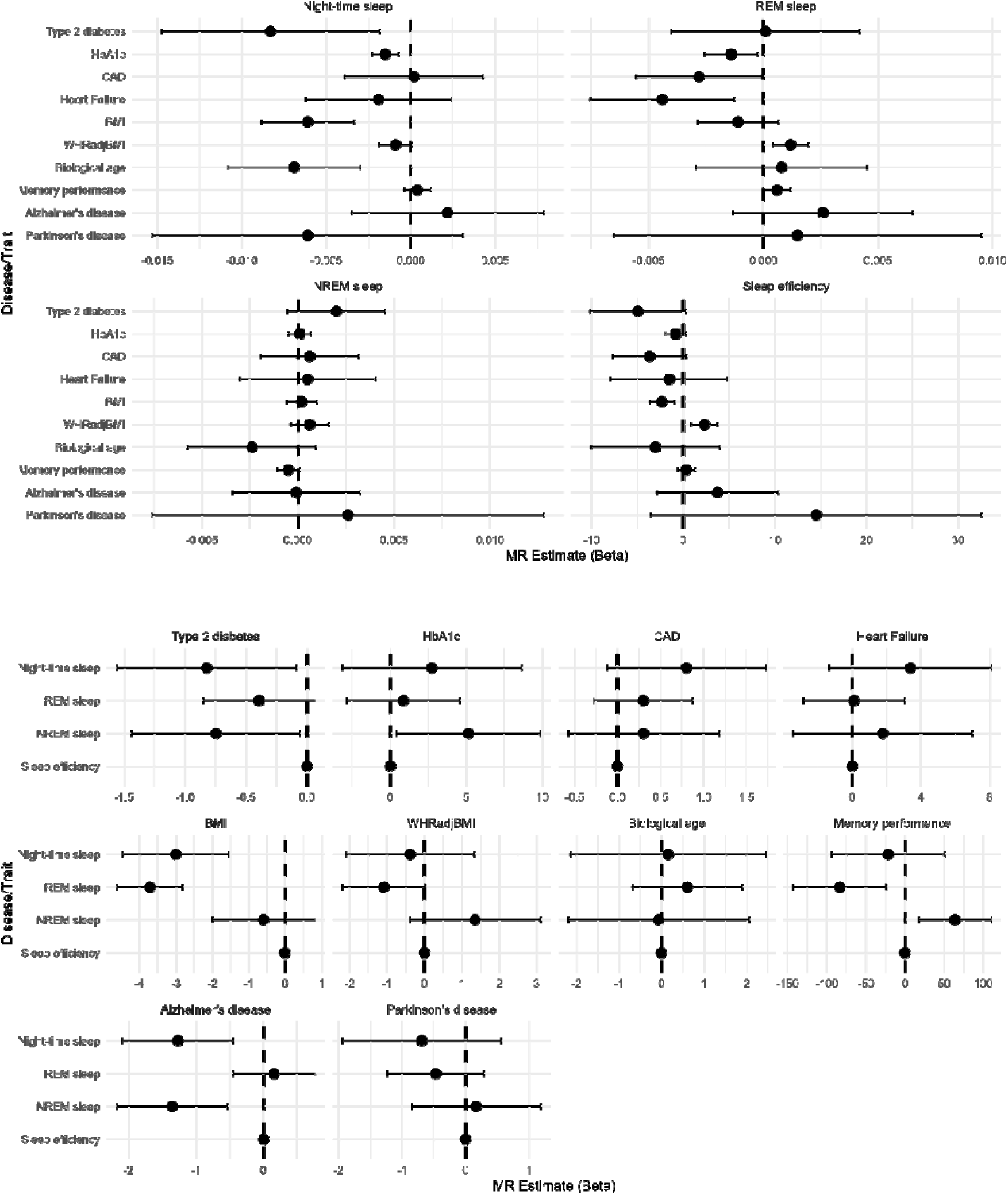
Bidirectional Mendelian randomization results: causal effects of sleep traits on health outcomes and vice versa. Results shown correspond to the inverse-variance weighted (IVW) method with random effects. Abbreviations: HbA1c: hemoglobin A1c; CAD: coronary artery disease; BMI: body mass index; WHRadjBMI: waist-hip ratio adjusted for BMI. Traits shown at the top of each panel represent the exposure in the Mendelian randomization analysis.

Suggestive evidence pointed to a higher risk of type 2 diabetes being associated with lower NREM sleep duration (β = -0.747 min/night, se = 0.352, P = 0.034), as well as better memory performance with shorter REM sleep duration (β = -83.11 min/night, se = 30.28, P = 0.006) and longer NREM sleep (β = 63.98 min/night, se = 23.75, P = 0.007).

## Discussion

In this genome-wide association study of over 80,000 UK Biobank participants, we investigated four accelerometer-derived sleep traits, including REM and NREM sleep, traits not previously examined in GWAS. We identified 20 autosomal loci, 12 of which are novel, providing the first genome-wide significant associations for REM and NREM sleep. These findings reveal phase-specific genetic contributions to human sleep architecture and suggest links between sleep traits and cardiometabolic outcomes.

We identified an association between a novel locus at 3p11.1 and REM sleep, with chromatin and eQTL analyses highlighting *HTR1F* as a regulatory candidate. This gene encodes a serotonin receptor and shows eQTL signals in metabolically relevant tissues including adipose, heart, and pancreas. While serotonin’s role in sleep regulation is well established, our findings suggest that *HTR1F* may also contribute to the link between REM sleep and cardiometabolic traits. Our findings are further supported by a recent study ^19^ identifying a regulatory variant at *HTR1F* associated with obstructive sleep apnea in non-obese individuals, where the same locus shows eQTL activity in adipose and heart tissues as well as in neurons. This work also links *HTR1F* variation to increased night-time arousals, reinforcing its role in sleep architecture and central sleep regulation. Additionally, we provide new insight into *MEIS1*’s phase-specific role in sleep architecture ^9^. A likely causal variant (rs113851554, PIP = 100%) at 2p14, previously associated with chronotype, insomnia, and RLS ^20^, was linked to increased REM and decreased NREM sleep. *MEIS1* is highly intolerant to loss-of-function mutations (pLI = 1), supporting its essential role in neuronal development and motor control. Despite excluding individuals with known movement disorders, undiagnosed cases cannot be ruled out, although the MEIS1 association remained robust.

Sex-stratified analyses identified loci associated with sleep traits in males and females, including *FOXP2*, *NRXN3*, and *LRP1B*. These genes are involved in neural development, hormonal signalling, or show sex-differential expression in brain tissue, supporting biological plausibility ^21–23^. For example, FOXP2 is modulated by androgens and affects social behaviour in a sex-specific manner ^21,22^, while NRXN3 influences GABAergic inhibition differently in males and females ^23^. Another notable example is TACR3, located near a female-specific intergenic signal for sleep efficiency, which encodes a receptor for neurokinin B and plays a critical role in hypothalamic regulation of puberty onset, neuroendocrine signalling, and cardiovascular control. Prior GWAS have rarely examined sex differences in sleep genetics; our findings support a sexually dimorphic architecture for REM and NREM sleep, meriting further investigation. In Mendelian randomization analyses, genetically shorter night-time sleep was associated with higher BMI, HbA1c, and T2DM risk, supporting previous epidemiological and genetic studies ^7,24,25^. Importantly, we also found suggestive causal effects of longer REM sleep on reduced risk of HF and CAD, outcomes for which the role of sleep architecture has been less well characterised ^26,27^. Higher sleep efficiency was associated with lower BMI, but also with increased WHRadjBMI, suggesting a complex, potentially region-specific influence on adiposity.

Our study benefits from the use of objectively measured sleep traits from wrist-worn accelerometers in a large sample (n = 80,013), and the exclusion of confounding factors such as shift work or daylight saving effects. Sleep stages were inferred using a deep neural network trained on over 1,100 nights of polysomnography, ensuring robust classification of REM and NREM periods. This is the first GWAS to focus specifically on these stages.

Nonetheless, limitations include the predominantly European ancestry and relatively healthy, high-SES profile of UK Biobank participants ^28^. Although this cohort is not fully representative, findings are generally reproducible in other populations ^29^. Nevertheless, independent replication in larger and more diverse cohorts will be important to validate these associations. Moreover, REM/NREM estimates were indirectly inferred from movement data, potentially introducing measurement imprecision. However, the observed associations with age, sex, and BMI support the validity of the derived traits.

In summary, we identified novel genetic loci for REM and NREM sleep and demonstrated phase-specific and sex-specific genetic influences on sleep architecture. These findings deepen our understanding of sleep genetics and its potential impact on metabolic and cardiovascular health. They also underscore the value of targeting both sleep duration and quality in preventive strategies aimed at reducing the burden of cardiometabolic disease. Importantly, the identification of candidate genes and regulatory elements lays the groundwork for future functional studies to dissect causal mechanisms underlying sleep regulation.

## Methods

### Participants and sleep traits

The study population was drawn from the UK Biobank cohort, a large, longitudinal, population-based study involving over 500,000 individuals ^30^. For the present analyses, we focused on a subset of 103,680 participants who wore a wrist-worn accelerometer for 7 days between 2013 and 2015 ^31^ and restricted the sample to individuals of European ancestry. Participants were excluded if their data could not be calibrated, exhibited unrealistically high values (e.g., average vector magnitude > 100 mg), or had insufficient wear-time (i.e., less than 3 days or no recorded wear in every hour of the 24-hour period).

Additional exclusions were applied to shift workers and individuals with data impacted by daylight saving time transitions (Supplementary Figure 1).

Sleep traits, including night-time sleep duration, sleep efficiency, and the durations of REM and NREM sleep, were derived using a self-supervised deep recurrent neural network for sleep stage classification, trained on over 1,100 nights of concurrent laboratory-based polysomnography and accelerometry data. The Kappa score for REM/NREM/wake classification was 0.32. The model’s predictions and polysomnography data show a difference of 48.2 minutes for sleep duration, -17.1 minutes for REM duration, 31.1 minutes for NREM duration, and 9.2% for sleep efficiency ^18^.

### Genotyping, imputation, and quality control

Genotyping and imputation procedures for UK Biobank participants have been previously described ^30^. In brief, genotyping was performed using the Affymetrix UK BiLEVE Axiom array for the initial ∼50,000 participants and the Affymetrix UK Biobank Axiom Array for approximately 450,000 participants. The two arrays are over 95% similar and together cover approximately 820,000 SNP and indel markers (http://www.ukbiobank.ac.uk/). Quality control and imputation of over 90 million SNPs, indels, and large structural variants were carried out centrally. Following imputation, additional SNP-level quality control was performed locally, excluding variants with an imputation INFO score < 0.3 or a minor allele frequency (MAF) < 0.01. Individual-level quality control was also conducted to remove samples identified as outliers based on heterozygosity, missingness, mismatched sex (where discrepancies between reported and genetically inferred sex were detected, indicating potential sample processing errors), or sex chromosome aneuploidy (Supplementary Figure 1).

### Genome-wide association analyses

All association tests were performed using REGENIE v.3.1.2 ^32^, which implements a linear mixed model (LMM) to account for population structure and relatedness, allowing for the inclusion of related individuals and thus enhancing the power to detect genetic associations. We analysed autosomal and X chromosome data assuming an additive genetic model, adjusted for age at accelerometry, sex, study centre, season of activity monitor wear, and genotyping array. Additionally, the first 10 principal components (PCs) of the white European subset included in the analysis were incorporated as covariates to control for subtle differences in ancestry. For the X chromosome analysis, male genotypes were coded as diploid (0,2).

To further improve statistical power in detecting genetic loci associated with sleep, we performed multitrait analysis of genome-wide association studies (MTAG) alongside single-trait GWAS. MTAG performs a meta-analysis of GWAS summary statistics across genetically correlated traits, addressing sample overlap to enhance power and precision ^33^.

### Sensitivity analyses

To evaluate whether observed associations for individual variants were influenced by specific subgroup characteristics, we performed several sensitivity analyses: (1) stratification by sex (males only, females only) (2) adjustment for body mass index (BMI; UK Biobank data field 21001), (3) exclusion of individuals with movement disorders (Supplementary Table 11), and (4) exclusion of individuals taking medications known to influence sleep (Supplementary Table 11).

### Genomic loci characterisation, linkage disequilibrium score regression, and fine-mapping

Independent genome-wide significant signals were identified using an LD-based clumping procedure implemented in PLINK v.1.9 ^34^. This procedure clusters SNPs based on their linkage disequilibrium (LD) with nearby SNPs and their P values. We used the following parameters: index SNP P value threshold of 5 × 10^-8^ (--clump-p1) and clumped SNP P value threshold of 0.05 (--clump-p2), with a physical distance threshold of 1,000 kb (--clump-kb) and an LD threshold of 0.01 (--clump-r2). SNPs within each clump were represented by the index SNP. Variant annotation was performed using the Ensembl Variant Effect Predictor (VEP) tool ^35^.

To identify additional independent signals in regions of association, conditional SNP association analysis was conducted using the Genome-wide Complex Trait Analysis (GCTA) v.1.24.4 tool ^36^ Any lead SNPs within known high-LD regions on chromosomes 6 (28.48-33.45 MB) and 17 (43.5-45.5 MB) were treated as a single locus in the GCTA analysis. Independent genome-wide significant signals were classified as novel if the lead SNP was not in LD (r² ≥0.1) with any previously reported variant associated with the same sleep trait. Previously reported associations were identified through the GWAS Catalog and cross-referenced with Open Targets Genetics ^37^, which integrates results from UK Biobank, FinnGen, and other sources.

LD score (LDSC) regression was performed for each GWAS to estimate SNP-based heritability, evaluate test statistic inflation, and assess genetic correlations between traits. The regression was performed by regressing the GWAS test statistics (χ^2^) onto each SNP’s LD score, which is the sum of squared correlations between the MAF of the SNP and those of all other SNPs. This regression allows for the estimation of heritability from the slope and can detect residual confounders through the intercept. The LDSC regression intercept accounts for inflation in the χ^2^ statistics that is not due to stratification or other confounding factors. Inflation caused by polygenicity will correlate with LD, while inflation from stratification or relatedness will not ^38^. LD scores and weights were downloaded from LDSC repository (https://data.broadinstitute.org/alkesgroup/LDSCORE) for European populations.

GWAS summary statistics were standardized using the munge function implemented in LDSC. To minimise bias from poorly imputed SNPs, an imputation quality score threshold of > 0.9 was applied. SNPs whose alleles did not match those in the 1000 Genomes reference or were strand-ambiguous were excluded.

Fine-mapping was performed using the “Sum of Single Effects” (SuSiE) Bayesian model ^39^, which is a stepwise conditional analysis that improves upon traditional methods by accounting for the uncertainty in selecting associated SNPs. This approach allows for the detection of multiple causal signals and identifies causal variants even when the SNP with the lowest P value is not the causal variant. To reduce the complexity of the analysis and focus on the most relevant variants, we limited the SNPs to those within a 500 kb window surrounding the lead variant at each locus. In-sample dosage-based LD matrices were computed using PLINK v.1.9 ^34^ and 95% credible sets (CS) were evaluated. A SNP was considered as a likely causal variant if the posterior inclusion probability (PIP) was ≥ 0.80, indicating a high probability of being associated with the trait. This threshold was chosen to prioritise variants with strong evidence of association, although the method does not provide definitive proof of causality.

### Functional downstream analyses

To conduct in silico downstream functional analyses of GWAS results, we used the FUMA (Functional Mapping and Annotation) GWAS platform v.1.5.2 ^40^ which implements the MAGMA (Multi-Marker Analysis of GenoMic Annotation) framework v.1.08 ^41^.

First, functional annotation of all genome-wide significant SNPs and SNPs in LD with them (r2≥0.6) was performed using Annotate Variation (ANNOVAR) enrichment test (gene-based annotation), which annotates the functional consequence of SNPs on Ensemble (v92) protein coding genes (e.g. intron and exon) . Functionally annotated SNPs were subsequently mapped to genes using three strategies: positional mapping (physical distance), expression quantitative trait loci (eQTL) mapping (eQTL association), and chromatin interaction.

For positional mapping, based on annotations obtained from ANNOVAR, the physical distance of each SNP from known protein-coding genes was set to the default window of 10 kb in the human reference assembly (hg19/GRCH37).

eQTL mapping was used to map independent significant SNP and SNPs in LD with them to genes that show a significant association with cis-eQTLs (i.e., allelic variation of the SNP associated with expression levels of the gene). This approach maps SNPs to genes located up to 1MB apart. We considered significant eQTL associations with an FDR < 0.05, as recommended by FUMA.

Chromatin interaction mapping was performed by overlapping independent significant SNPs and SNPs in LD with them with one end of significantly interacting regions in tissue/cell types. These SNPs were then mapped to genes whose promoter regions (by default, 250 bp upstream and 500 bp downstream of the transcription start site) overlap with the other end of the significant interactions. This mapping can involve long-range interactions as there is no distance boundary. FUMA uses data on the 3D structure of chromatin interactions from Hi-C data for 23 tissues and cell types, as well as tissue and cell type data from FANTOM and PsychENCODE. The significance threshold was defined as FDR < 1.0 x 10 in FUMA, based on prior recommendations ^42^.

Furthermore, MAGMA was used to perform gene-based, gene-set and gene-property (tissue gene expression) analyses of the full GWAS summary results. In brief, gene-based analysis computes gene-based P values for SNPs mapped to protein coding genes, using a SNP-wise mode that aggregates SNP P values into a gene-level test statistic. Gene boundaries were determined using NCBI build 37, encompassing 18,877 protein-coding genes. SNPs located within gene boundaries were included in the analysis to derive P values reflecting associations with sleep traits. Linkage disequilibrium within and between genes was assessed using the UK Biobank release2b 10K White British reference panel. A Bonferroni correction was applied to account for multiple testing, setting the genome-wide significance threshold at P ≤ 6.6 × 10^−7^, considering 18,877 genes and four sleep traits.

A competitive gene-set analysis was performed to investigate whether the polygenic signal from the GWASs clustered in specific biological pathways. Competitive tests control for Type 1 error rate and assess whether genes within the set are more strongly associated with sleep traits compared to other genes (39). A total of 10,894 gene-sets from Gene Ontology (40), Reactome (41), and SigDB (42) were analysed for enrichment in sleep measures. Bonferroni correction was again applied to account for the multiple tests across the 10,894 gene sets.

Gene-property analysis was conducted to explore whether tissue-specific expression levels in 30 broad tissue types and 53 specific tissues were predictive of a gene’s association with sleep traits. Tissue types were sourced from the GTEx v8 RNA-seq database (43), and expression values were log2 transformed with a pseudocount of 1 after winsorising at 50, with the average expression value taken from each tissue. Multiple testing was controlled for using Bonferroni correction.

### eQTL colocalization analysis

To assess whether the same variant influences both sleep traits and gene expression, a Bayesian colocalization analysis was performed using GTEx v8 eQTL data. Analyses were conducted with the Coloc method ^43^, implemented in the xQTLbiolinks R package ^44^ (https://github.com/lilab-bioinfo/xQTLbiolinks). Sentinel SNPs were identified for each GWAS using the xQTLanalyze_getSentinelSnp function with parameters P < 5.0 x 10^-8^ and SNP-to-SNP distance > 1 Mb. Genes within 1Mb of sentinel SNPs were considered for analysis (Supplementary Table 12). The GWAS genome version was converted from GRCh37 to GRCh38 for compatibility with GTEx v8.

We assumed a prior probability that a SNP is associated with either sleep or gene expression (default = 1.0 x 10^-4^), and both GWAS and gene expression (default = 1.0 x 10^-5^) for all Coloc analyses. These priors were initially recommended for the analysis of eQTL data ^45^ and are widely adopted in applied practice. The Coloc method calculates posterior probabilities for five hypotheses, including the hypothesis of shared causal variants between sleep traits and gene expression. A posterior probability (PP.H4) ≥ 75% was used as a threshold to indicate strong evidence for colocalization, leading to the prioritization of genes within the corresponding loci. A comprehensive list of tissues, including brain regions, was investigated (Supplementary Table 13).

### LDSC genetic correlation, Mendelian randomization, and pairwise colocalization analysis

To explore the genetic overlap between sleep traits and ten related diseases/traits − type 2 diabetes mellitus (T2DM), hemoglobin A1c (HbA1c), coronary artery disease (CAD), heart failure (HF), body mass index (BMI), waist-to-hip ratio adjusted for BMI (WHRadjBMI), biological age (an estimate of an individual’s age based on biological markers, reflecting overall health status and aging process), memory performance, Alzheimer’s disease (AD), and Parkinson’s disease (PD) − we estimated genetic correlations using LDSC. These traits were chosen for their known or hypothesised links with sleep, particularly concerning cardiometabolic health, aging, and neurodegenerative diseases.

The purpose was to identify potential biological connections that could be explored further with causal inference methods like Mendelian randomization (MR).

To further explore the potential relationships, including possible causal links, between sleep traits and these ten diseases/traits, we conducted a two-sample bidirectional MR analysis. Instrumental variables (IVs) were identified from our sleep GWAS. We included only independent SNPs (r^2^ < 0.01 and window size=1,000 kb) that were genome-wide significant (P < 5 x 10^−8^) and had a minor allele frequency > 0.01. SNPs with weak statistical power (F statistics < 10) were excluded ^46^.

For the outcome datasets, we used publicly available summary-level GWAS data for diseases and traits related to sleep in populations of European ancestry. Where exposure-associated SNPs were unavailable in the outcome dataset, correlated SNPs (r^2^ ≥ 0.9) were used as substitutes. Effect alleles were harmonised across datasets, and incompatible or palindromic SNPs were excluded. Data sources and the list of IVs are summarised in Supplementary Table 14.

The primary MR analysis was performed using the inverse variance weighted (IVW) method with a random-effects model to combine Wald ratios for each SNP ^47^. MR Egger ^48^ and weighted median ^49^ methods were used as complementary approaches to assess directional pleiotropy. The MR-Egger intercept was used to estimate directional pleiotropy, with the slope providing a valid estimate when pleiotropy was detected. The weighted median method was applied as it provides valid causal estimates even when up to 50% of the instruments are invalid. The MR-PRESSO ^50^ global test and MR robust adjusted profile score (MR.RAPS) ^51^ methods were applied to account for pleiotropy and to correct for heterogeneity in instrument-exposure associations and outlier SNPs, thus reducing bias from these sources. MR Steiger filtering was used to assess the most likely direction of effect for each SNP. Outliers and SNPs that were inconsistent with the expected direction of effect were removed, and MR causal estimates were re-evaluated. If only one SNP or fewer than three SNPs were available, the Wald ratio and fixed effects IVW were utilised, respectively. Cochrane’s Q-statistic was used to assess the heterogeneity of SNP effects, with smaller P values indicating higher heterogeneity and potential for directional pleiotropy. This test was consistently applied across all analyses, with caution exercised in cases where the number of SNPs was limited, to avoid over-interpreting results with low statistical power. Bonferroni correction was applied to adjust for multiple comparisons, with a P value threshold of < 0.005 for the ten diseases/traits used as exposures and < 0.012 for the four sleep traits used as exposures, to support evidence for a potential association between sleep traits and each outcome. To be considered robust, results were required to show a statistically significant association (P < 0.005 or 0.012, as per Bonferroni correction) using IVW, with directionally concordant evidence from secondary methods. Results were further deemed reliable if the MR–Egger intercept and MR-PRESSO global test were non-significant (P > 0.05), indicating no evidence of directional pleiotropy or outlier bias. Discrepancies in directionality observed with MR–Egger were interpreted cautiously, as they may reflect the method’s lower statistical power rather than pleiotropic bias. P values between 0.005 and 0.05 or between 0.012 and 0.05 were regarded as suggestive evidence of an association.

All MR analyses were performed using R (version 4.2.0) and the “TwoSampleMR” package (version 0.5.6), with MR-PRESSO and RAPS analyses conducted using the “MRPRESSO” and “MR.raps” R packages.

Approximate Bayes factor (ABF) colocalization analysis was used to assess shared genetic causal variants between sleep traits and related health outcomes. Colocalization was performed using the Coloc method, which allows for the detection of shared causal variants between traits at the same locus ^52^. A posterior probability of PP.H4 ≥ 75% was considered suggestive for colocalization, suggesting a common genetic mechanism driving both traits.

Sensitivity analyses were performed to examine the impact of prior probabilities on our results.

## Data Availability

All data produced in the present study are available upon reasonable request to the authors.

## Acknowledgements

The authors acknowledge funding support from Novo Nordisk. They thank the participants of the UK Biobank, as well as the research teams involved in data collection at baseline and during the accelerometry study.

## Conflicts of Interest

AD is supported by grants from the Wellcome Trust [223100/Z/21/Z], Novo Nordisk, Swiss Re, Health Data Research UK, and the British Heart Foundation Centre of Research Excellence [RE/18/3/34214]; has accepted consulting fees from the University of Wisconsin (NIH R01 grant) and Harvard University (NIH R01 grant); received support for presentations or attendance at several conferences; and has received a donation from SwissRe for accelerometer data collection in the China Kadoorie Biobank. LP is supported by Novo Nordisk. LC and JMMH are full-time employees of Novo Nordisk and hold shares in Novo Nordisk A/S. The other authors declare no competing interests.

## Notes

### Author Declarations

The Ethics Committee of the University of Oxford gave ethical approval for this work. The UK Biobank received ethical approval from the North West Multi-centre Research Ethics Committee (REC reference: 16/NW/0274), and this research was conducted under application number 59070 of the UK Biobank Resource. All participants provided written informed consent.

