## Supplementary Figures for "Discovering the genetic architecture of sleep regulation: genome-wide association study of device-measured sleep traits"

**Supplementary Figure 1**. Participant flow diagram for the GWAS of wearable device-derived sleep measurements in the UK Biobank.

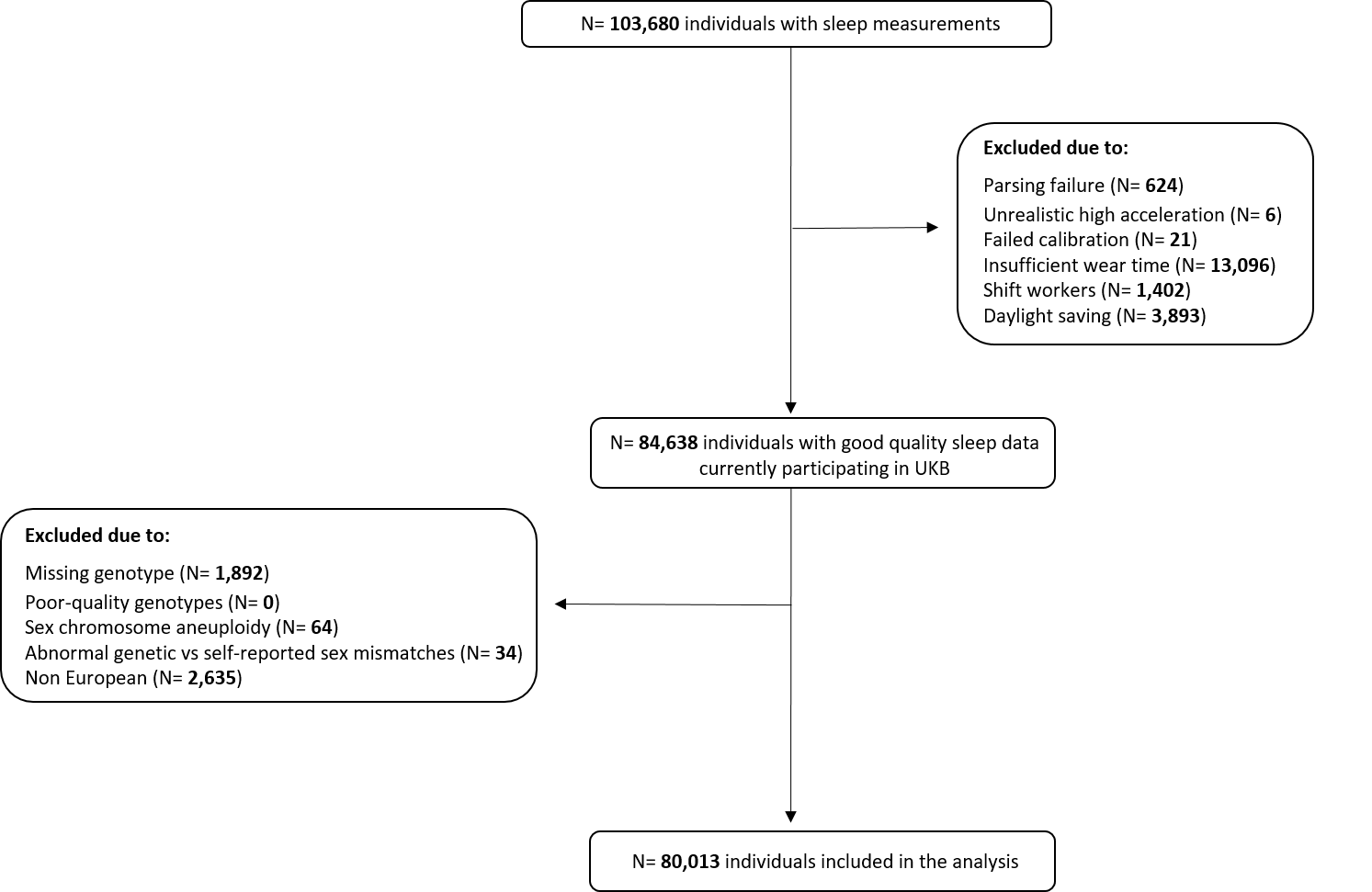

**Supplementary Figure 2**. Q-Q plots and LD Score regression estimates (SNP-based heritability (h2SNP), genomic control lambda (λ), intercept, and ratio) for genome-wide association analysis of sleep traits: a) night-time sleep duration, b) sleep efficiency, c) REM sleep duration, and d) Non-REM sleep duration.

a)

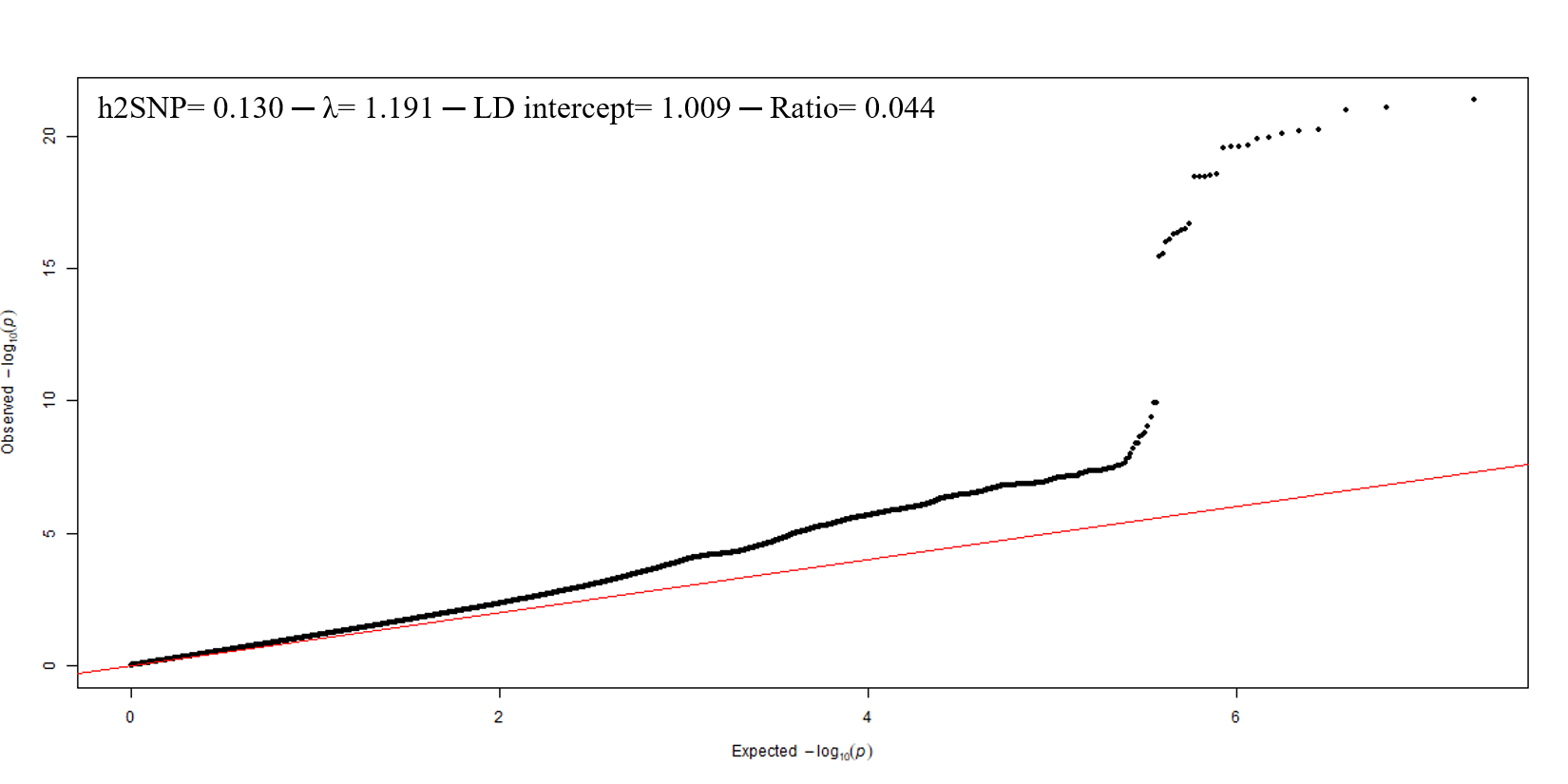

b)

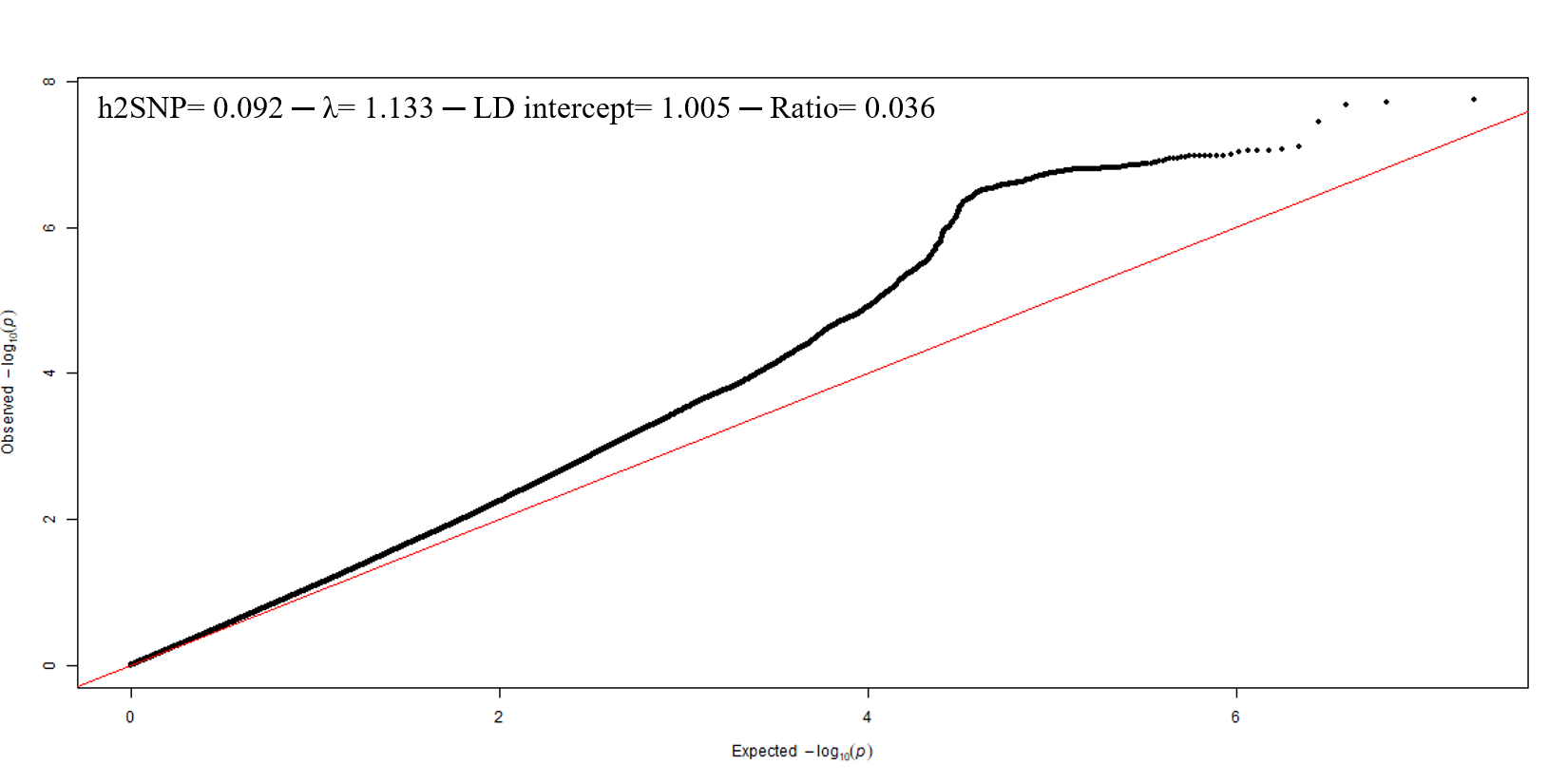

c)

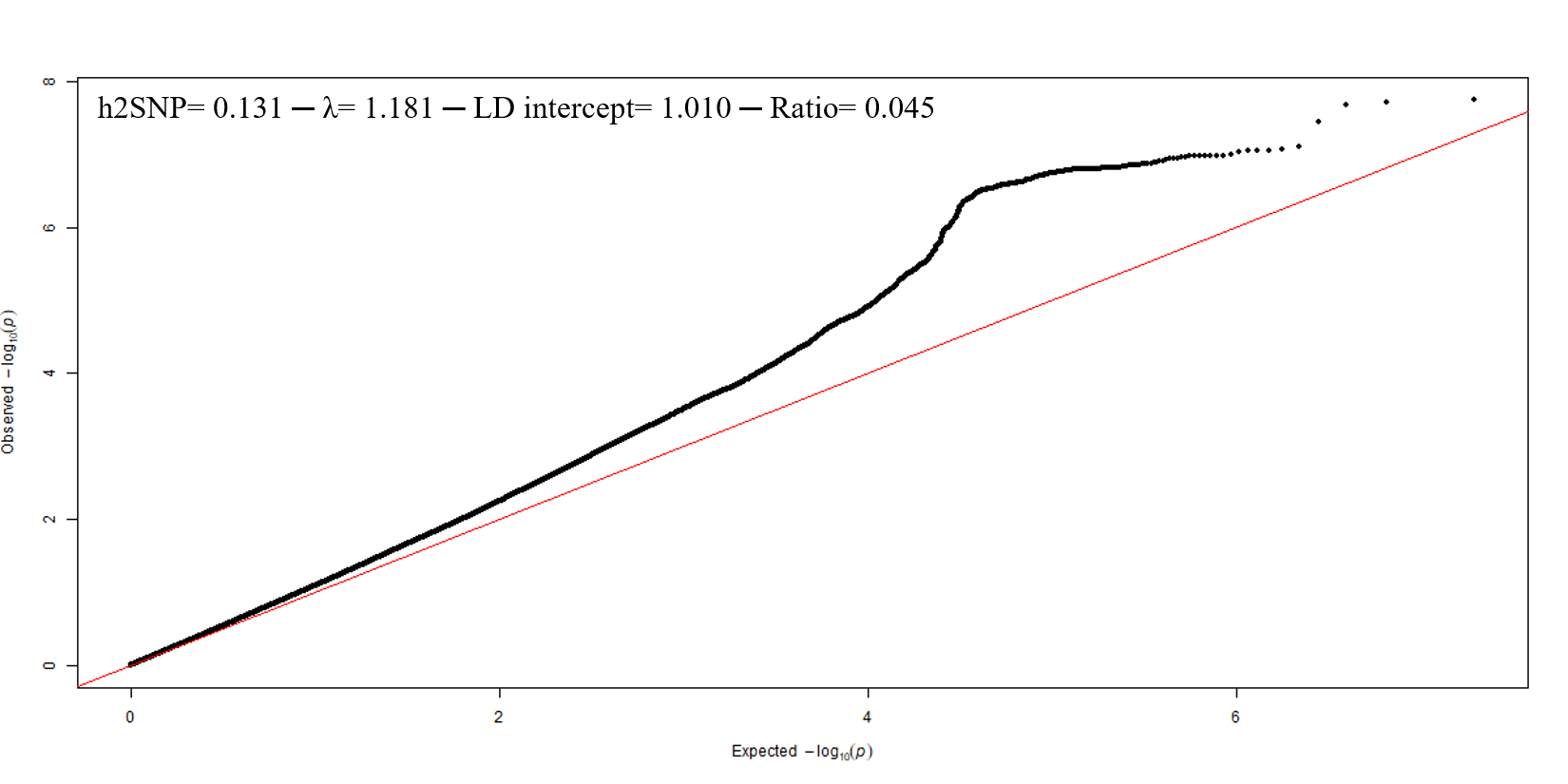

d)

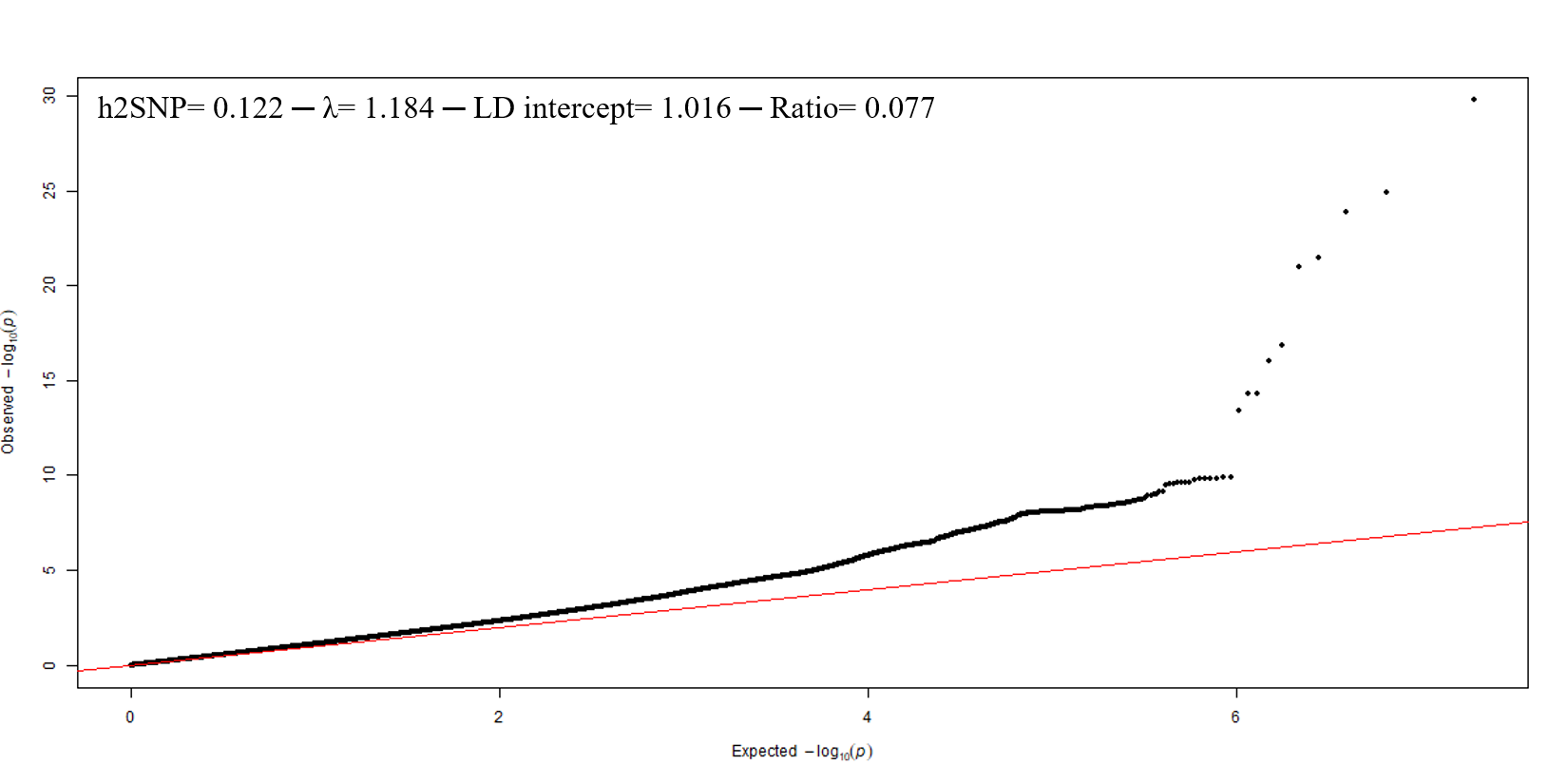

**Supplementary Figure 3**. Phenotypic (Pearson correlation - r) and genetic (measured by LDSC - r_g_) correlation between sleep traits.

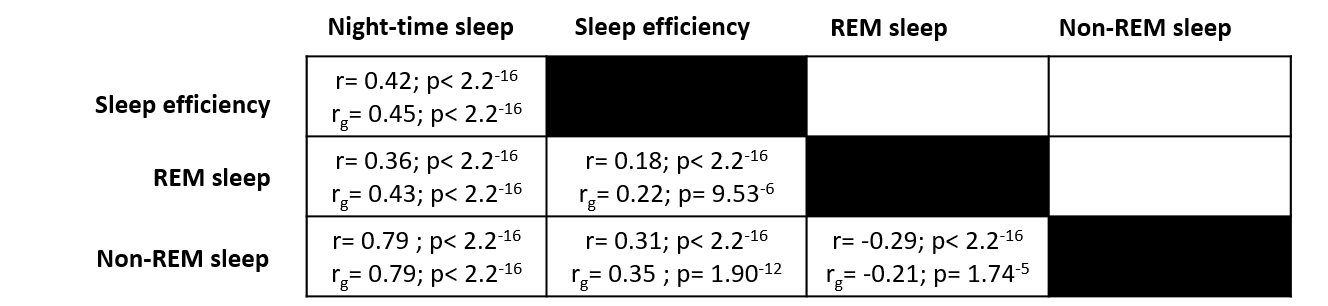

**Supplementary Figure 4**. Manhattan plot of sleep traits: a) night-time sleep duration, b) sleep efficiency, c) REM sleep duration, and d) Non-REM sleep duration.

a)

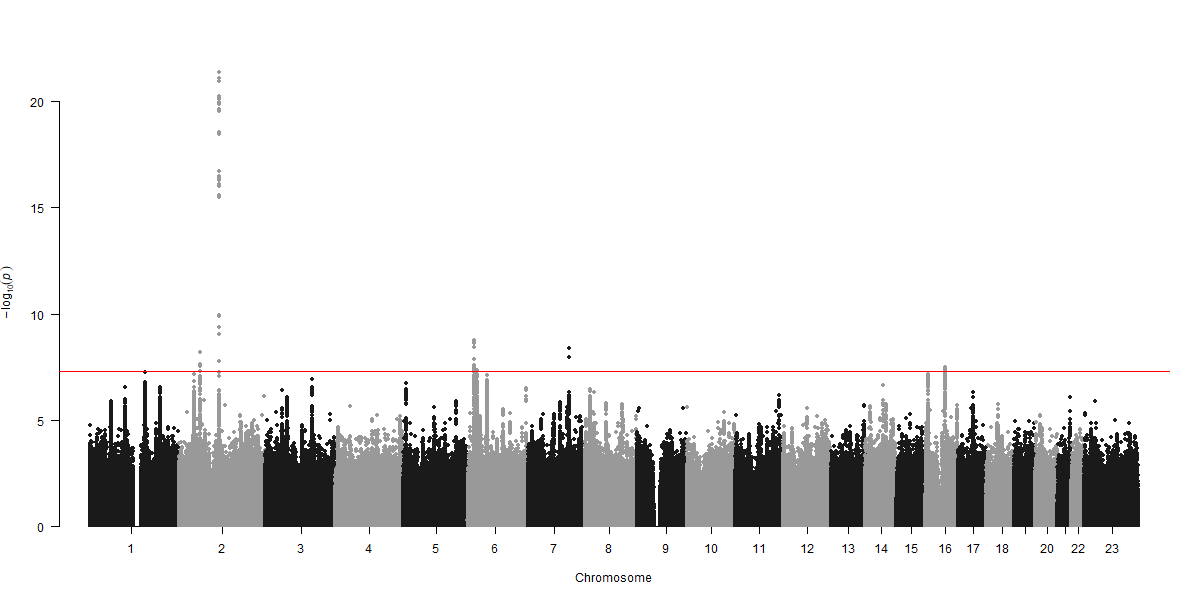

b)

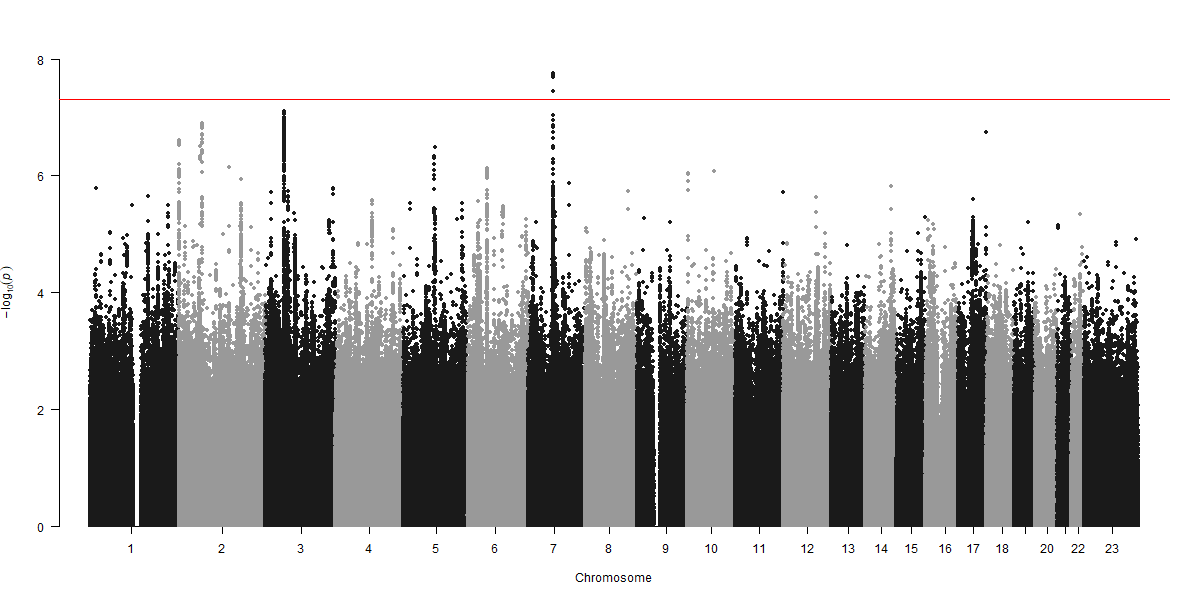

c)

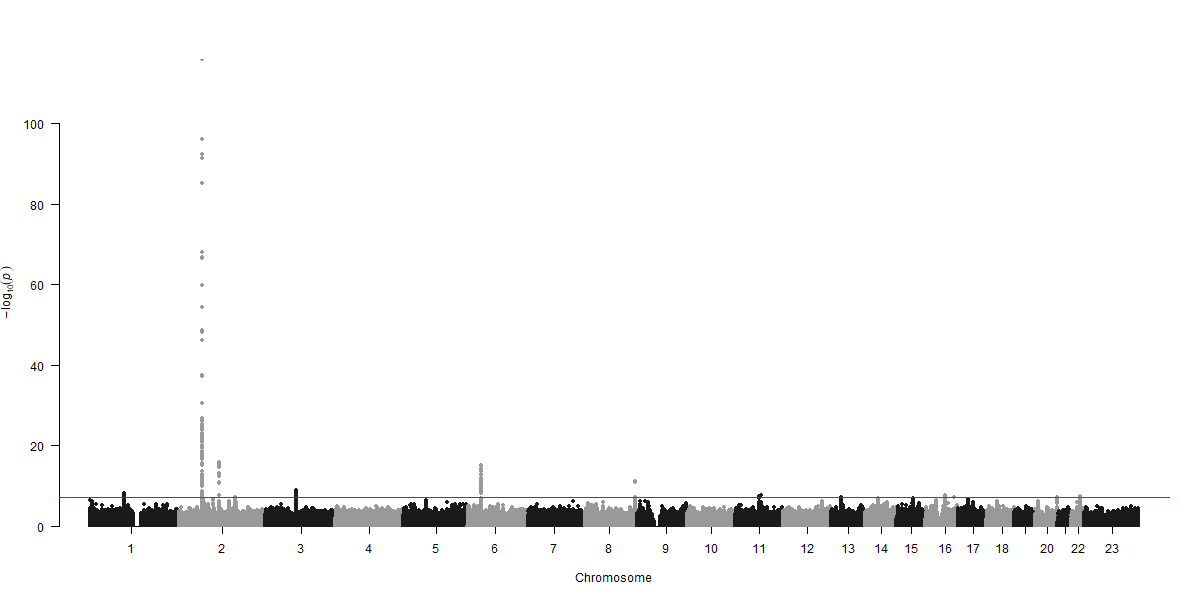

d)

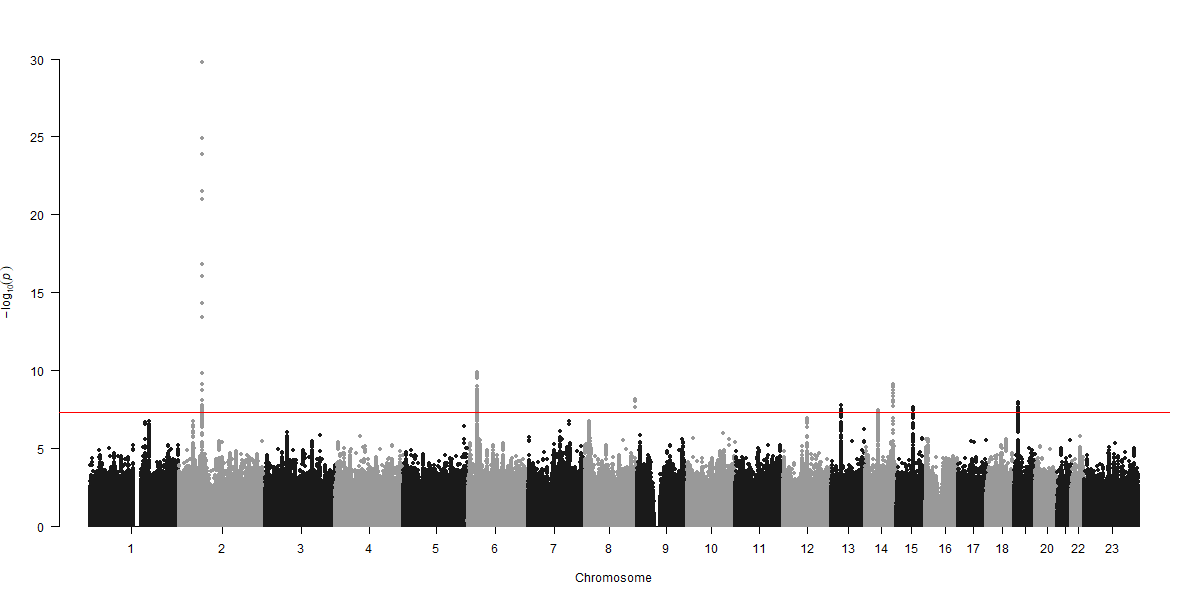

**Supplementary Figure 5**. Locus zoom plots for associated loci across sleep traits (night-time sleep: i) 2p16.1, ii) 2q14.1, iii) 6p22.3, iv) 6p22.2, v) 7q31.1, and vi) 16q12.2; sleep efficiency: vii) 7q11.22; REM sleep: viii) 1p21.3, ix) 2p14, x) 2q14.1, xi) 3p11.1, xii) 6p21.2, xiii) 8q24.3, xiv) 11q13.2, xv) 11q13.4, xvi) 16q12.2, and xvii) 22q13.1; Non-REM sleep: xviii) 2p14, xix) 6p22.2, xx) 8q24.3, xxi) 13q14.2, xxii) 14q22.3, xxiii) 14q32.2, xxiv) 15q23, and xxv) 19p13.2). For loci 2q14.1 and 11q13.2, a proxy SNP in perfect LD was used for plotting due to the unavailability of the lead SNP.

i)

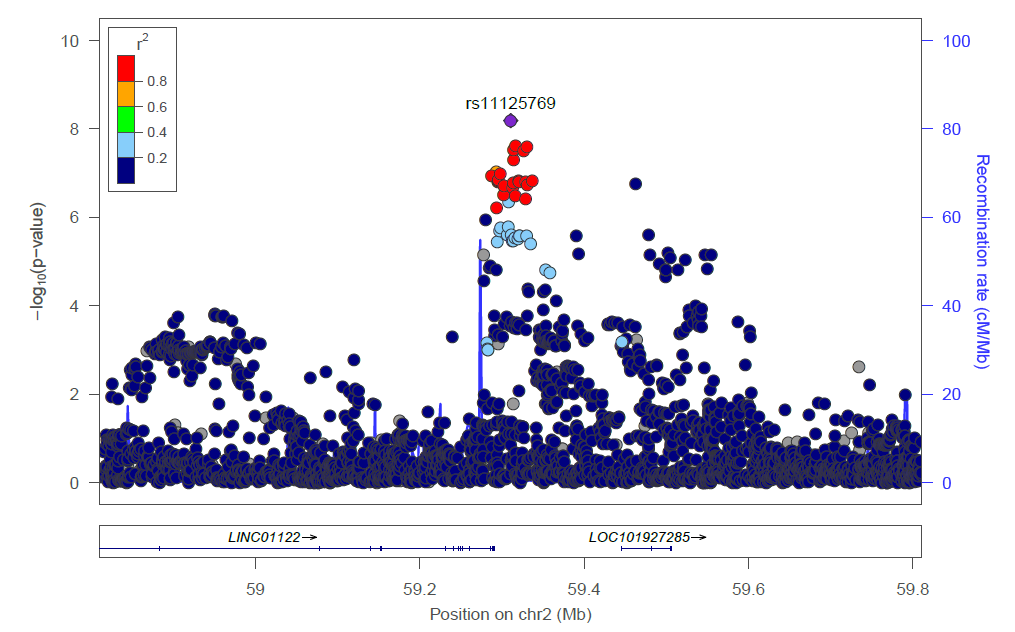

ii)

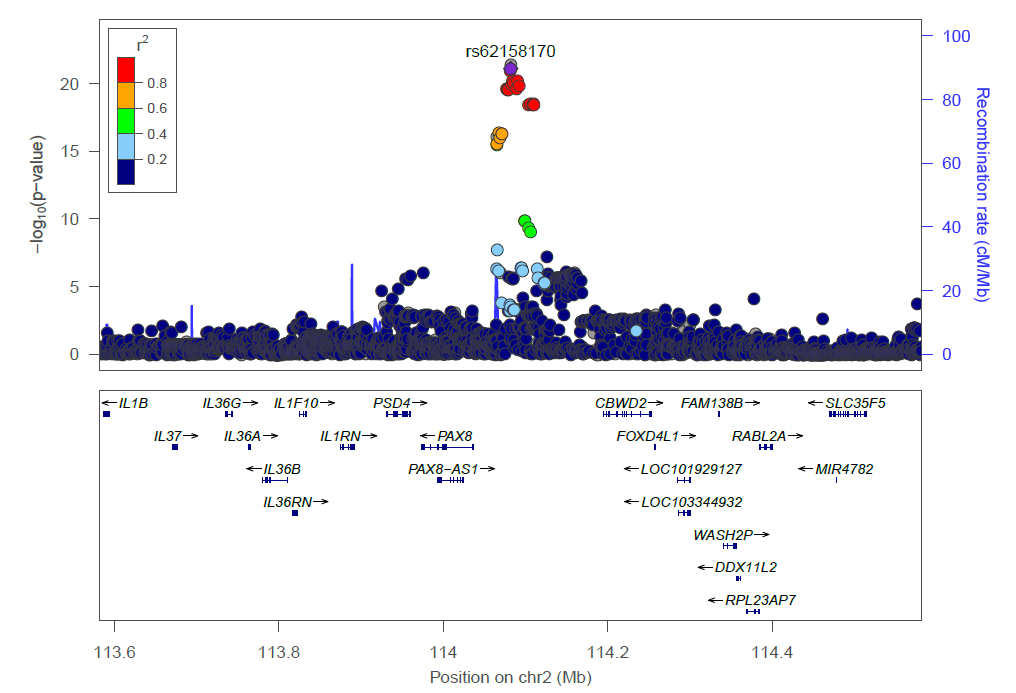

iii)

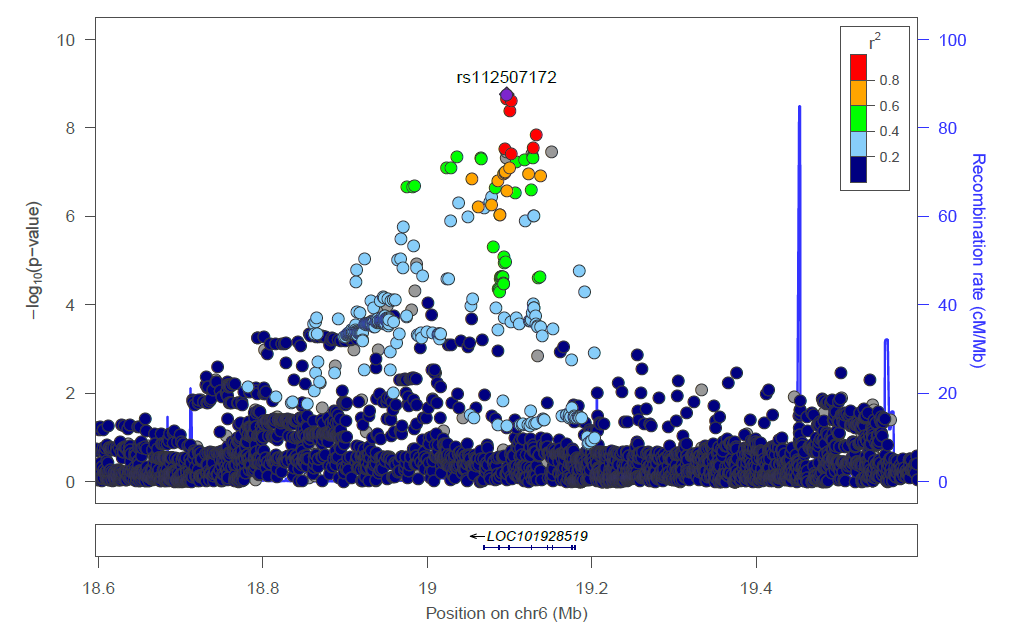

iv)

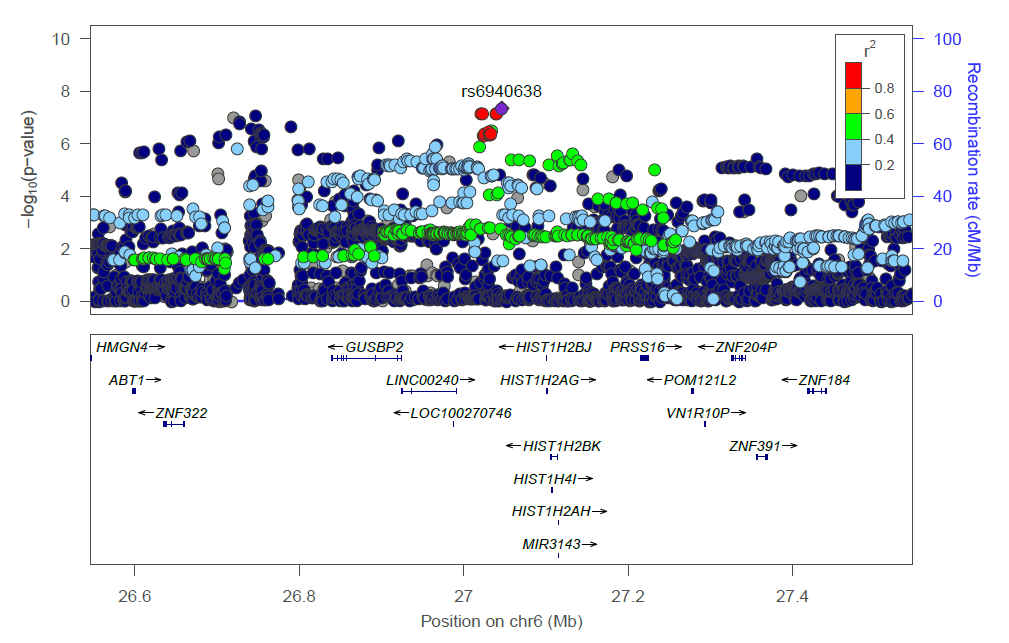

v)

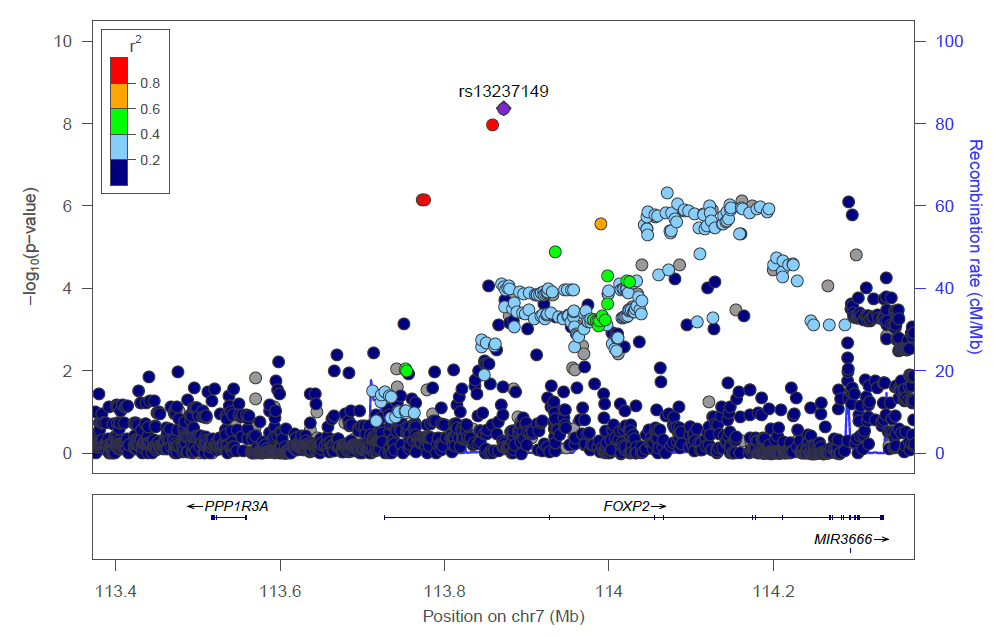

vi)

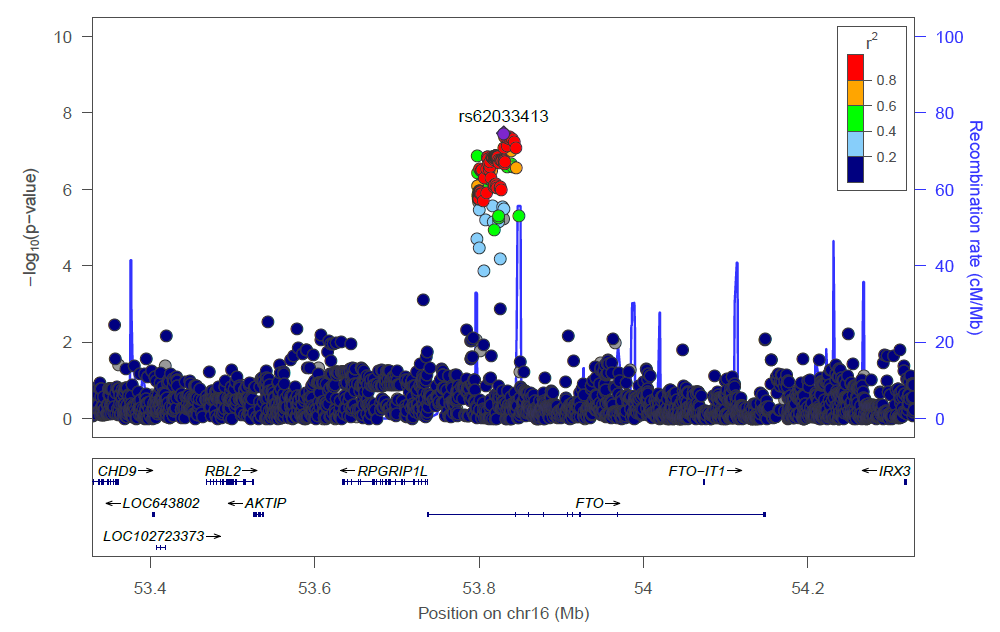

vii)

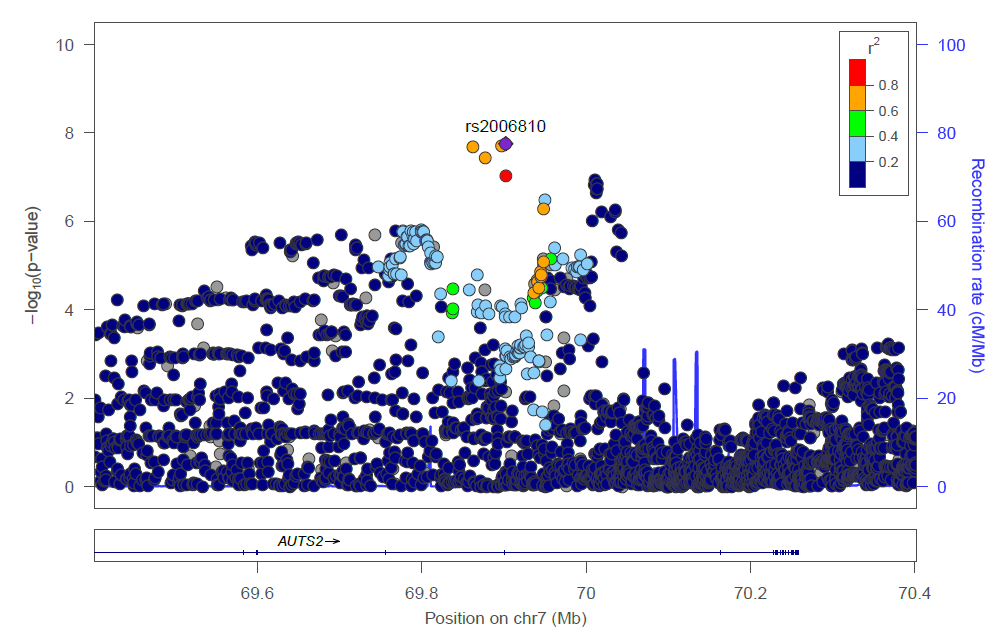

viii)

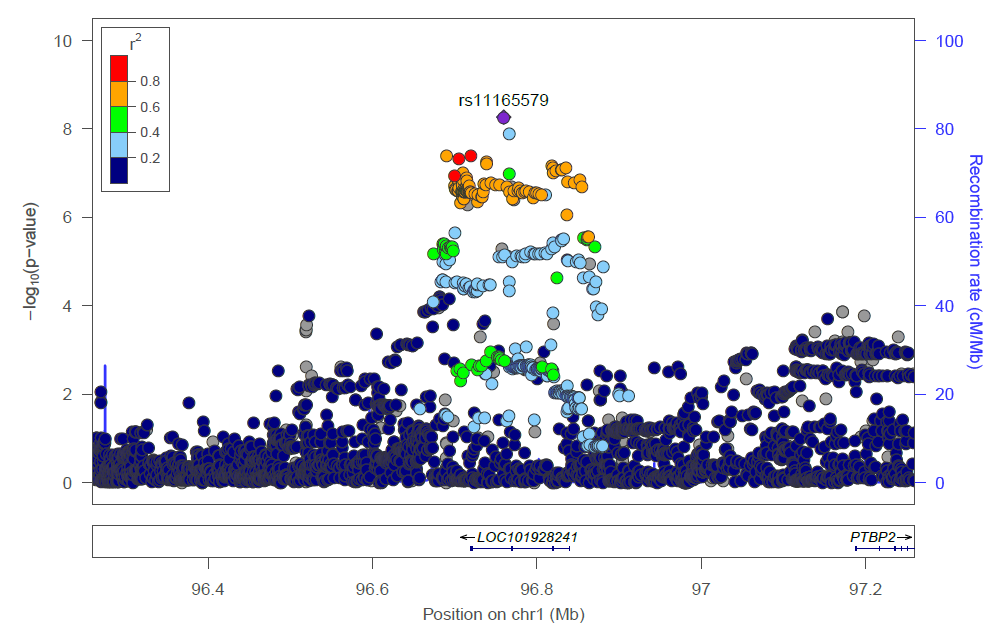

ix)

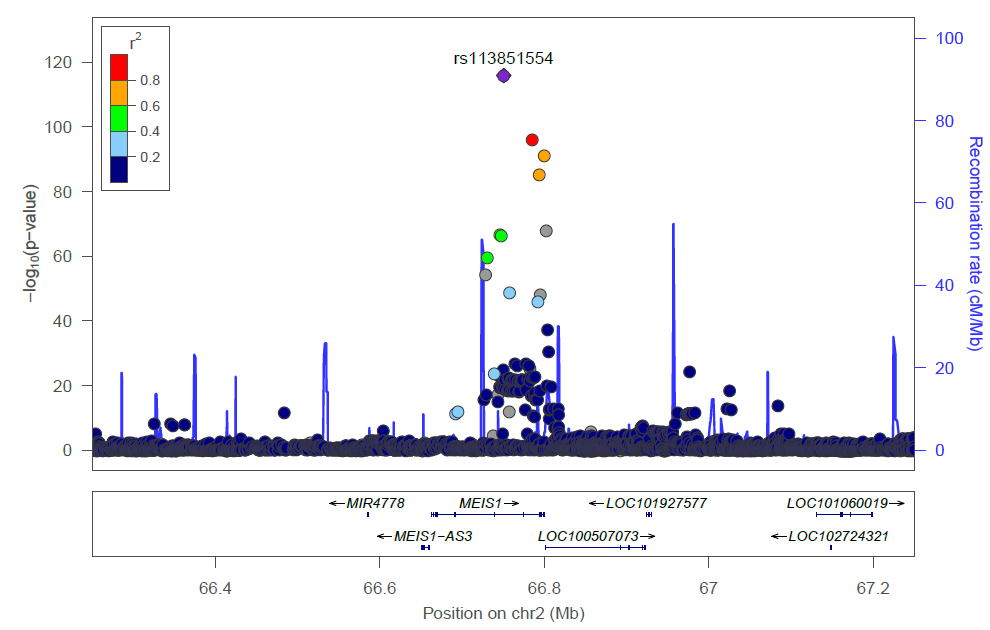

x)

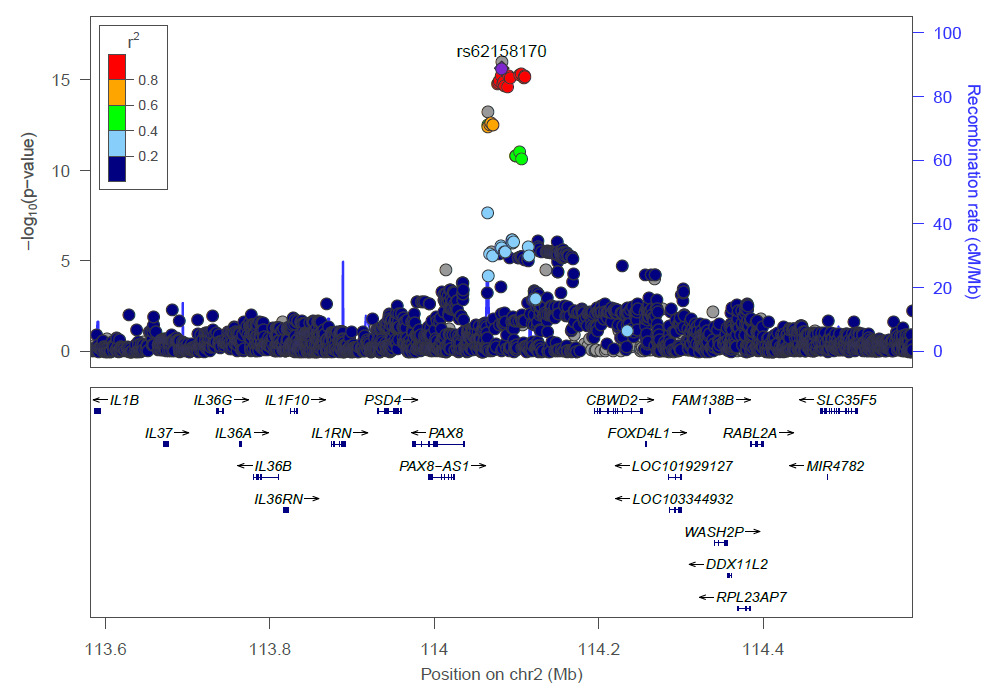

xi)

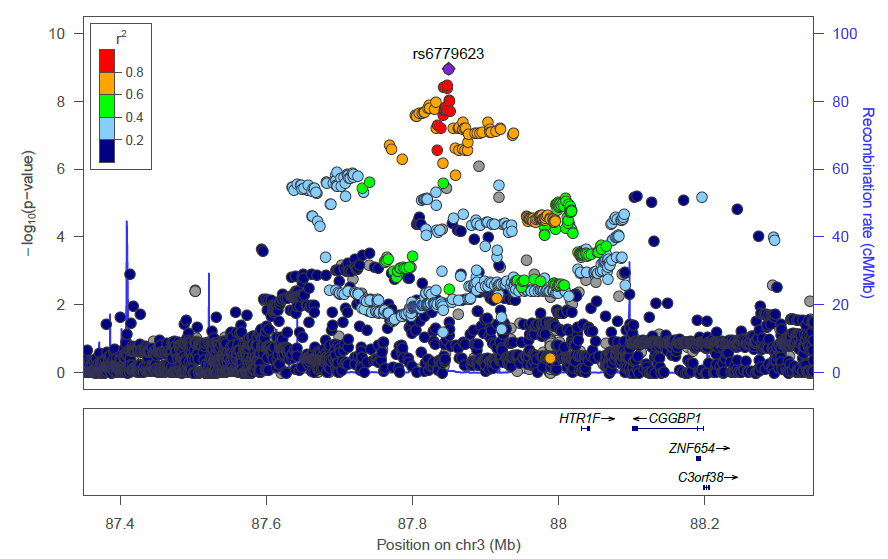

xii)

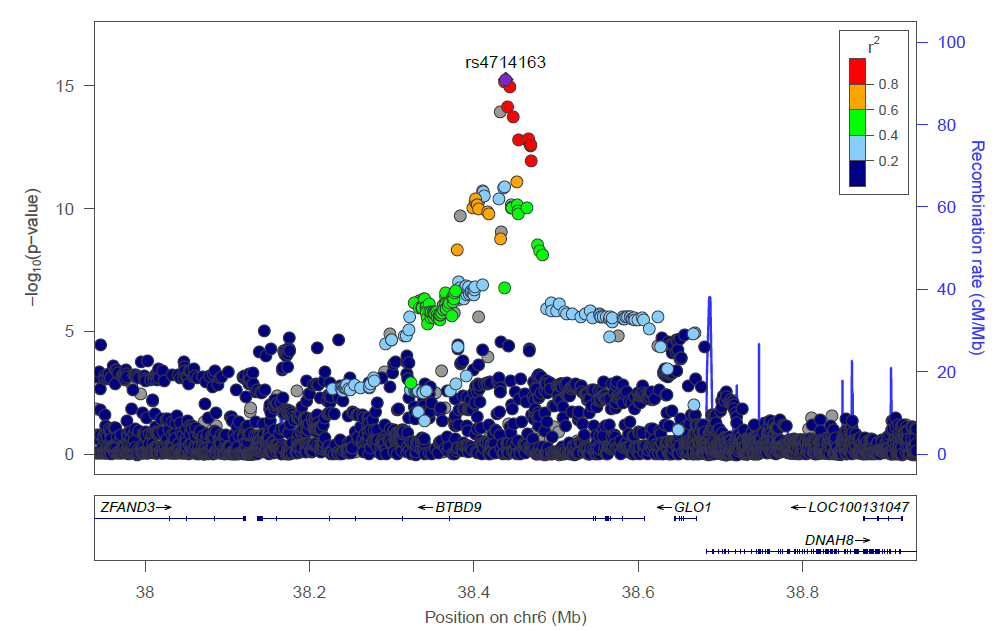

xiii)

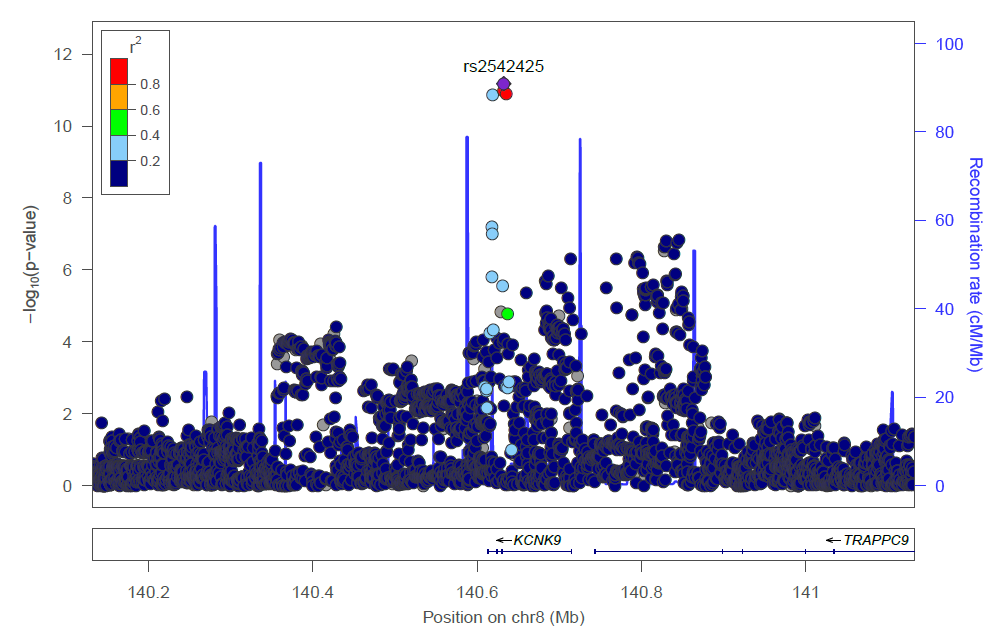

xiv)

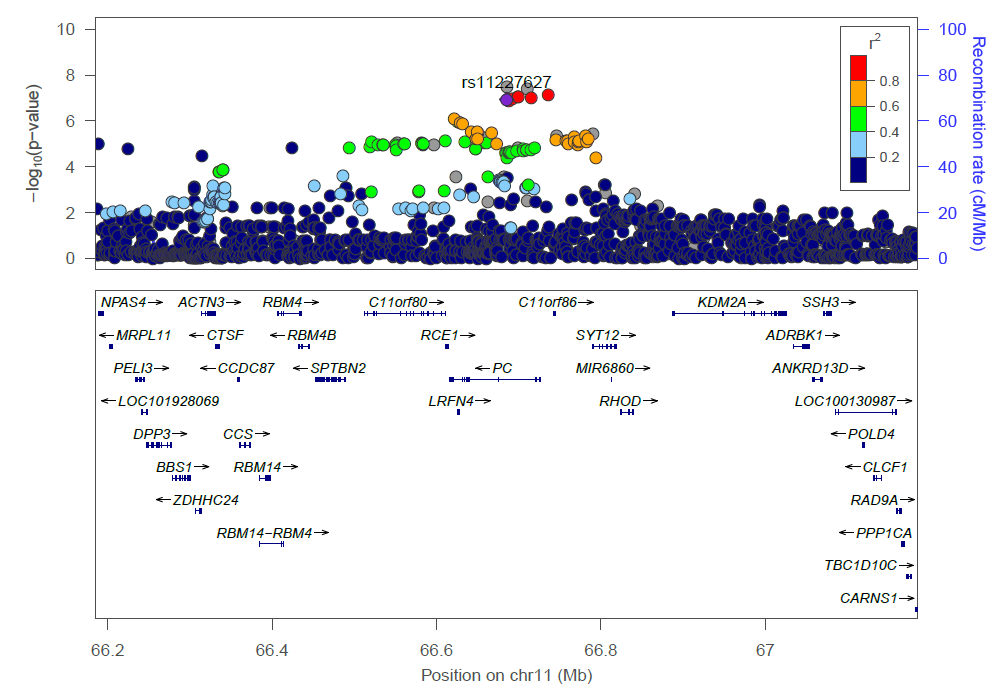

xv)

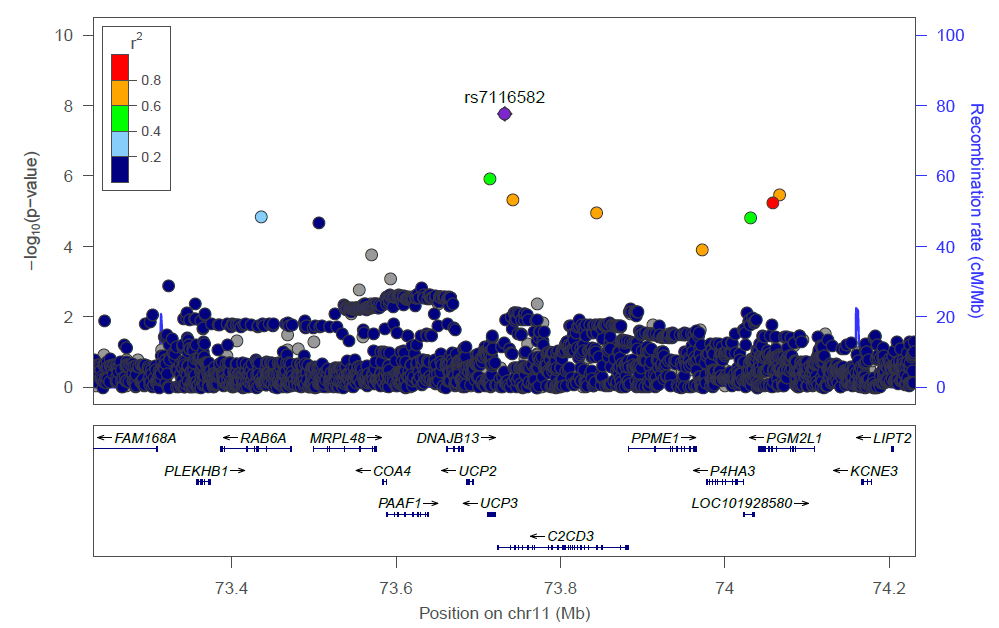

xvi)

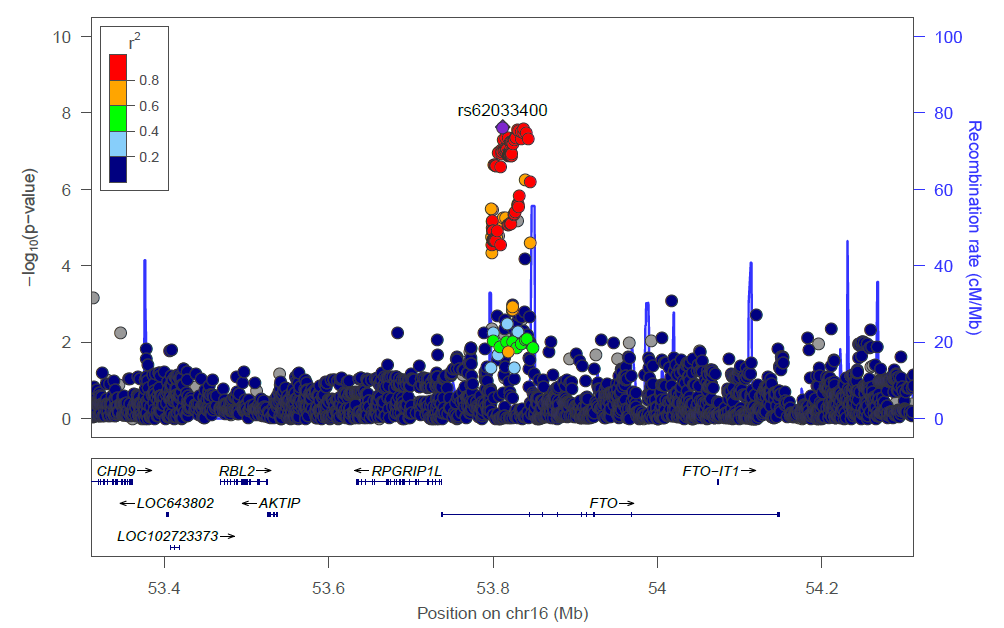

xvii)

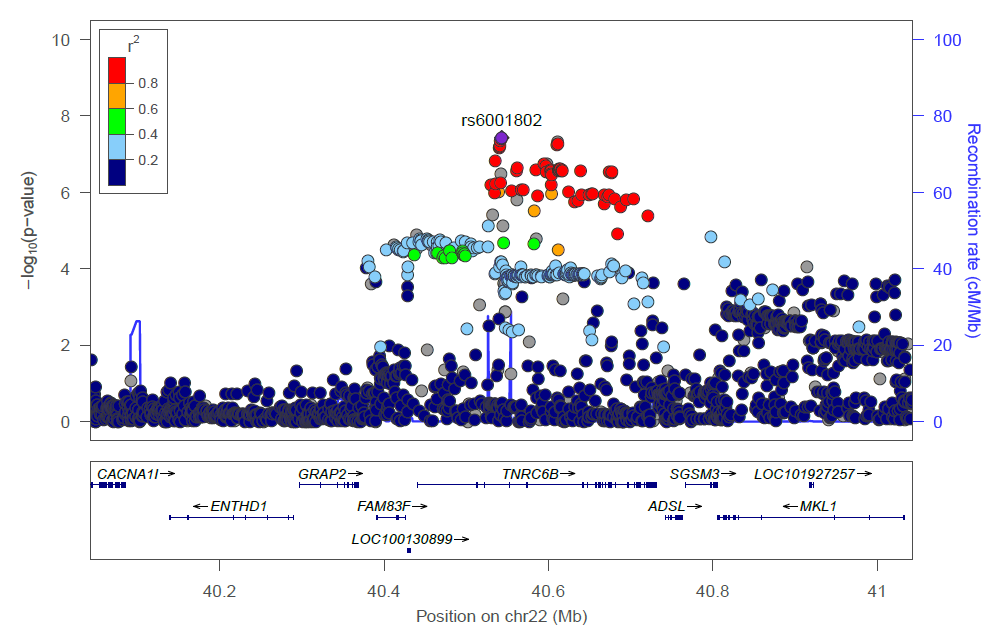

xviii)

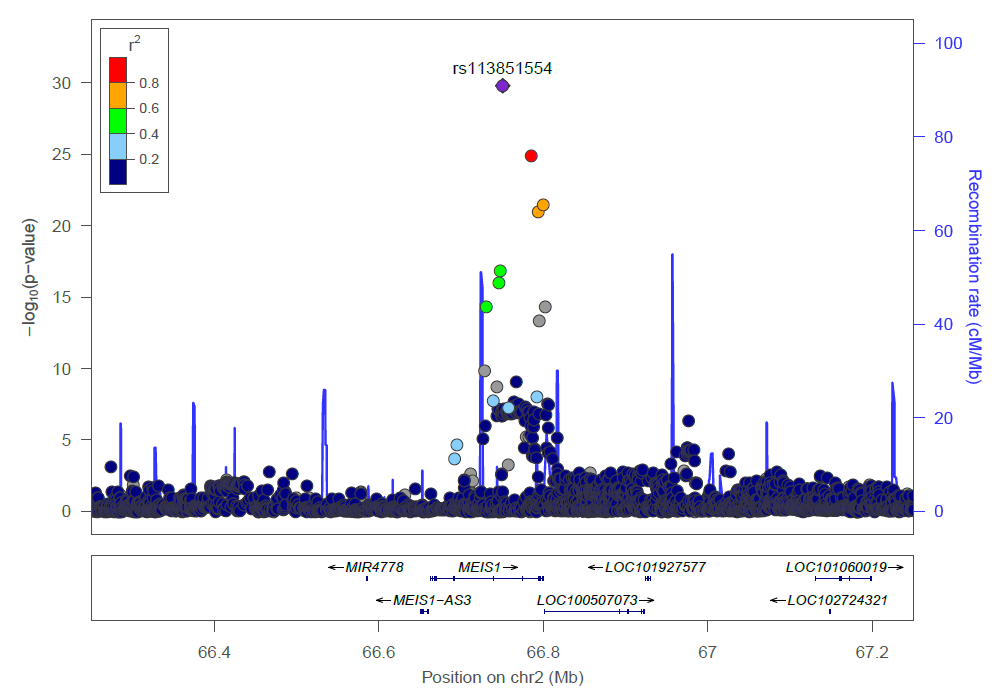

xix)

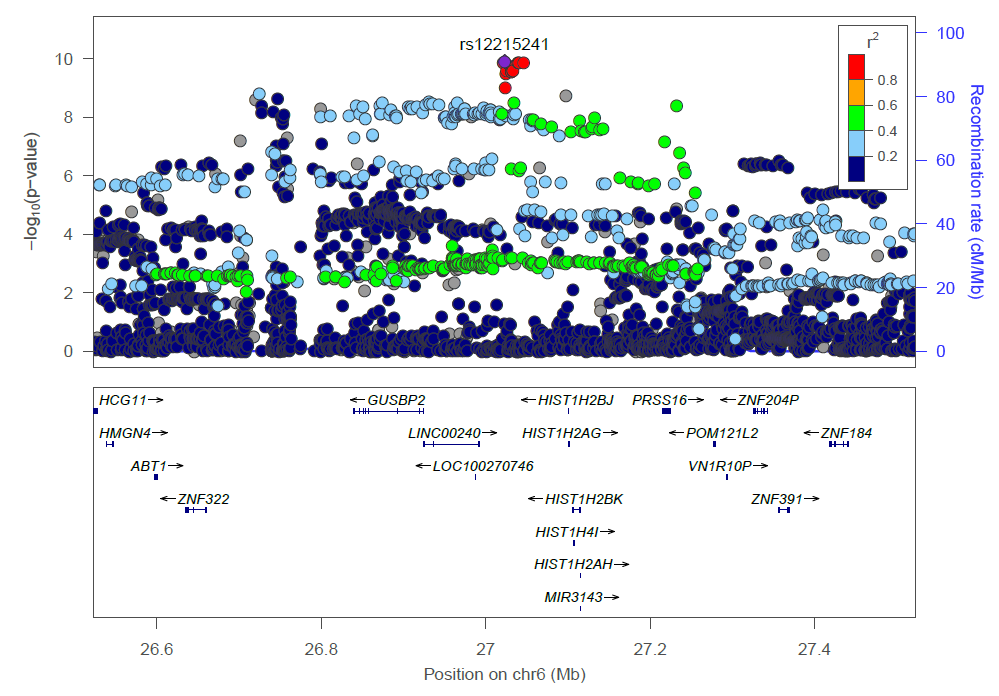

xx)

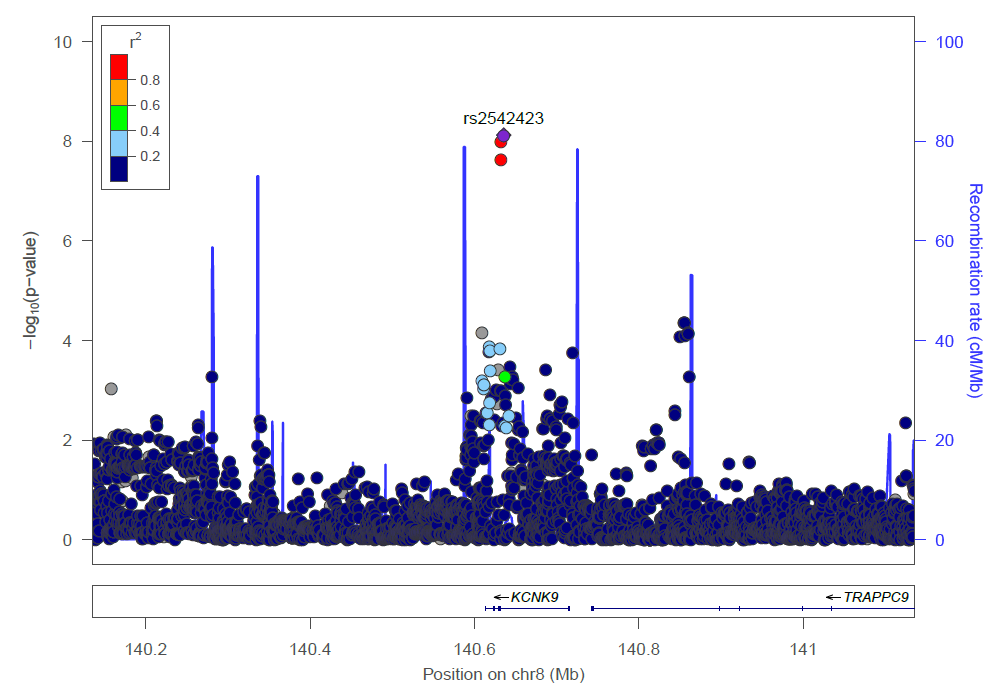

xxi)

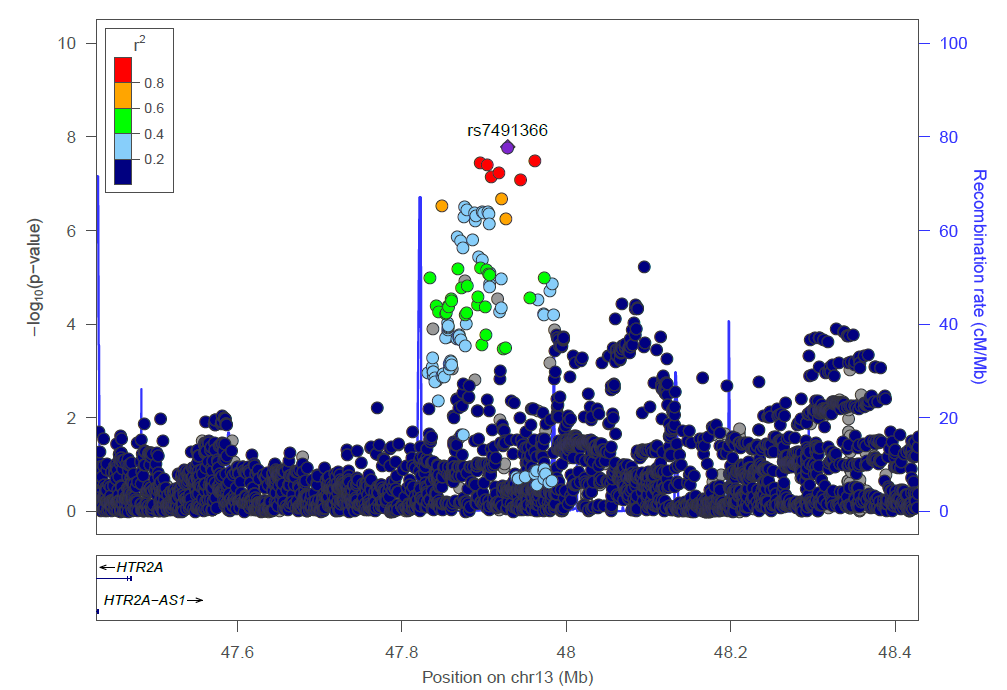

xxii)

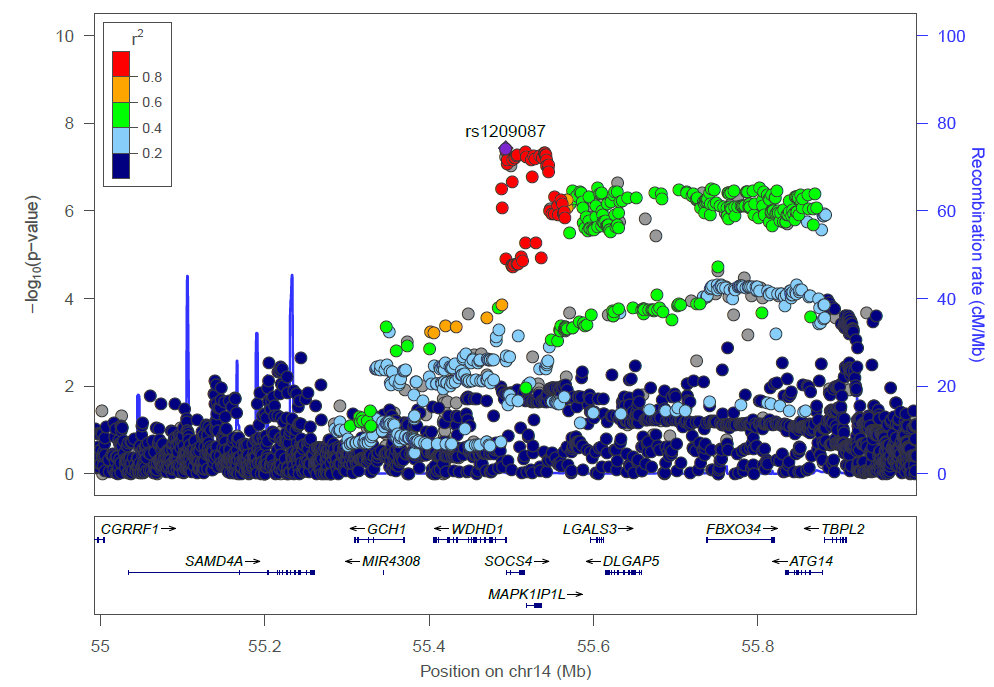

xxiii)

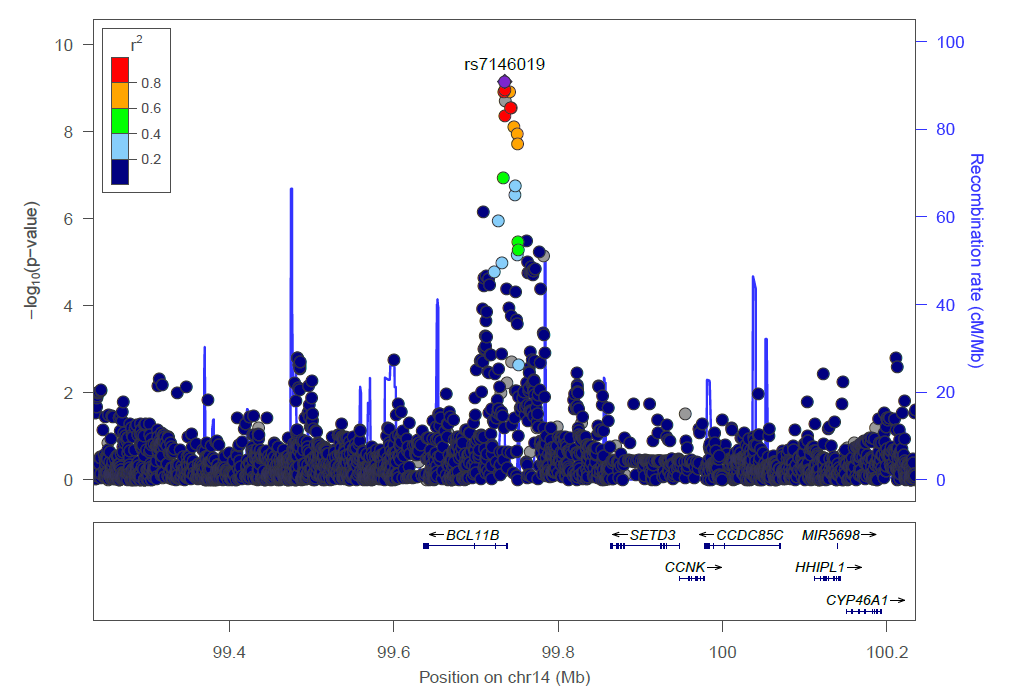

xxiv)

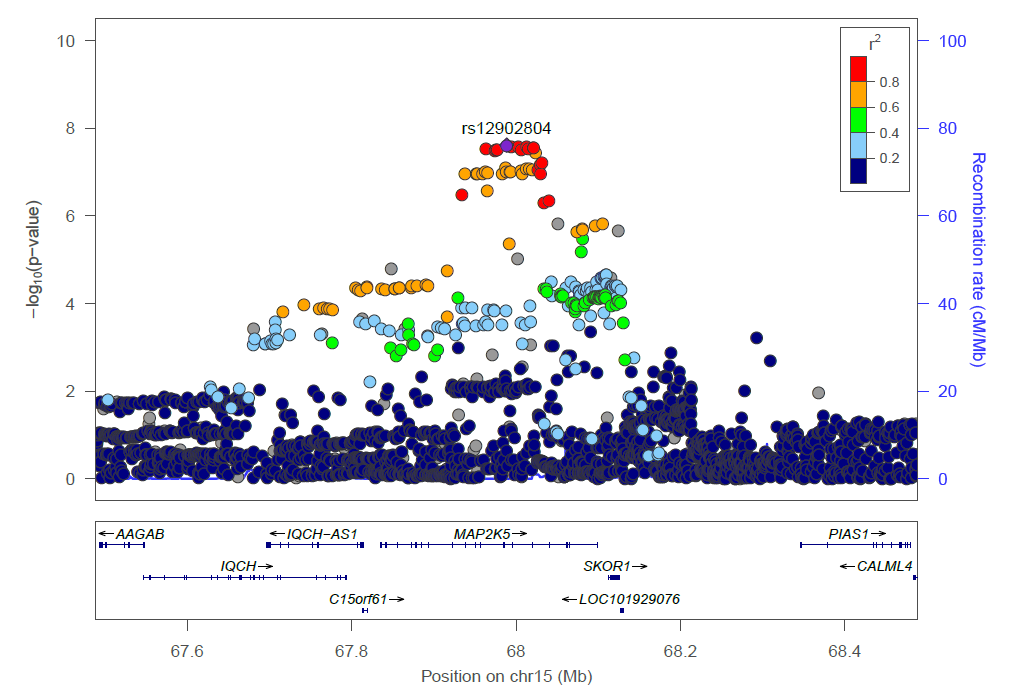

xxv)

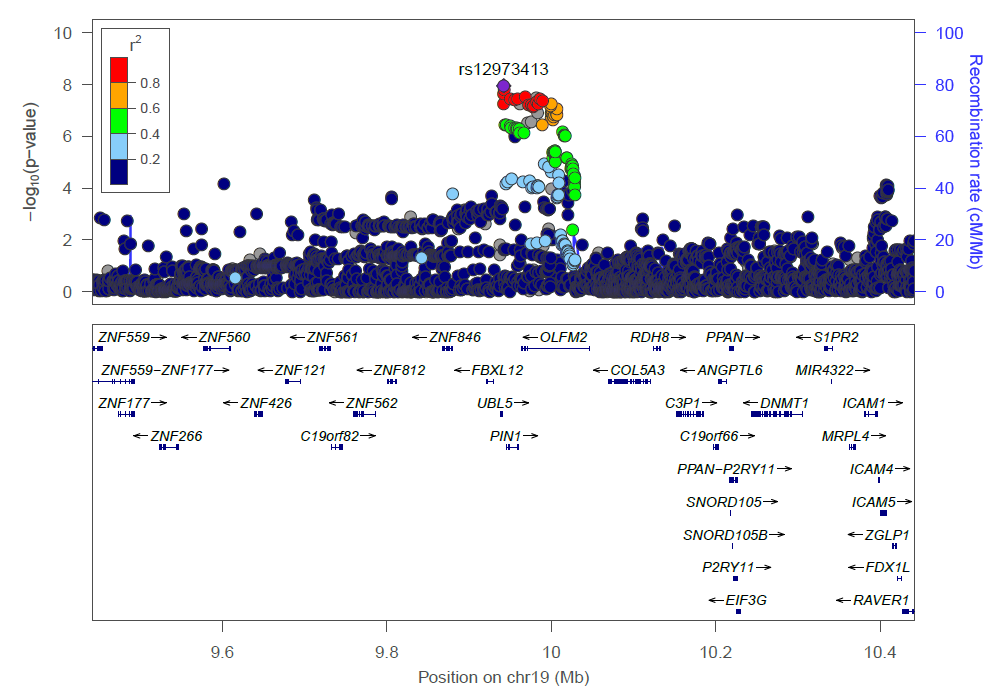

**Supplementary Figure 6**. Manhattan plot for the overall sample and by sex of sleep traits: a) night-time sleep duration, b) sleep efficiency, c) REM sleep duration, and d) Non-REM sleep duration.

a)

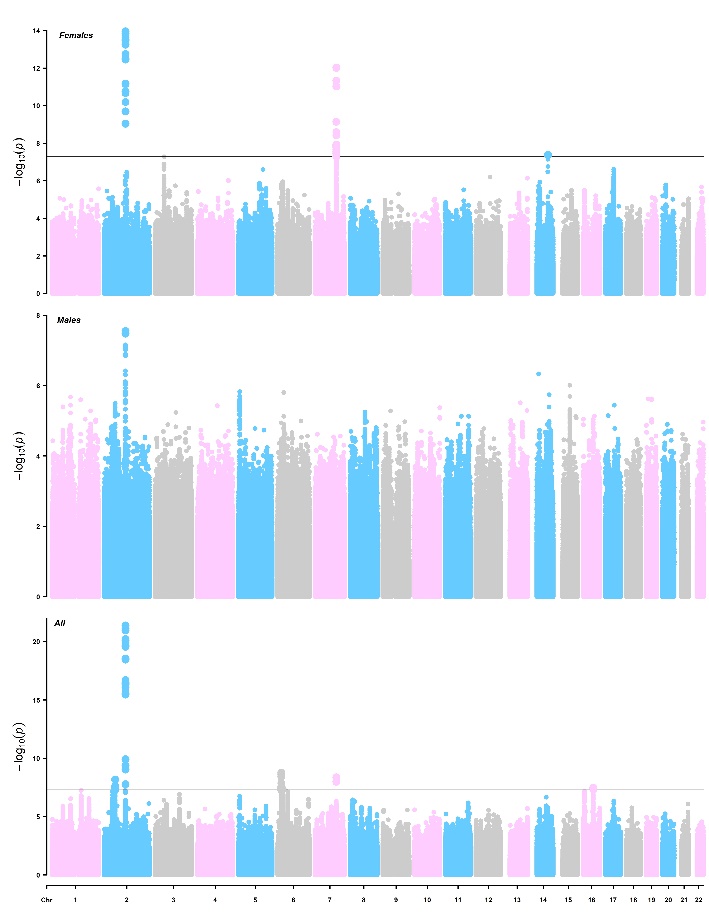

b)

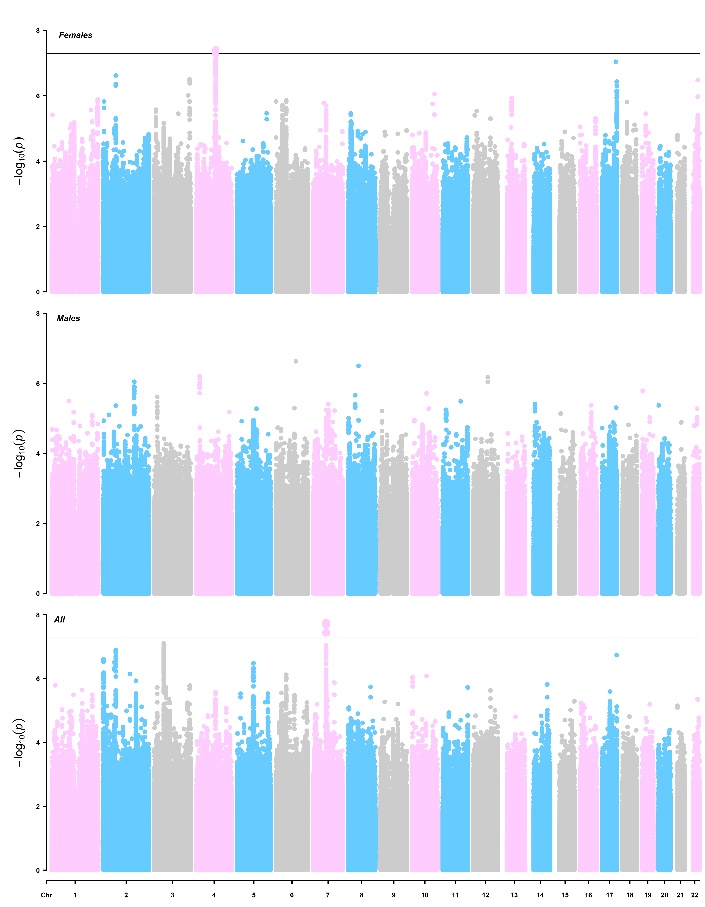

c)

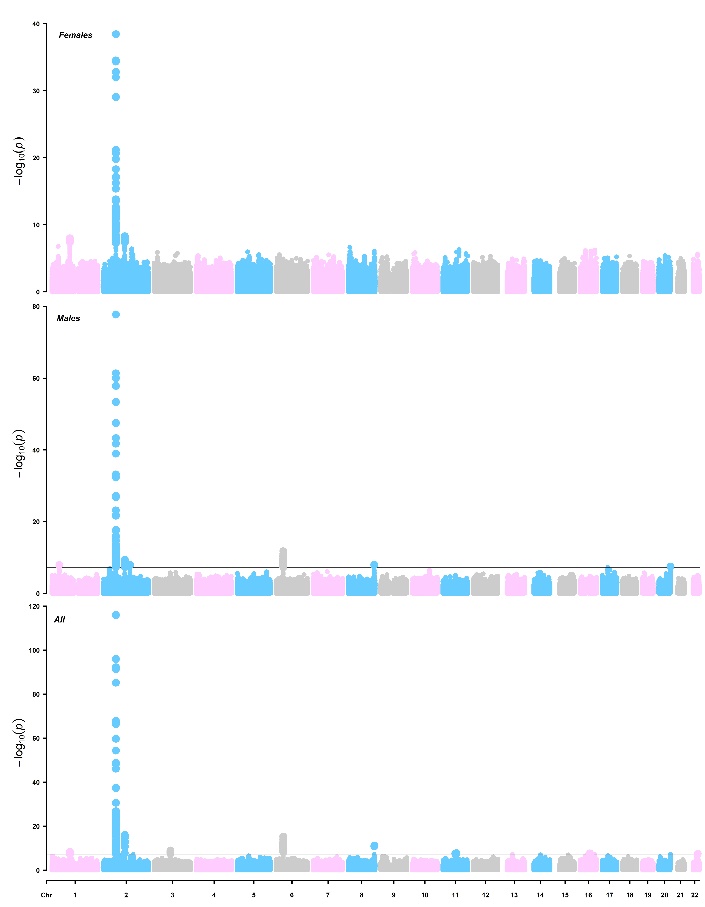

d)

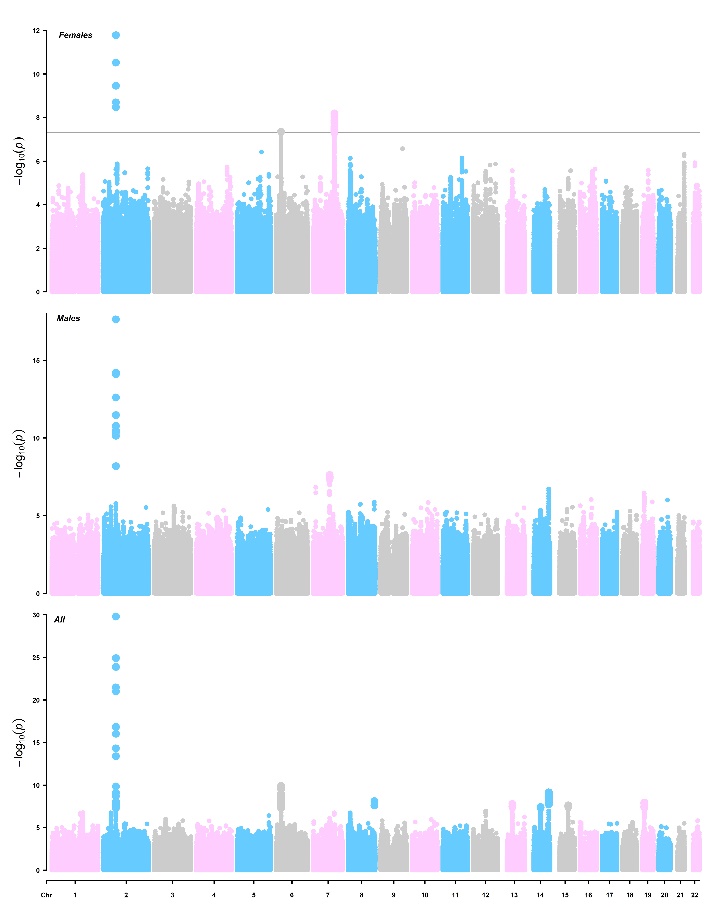

**Supplementary Figure 7**. Tissue enrichment of genes at associated loci for accelerometer-based sleep traits: a) night-time sleep duration; b) sleep efficiency; c) REM sleep; d) Non-REM sleep. Genes were identified using MAGMA and then analysed for enrichment using the GTEx RNA-seq database (v8) to investigate expression across 30 general tissues and 53 tissue types.

a)

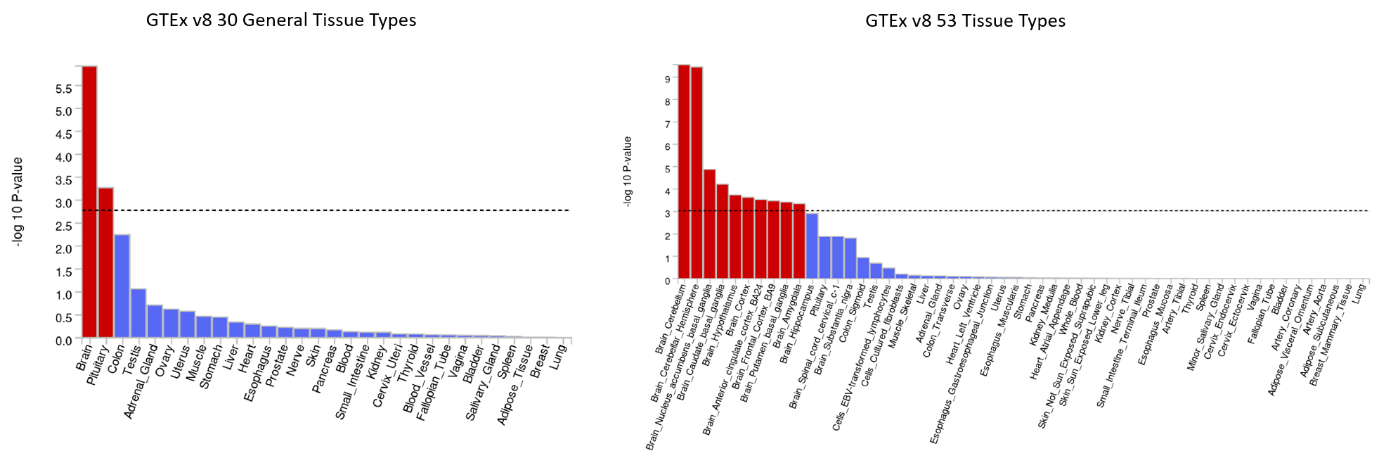

b)

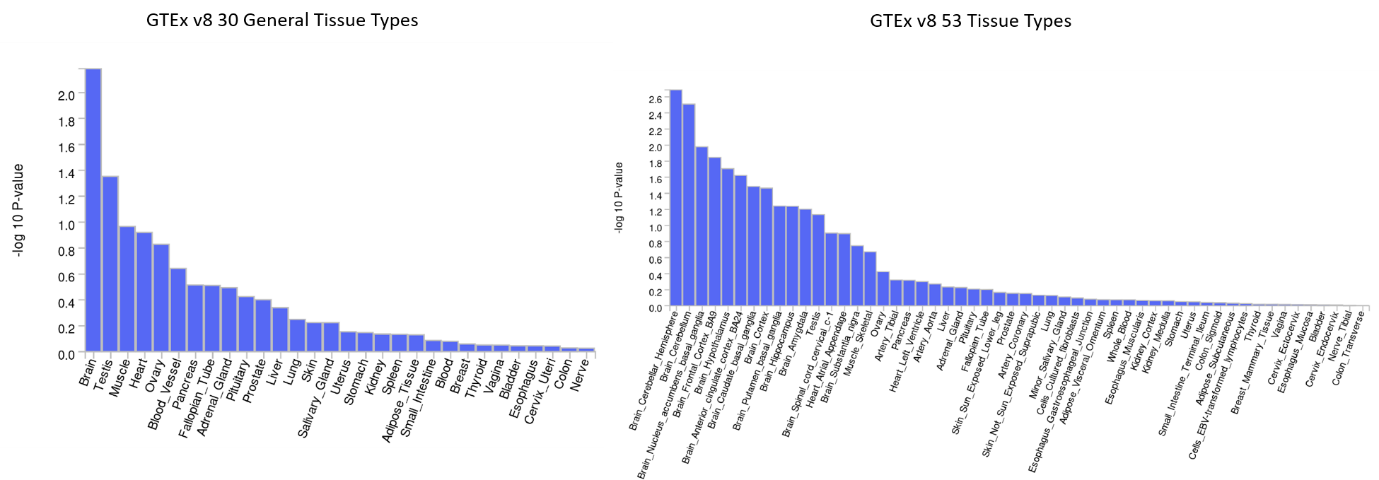

c)

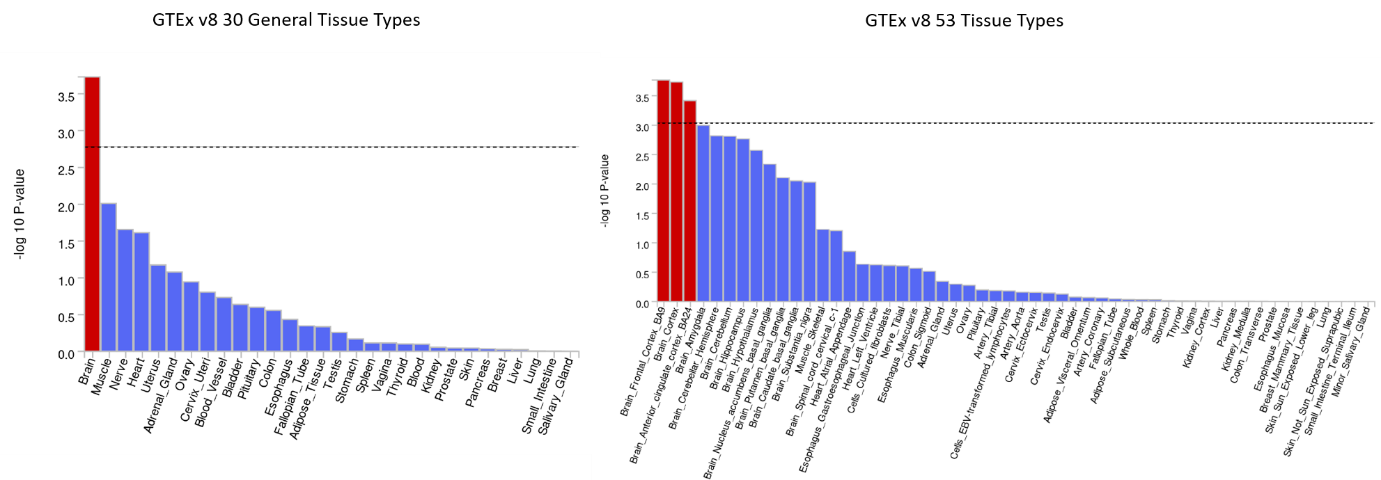

d)

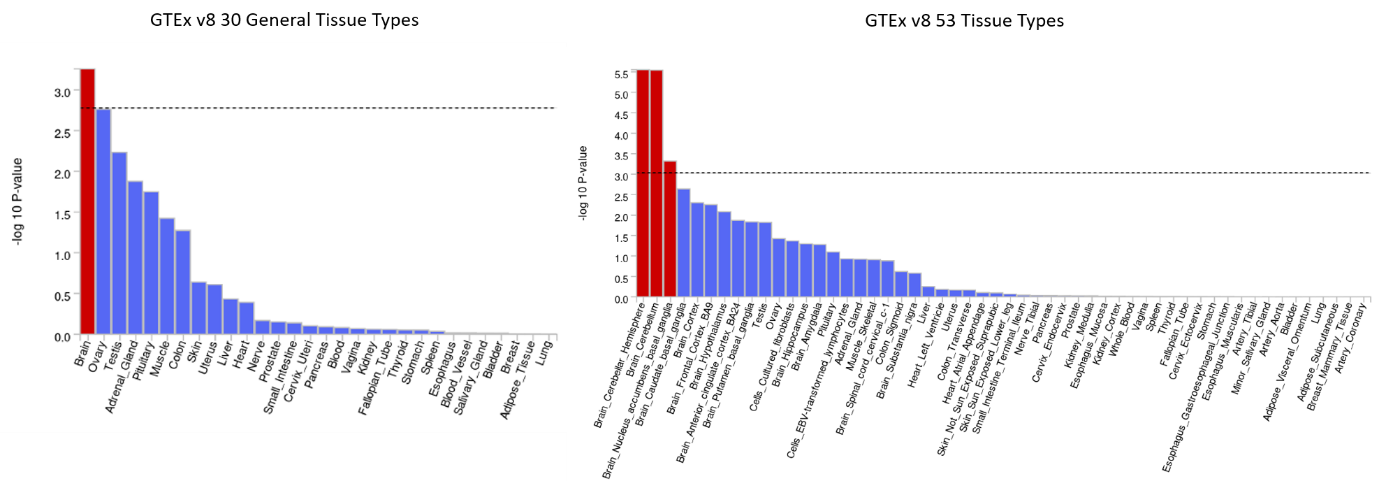

**Supplementary Figure 8**. Circular plots of functional mapping for accelerometer-based sleep traits: a) night-time sleep duration; b) sleep efficiency; c) REM sleep; d) Non-REM sleep. Genomic risk loci are highlighted in blue. Genes mapped only by chromatin interactions in orange, only by eQTLs in green, by both in red. Links colored orange are chromatin interactions, links colored green are eQTLs.

a)

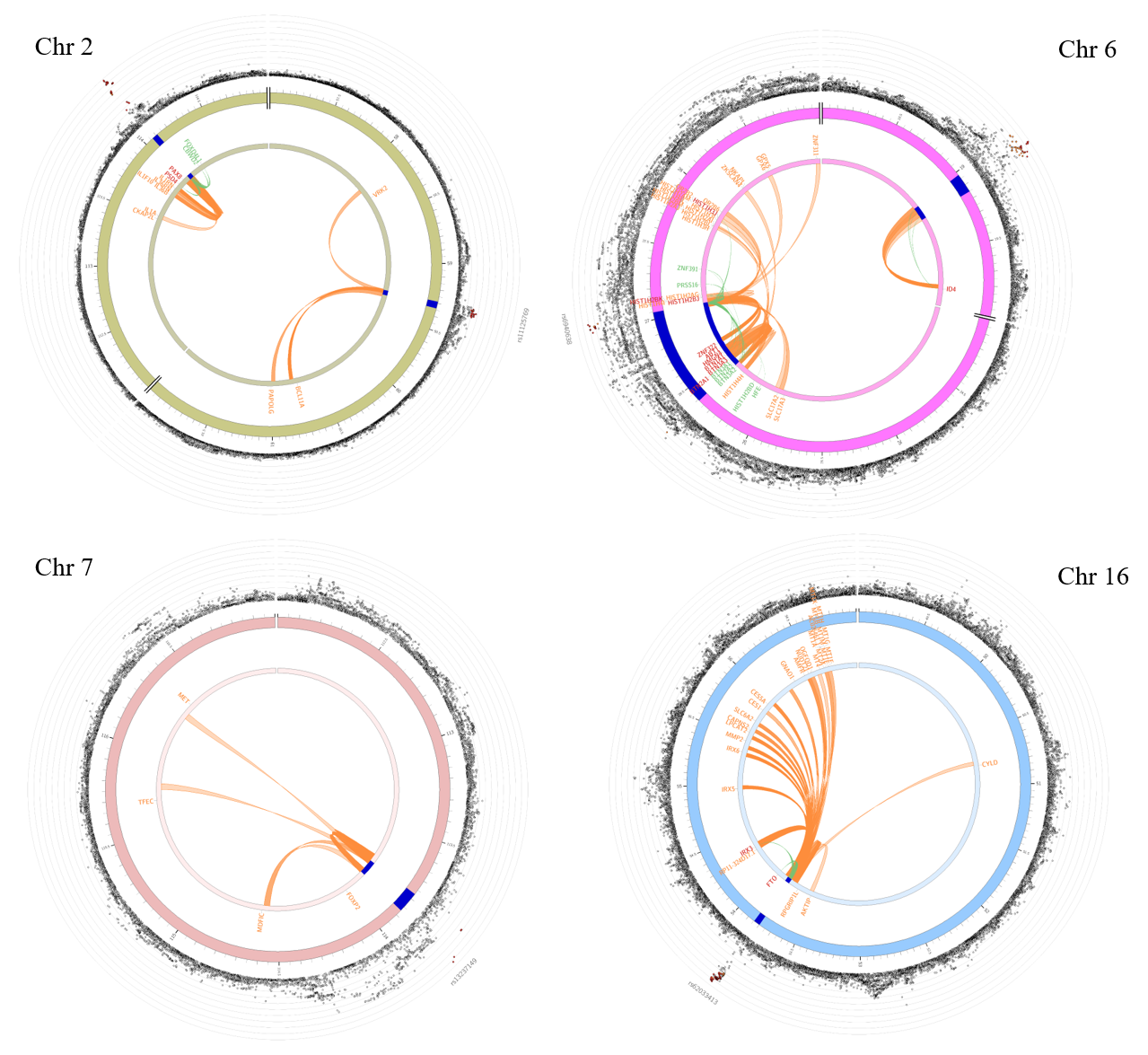

b)

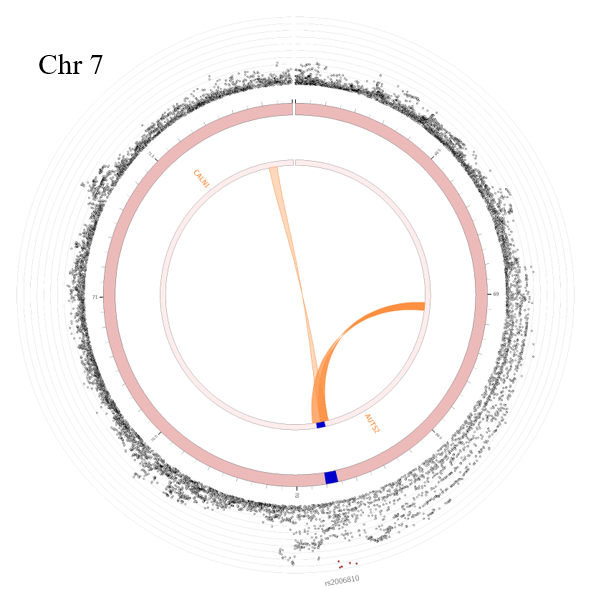

c)

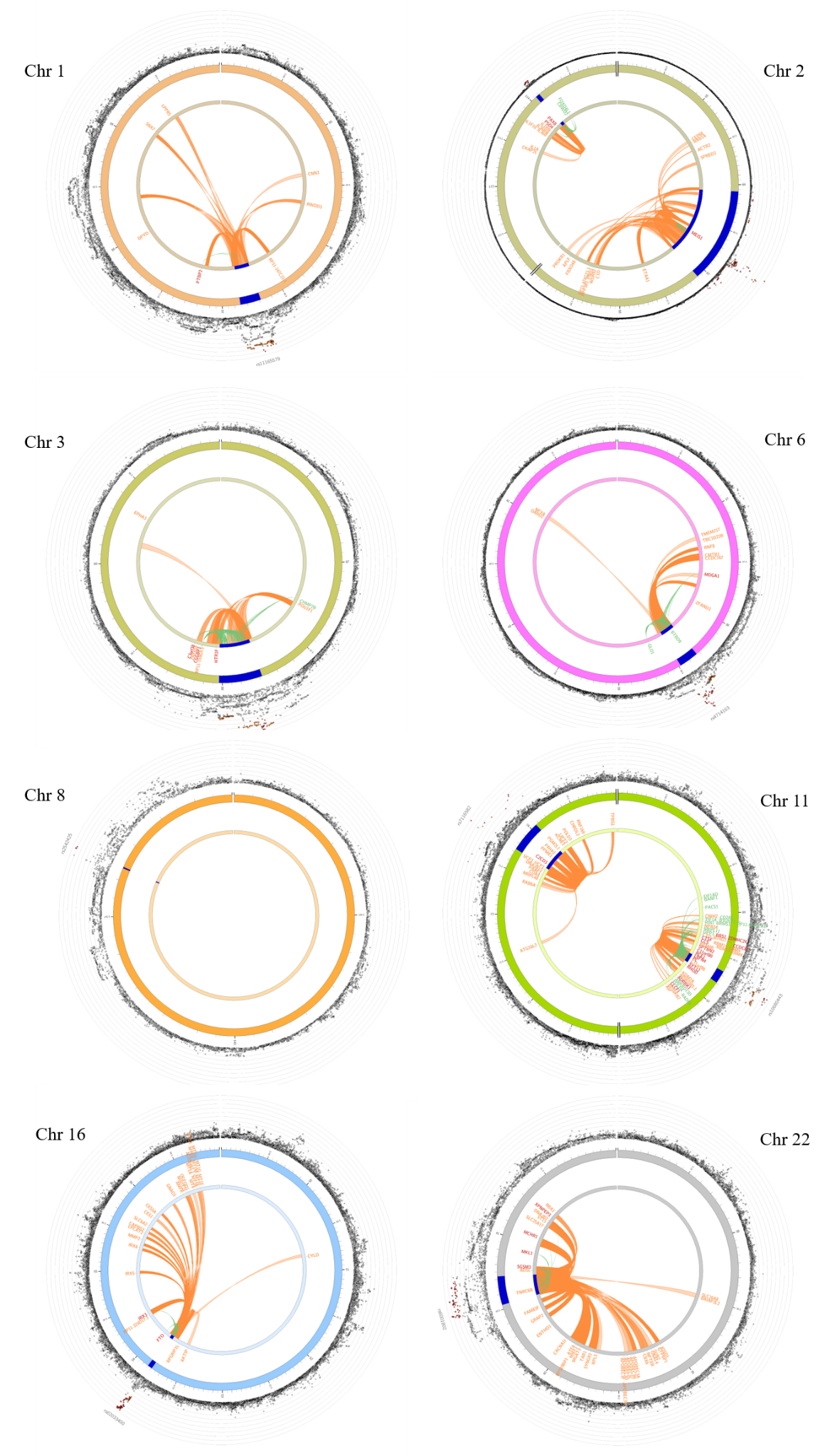

d)

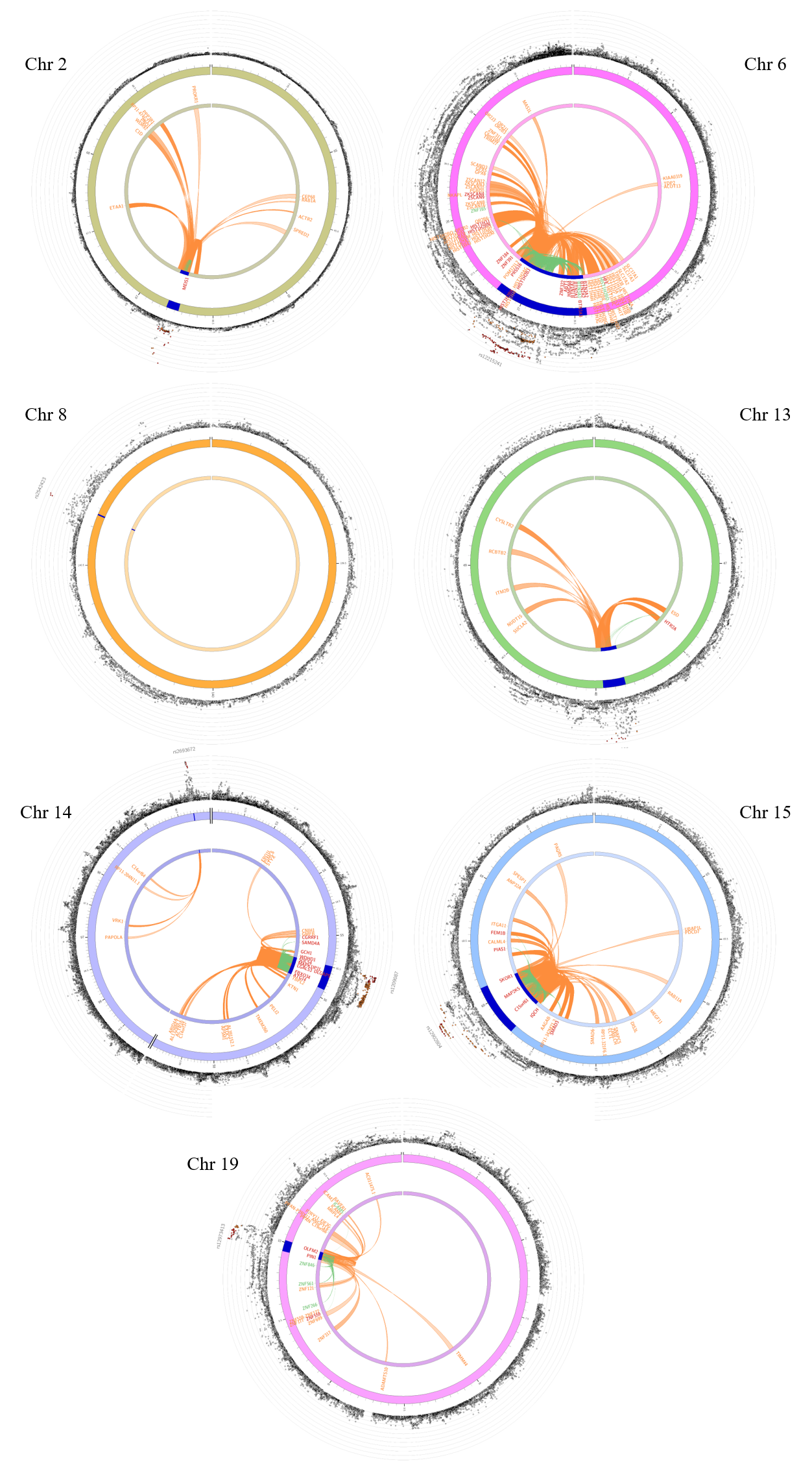

**Supplementary Figure 9**. LocusCompare plots of significant GWAS-eQTL colocalizations with a p-value ≤ 5 x 10^-8^ in both GWAS and eQTL data for sleep traits (night-time sleep: i) at the 2q14.1 locus for FOXD4L1 in the thyroid, ii) at the 2q14.1 locus for CBWD2 in the thyroid; REM sleep: iii) at the 2q14.1 locus for FOXD4L1 in the thyroid, iv) at the 2q14.1 locus for CBWD2 in the thyroid, v) at the 3p11.1 locus for HTR1F in the adipose visceral omentum, vi) at the 3p11.1 locus for HTR1F in the heart left ventricle, vii) at the 3p11.1 locus for HTR1F in the spleen, viii) at the 3p11.1 locus for HTR1F in the thyroid; Non-REM sleep: ix) at the 6p22.2 locus for BTN2A2 in the pancreas, x) at the 6p22.2 locus for HIST1H2BK in the tibial nerve, xi) at the 14q22.3 locus for WDHD1 in the esophagus mucosa, xii) at the 15q23 locus for SKOR1 in the thyroid, xiii) at the 19p13.2 locus for PIN1DT in the brain frontal cortex). Left hand panel represents GWAS (x-axis) and eQTL (y-axis) -log p values for the gene variants at the locus. Right hand panels represent association data with the -log p values on the y-axis for the eQTL (upper panel) and GWAS (lower panel).

i)

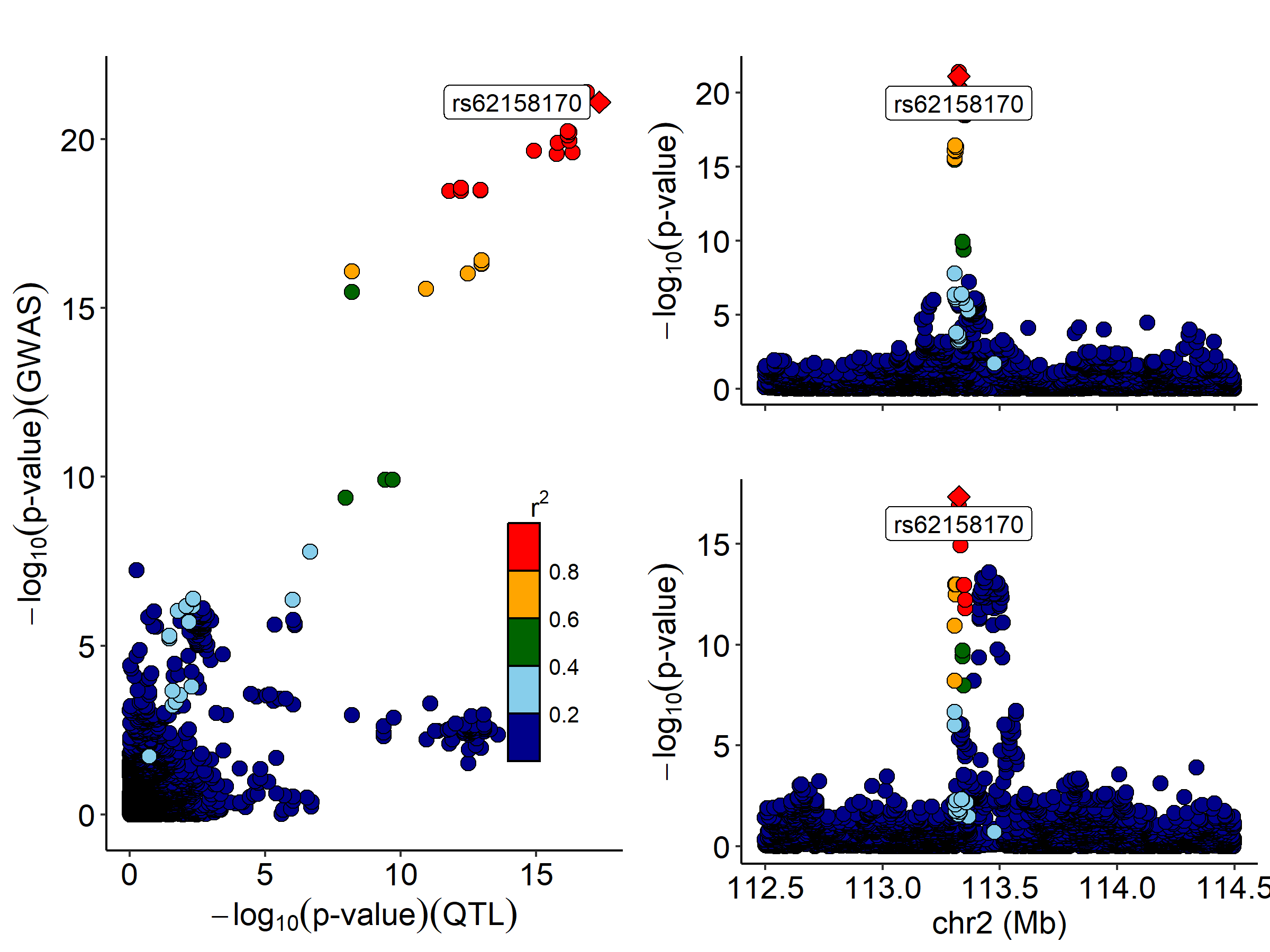

ii)

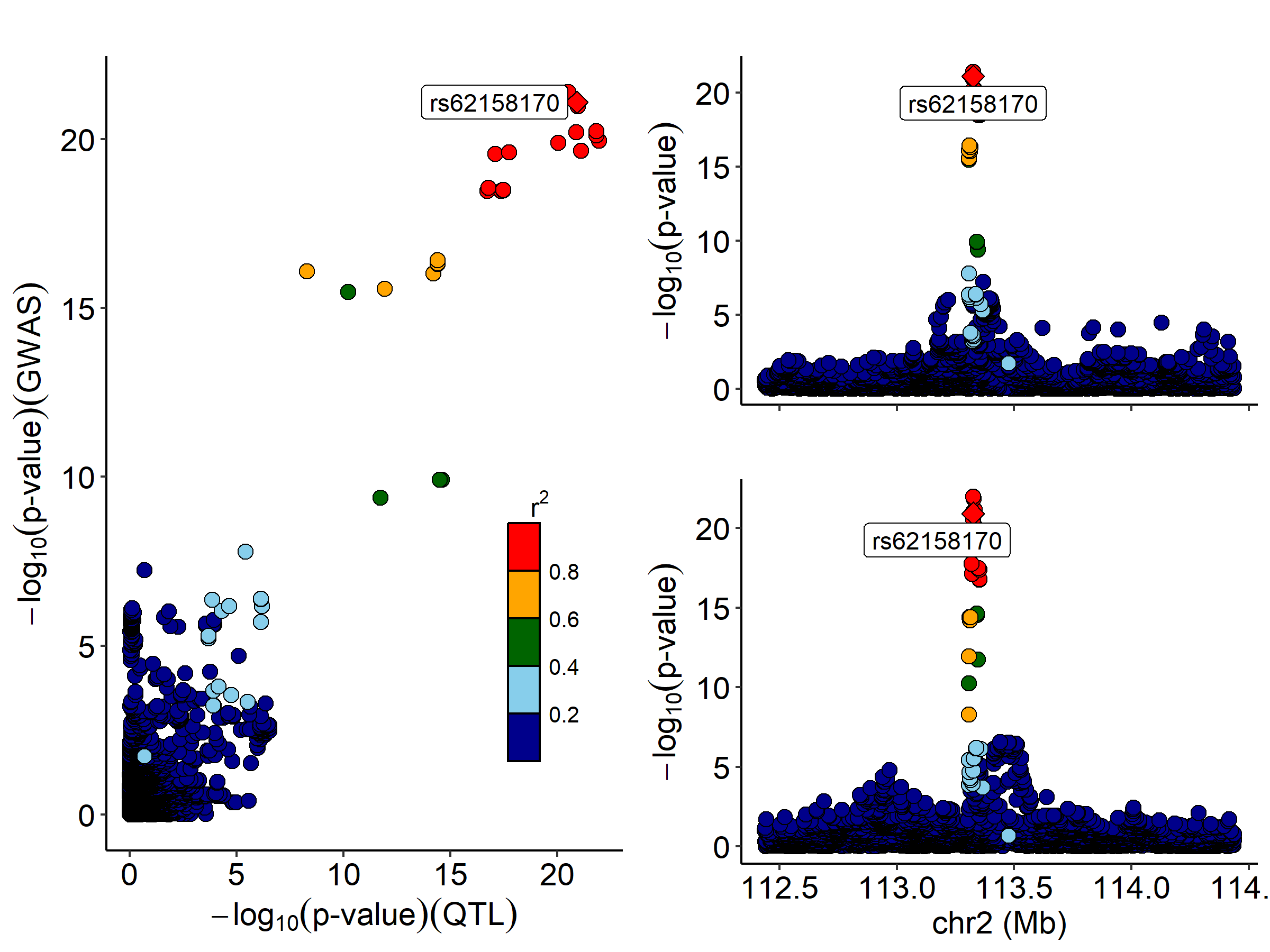

iii)

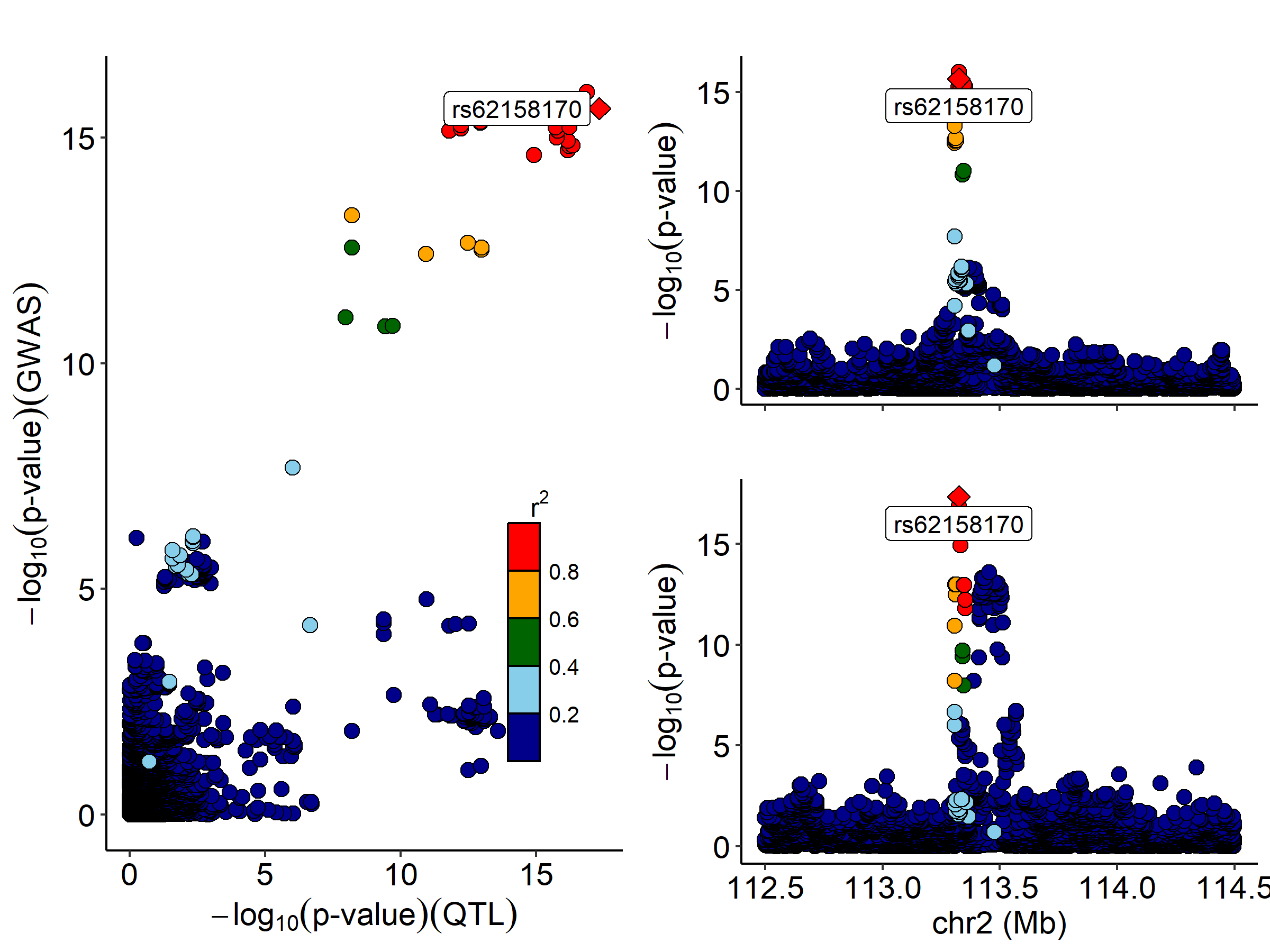

iv)

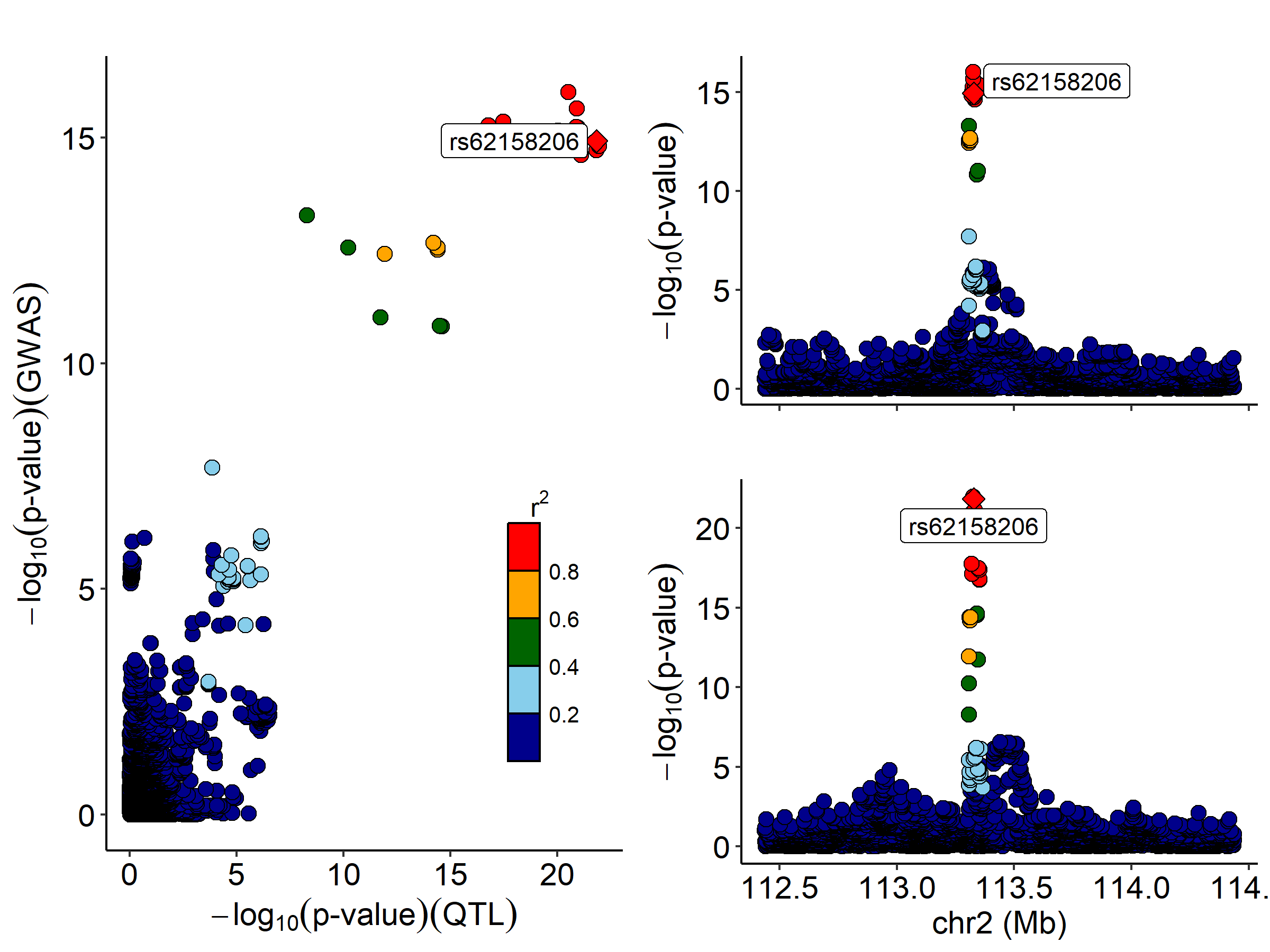

v)

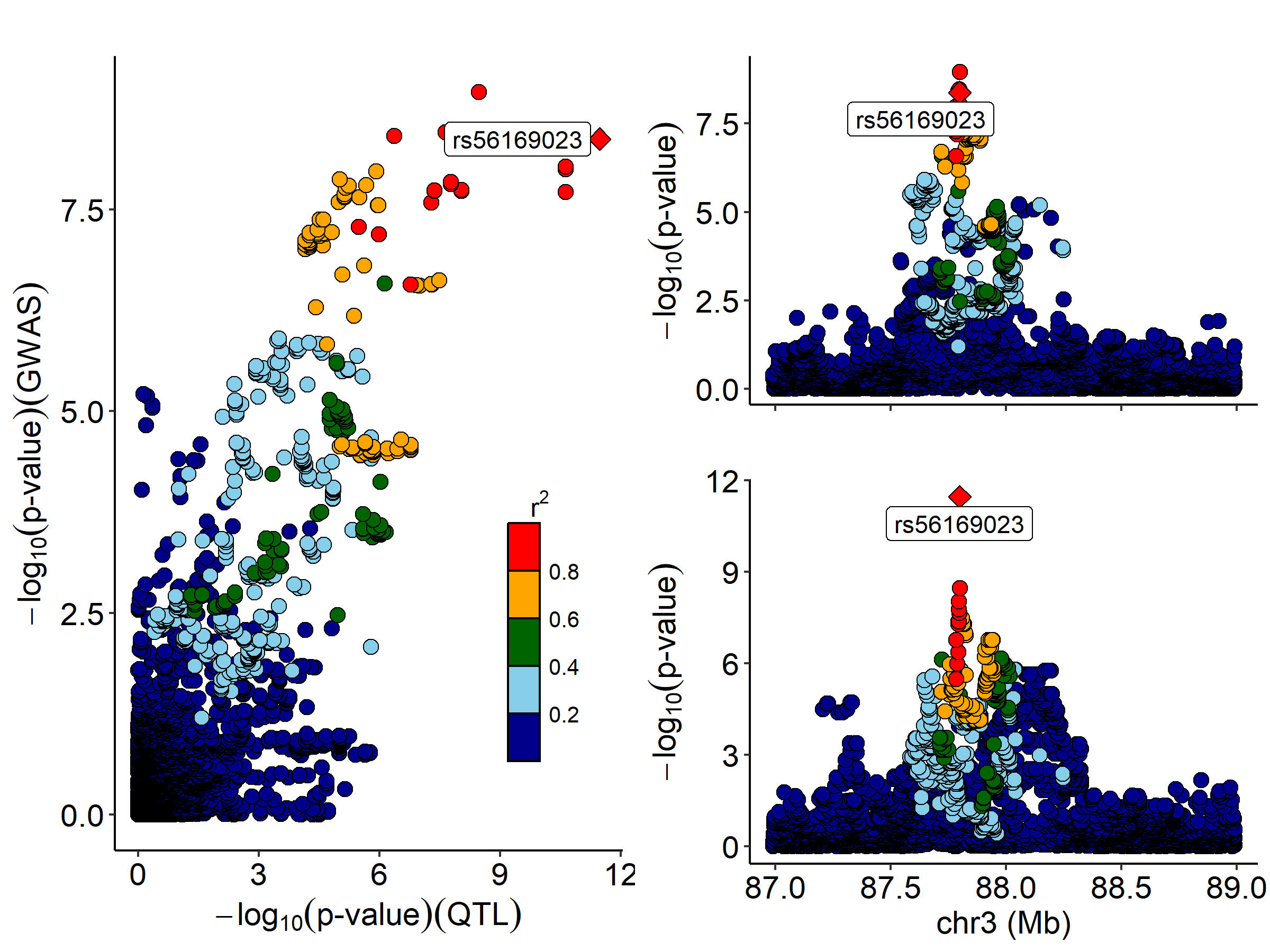

vi)

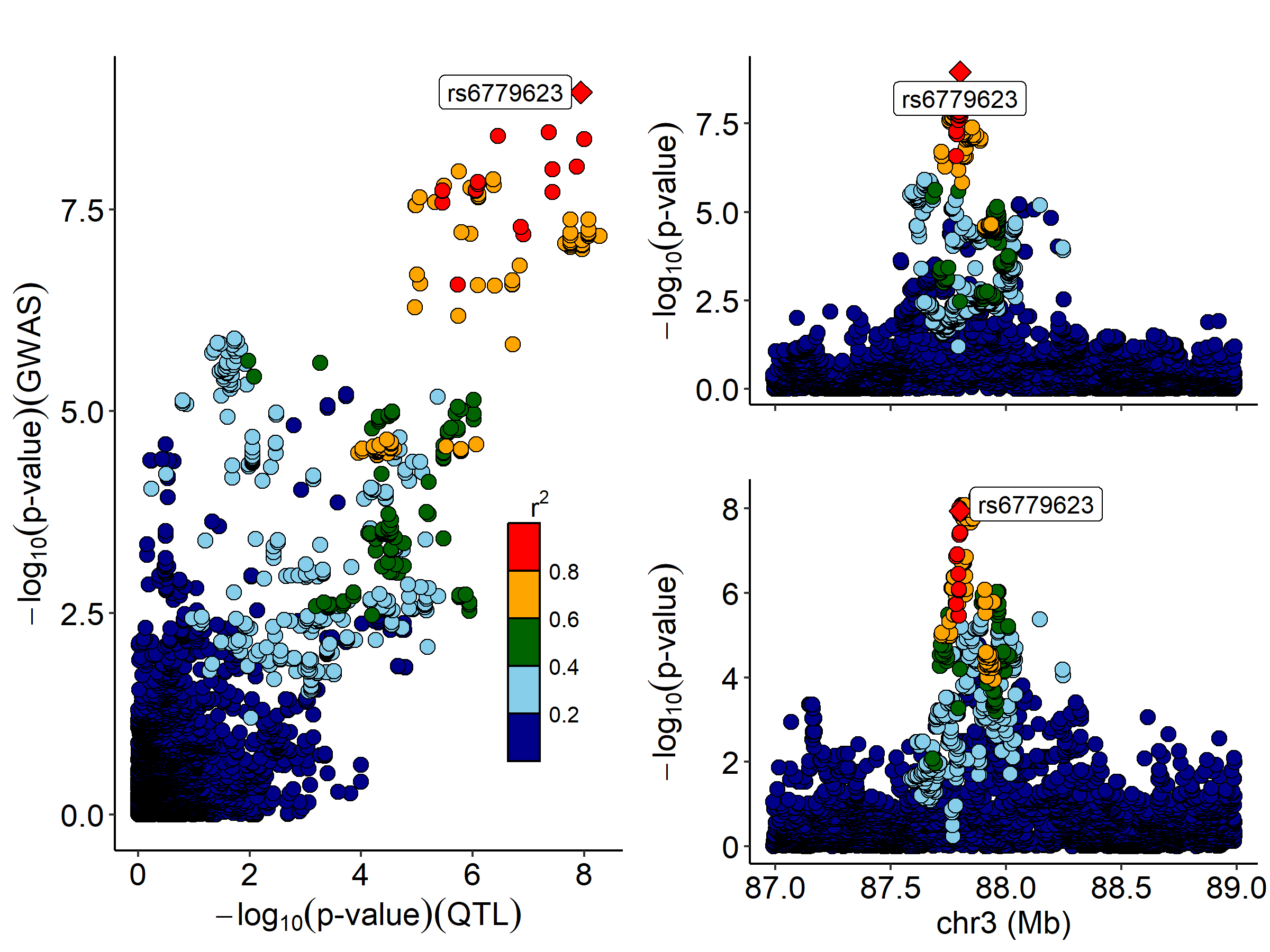

vii)

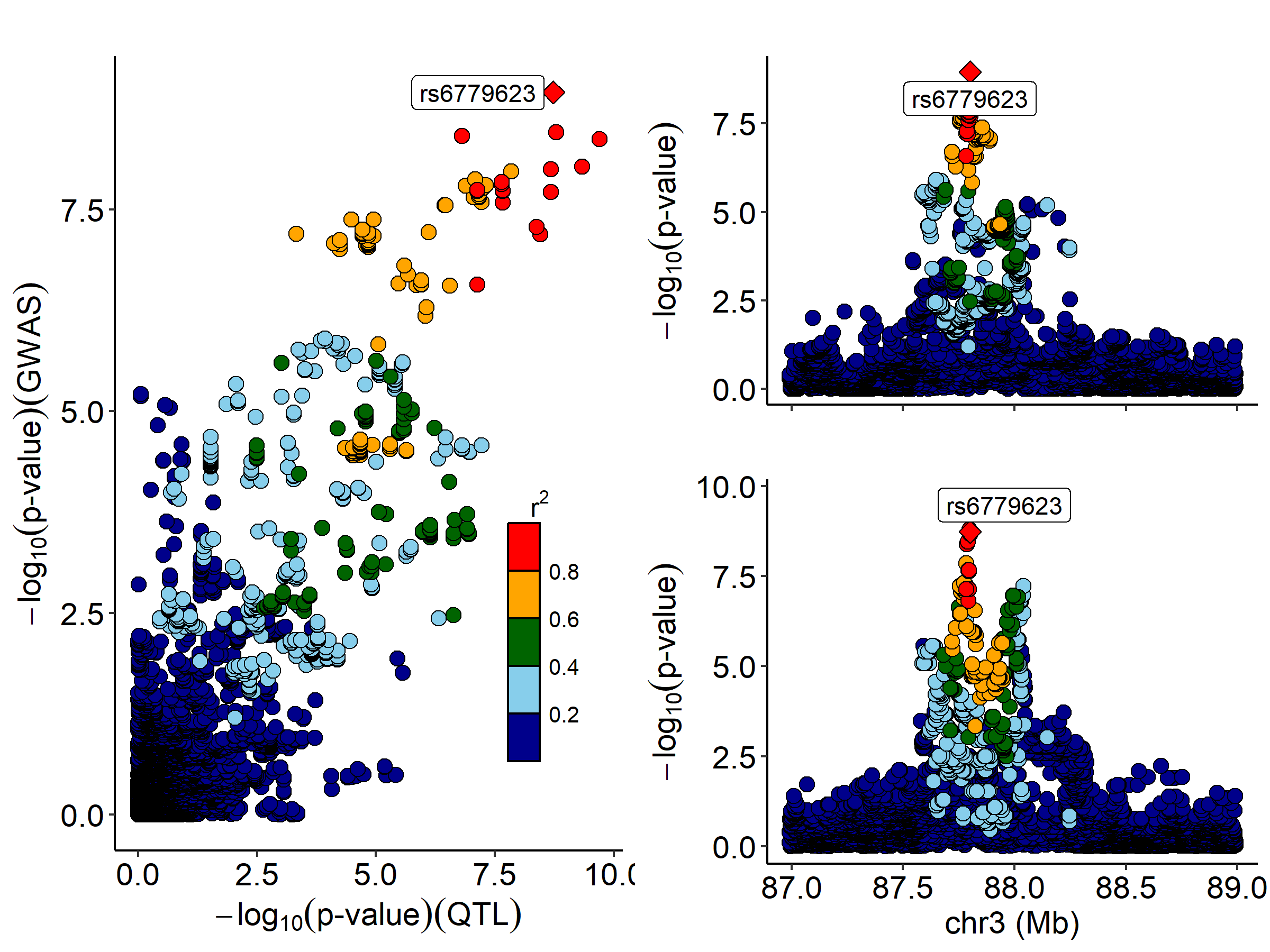

viii)

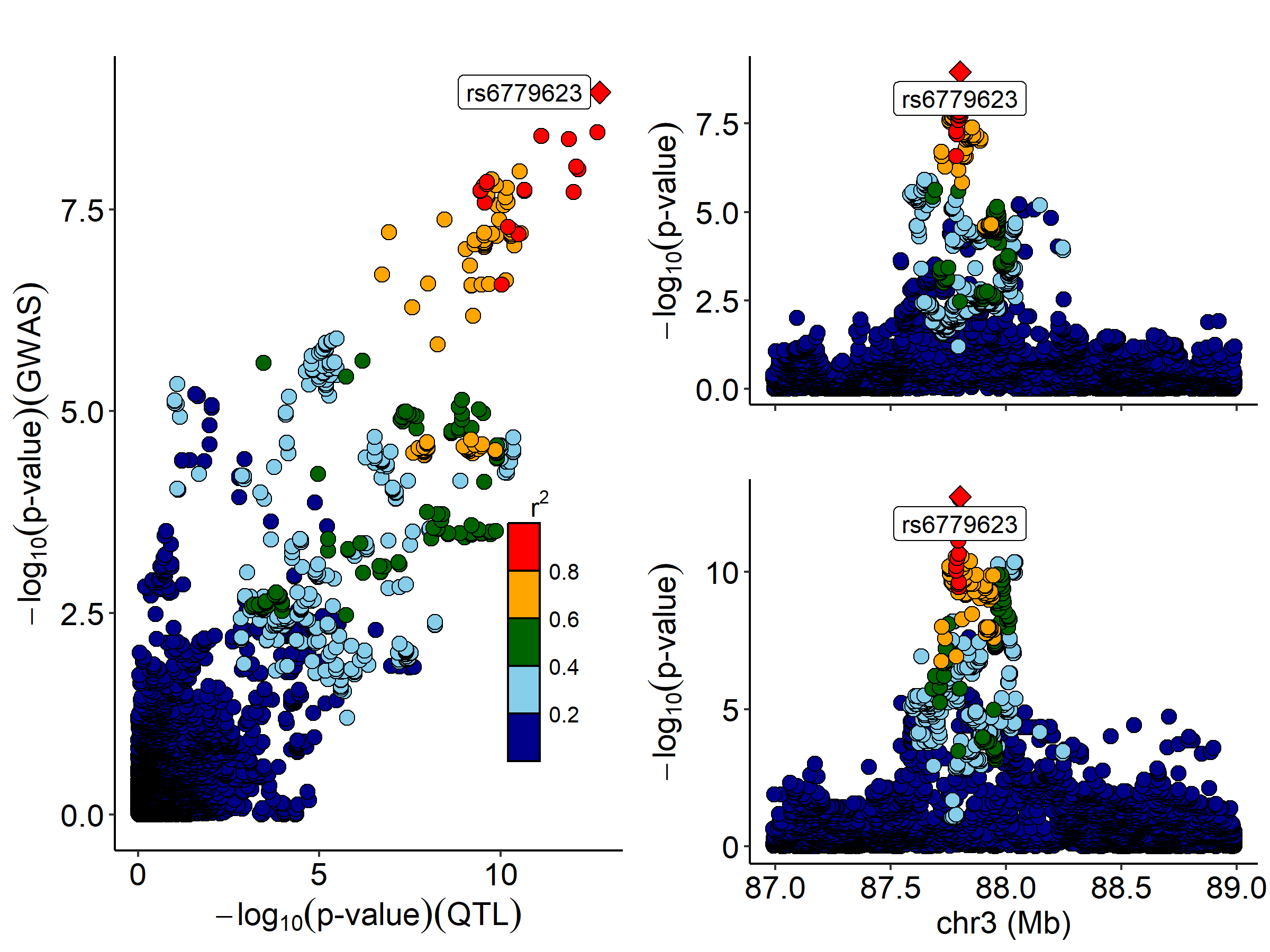

ix)

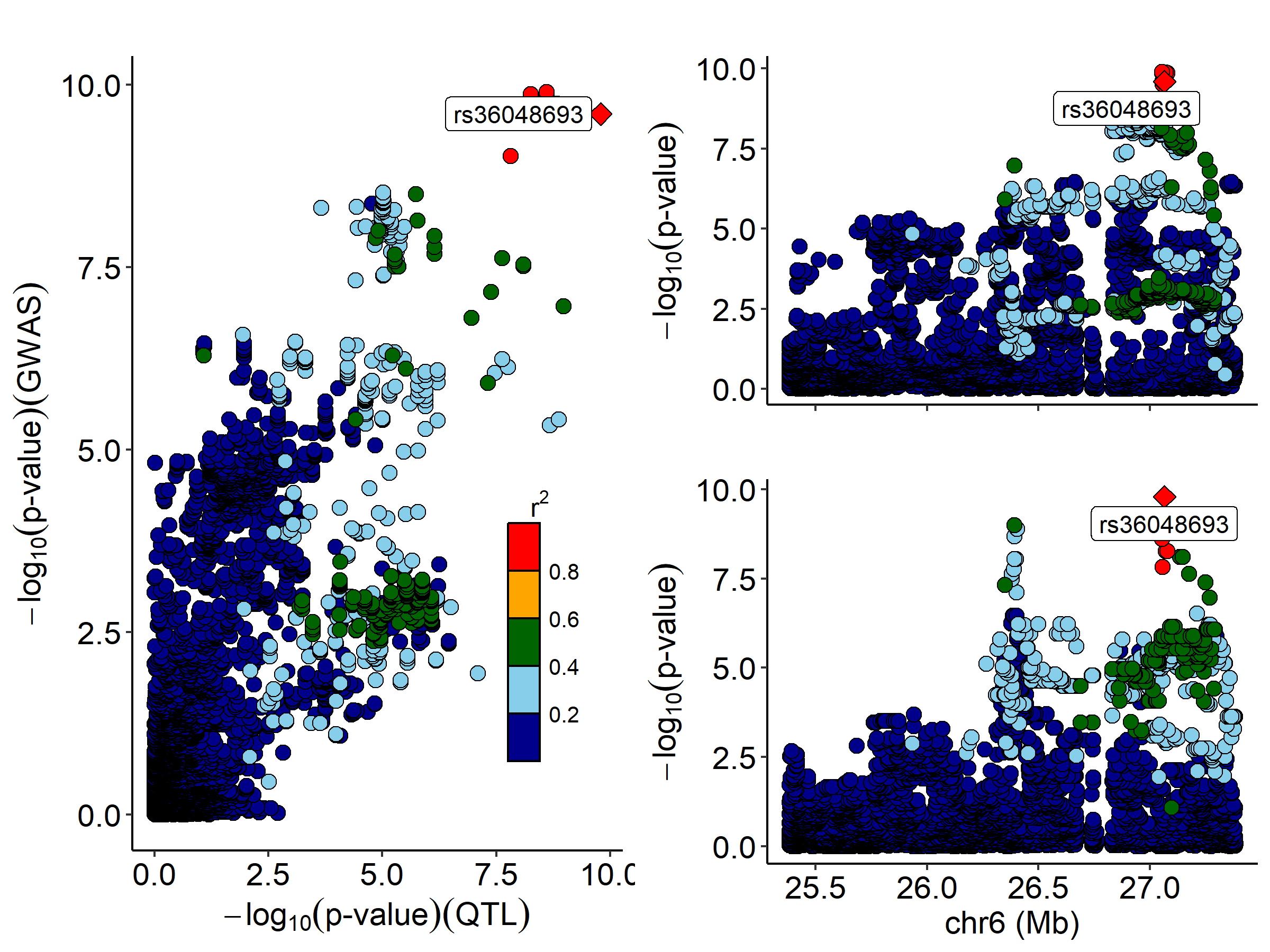

x)

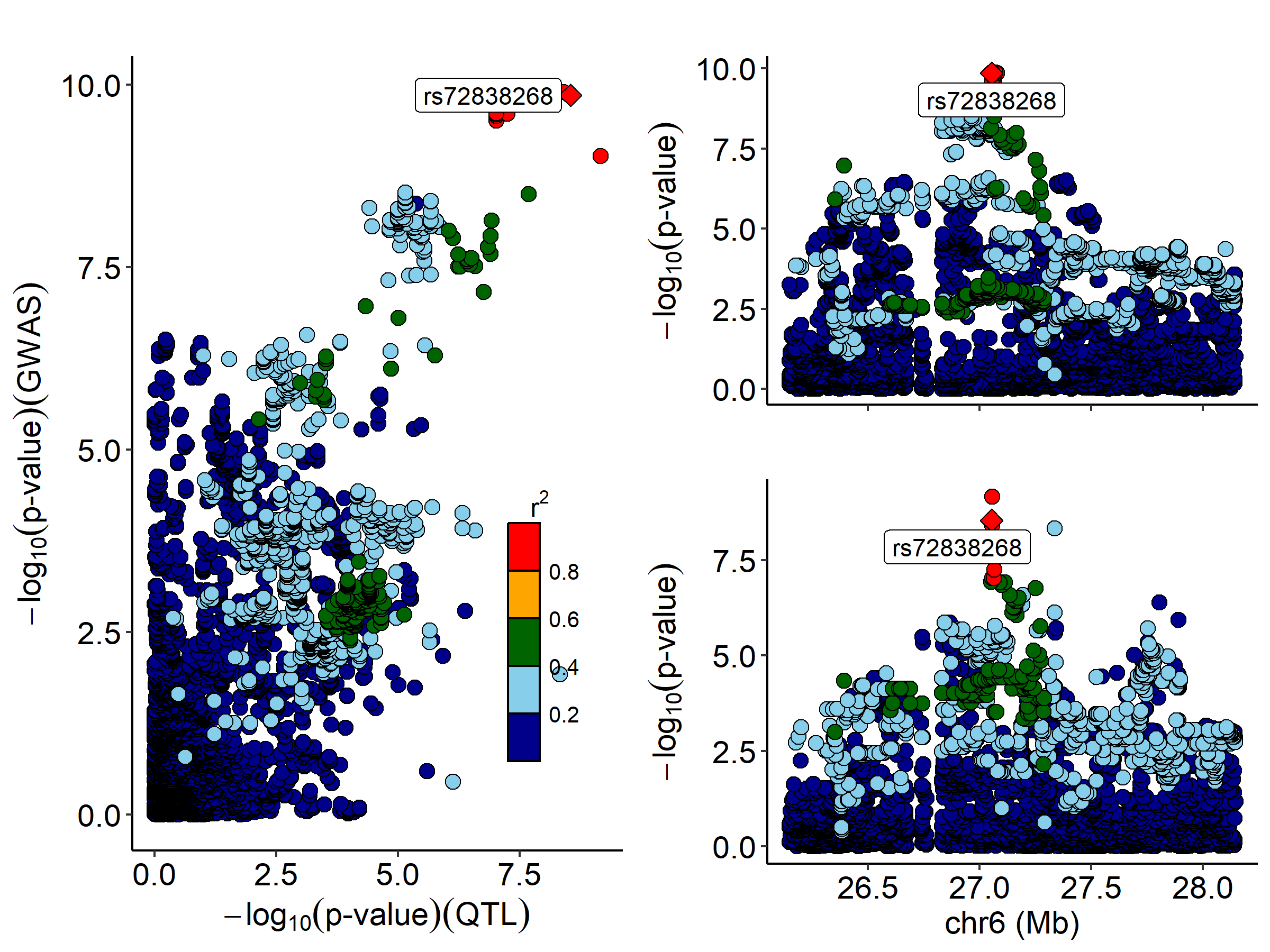

xi)

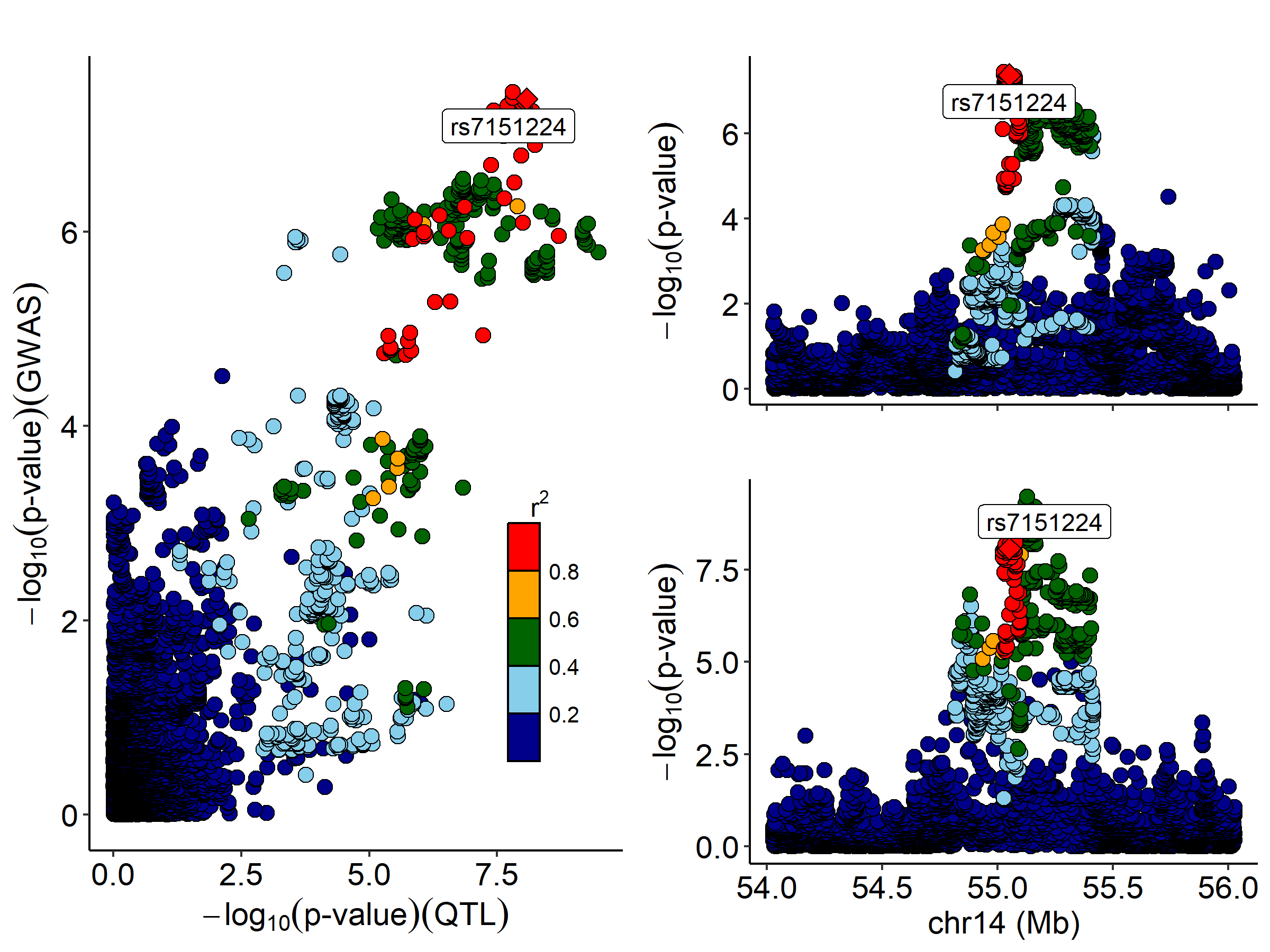

xii)

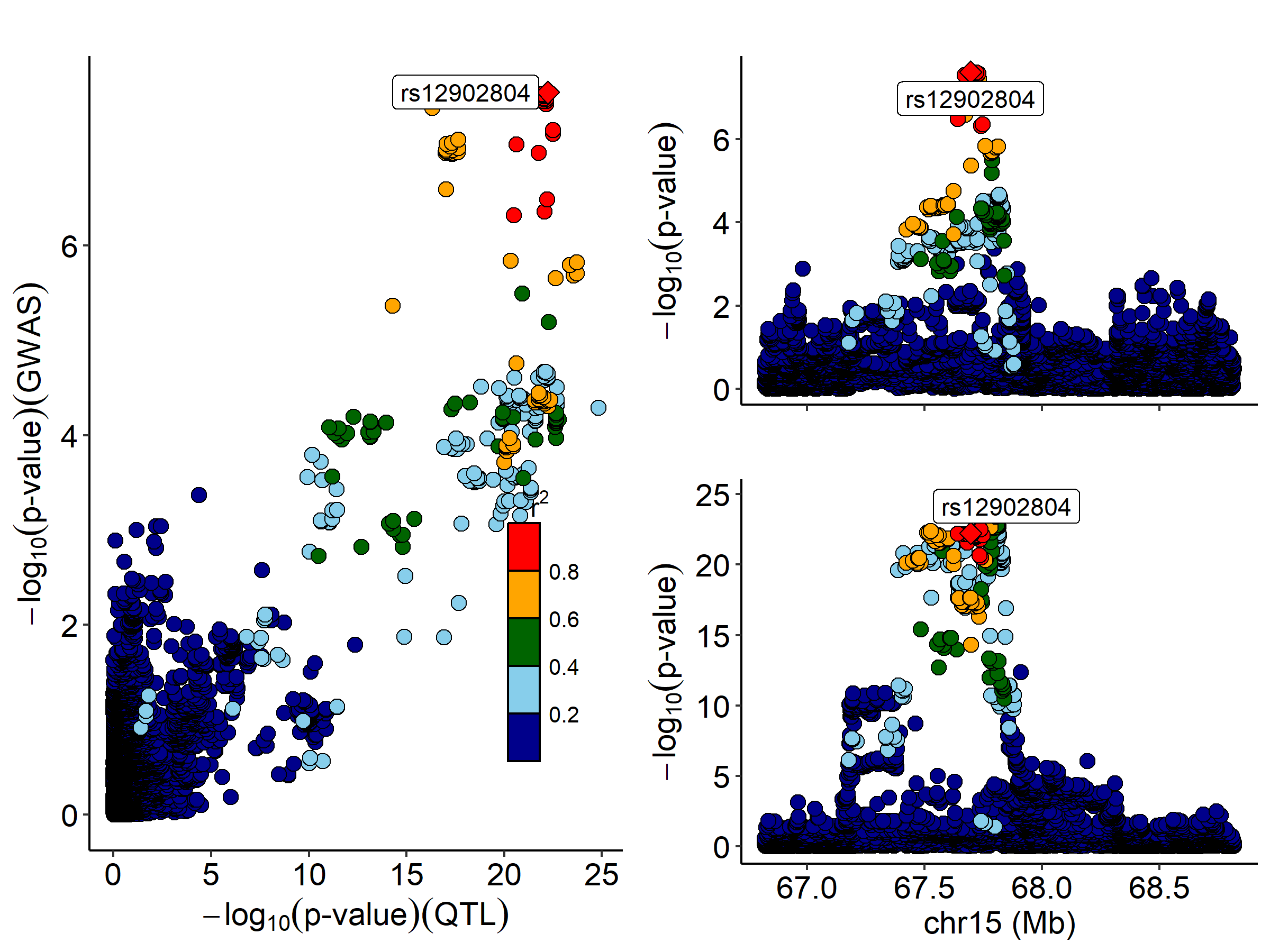

xiii)

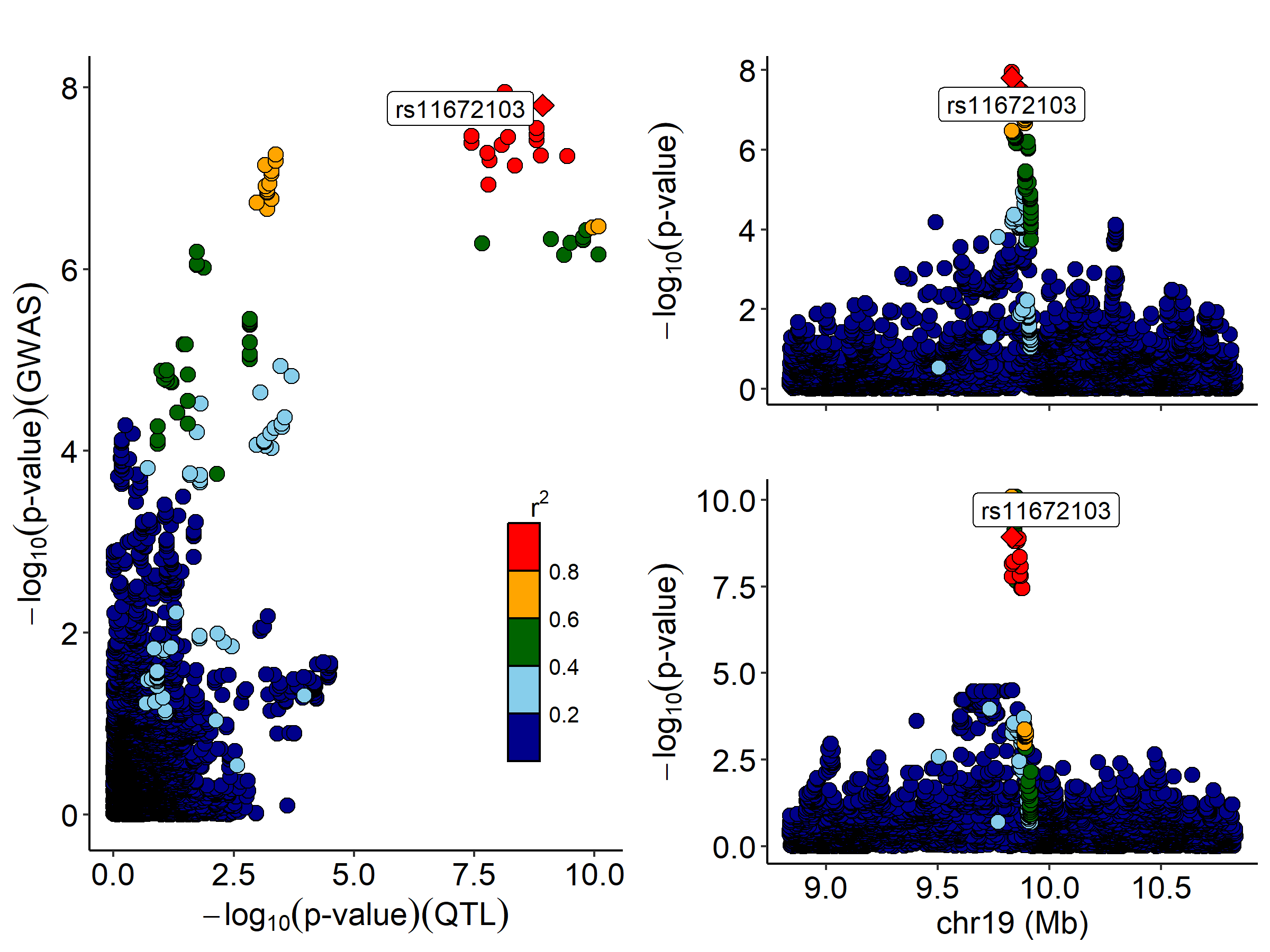
